# Effects of HIIT and HIIT plus Resistance Training on Cerebral Blood Flow and Other Health Outcomes in Individuals with Coronary Artery Disease: The Heart-Brain Randomized Controlled Trial

**DOI:** 10.64898/2026.03.23.26347205

**Authors:** Lucía Sánchez-Aranda, Angel Toval, Esmee A. Bakker, Patricio Solis-Urra, Isabel Martín-Fuentes, Javier Fernández-Ortega, Rosa María Alonso-Cuenca, Marcos Olvera-Rojas, Beatriz Fernandez-Gamez, Andrea Coca-Pulido, Alberto González-García, Darío Bellón, Alessandro Sclafani, Javier Sanchez-Martinez, Ricardo Rivera-López, Norberto Herrera-Gómez, Emilio J. Barranco-Moreno, Francisco J. Amaro-Gahete, Anna Carlén, Jairo H. Migueles, Danny JJ Wang, Kirk I. Erickson, Eduardo Moreno-Escobar, Rocío García-Orta, Irene Esteban-Cornejo, Francisco B. Ortega

**Author notes:** Correspondence: Francisco B. Ortega and Angel Toval, Department of Physical and Sports Education, Faculty of Sport Sciences, University of Granada, Granada, Spain. E-mail address and.

## Abstract

**Background:** Individuals with coronary artery disease (CAD) are at higher risk of cognitive decline and dementia, in which gray matter cerebral blood flow (CBF) plays a critical role. This study investigated the effects of High Intensity Interval Training (HIIT) and HIIT plus resistance training (RT) on CBF and other health outcomes in individuals with CAD.

**Methods:** This trial included 105 participants with CAD (age 62.1±6.6 years, 21% women) randomly assigned to HIIT+RT (n=37), HIIT (n=35) or usual care (n=33). The primary outcome was the change in global CBF from baseline to 12-week follow-up. Secondary outcomes included: region-specific CBF (hippocampus, precuneus, and anterior/posterior cingulate cortex), cognitive function (general cognition, episodic memory, processing speed, working memory and executive function/attentional control), peak oxygen uptake (VO_2_peak), muscular fitness (30s sit-to-stand) and body composition [weight, body mass index (BMI), and fat and muscle mass). Data were analyzed using available-case intention-to-treat constrained (baseline-adjusted) linear mixed models. Predefined subgroup analyses were conducted for age, sex, education, and baseline level of the outcome studied.

**Results:** No significant between-group differences were observed in CBF changes in the whole sample. However, participants with lower CBF at baseline showed greater CBF increases in the HIIT group compared to both usual care (+7.1 ml/100g/min, P=0.02) and HIIT+RT (+5.53 ml/100g/min, P=0.04). No effects were observed on regional CBF or cognition. Both exercise groups improved VO_2_peak compared to usual care (HIIT+RT: +2.6; HIIT: +2.5 mL/kg/min, both P<0.001). Only HIIT+RT increased muscular fitness (vs usual care: +3.3; vs HIIT: +3.1 repetitions, both P<0.001), and only HIIT decreased BMI (vs usual care: -0.47; vs HIIT+RT: -0.44 kg/m^2^, both P≤0.03). No life-threatening events or deaths occurred during 1995 training sessions in the exercise groups, nor in the usual care group.

**Conclusion:** Twelve weeks of HIIT+RT or HIIT did not increase CBF in the whole sample with CAD, but HIIT effectively increased CBF in those who had poorer CBF at baseline. While no cognitive benefits were observed, we found exercise-specific improvements in other clinically relevant outcomes, such as VO₂peak, muscular fitness, and BMI.

**Clinical Perspective:** *What’s new?:*

- This study investigated the effectiveness of two high intensity interval training (HIIT)-based programs on cerebral blood flow (CBF), cognition and other clinically relevant outcomes in individuals with coronary artery disease (CAD). This is particularly important given the accelerated cognitive decline, dementia risk, and inherent poorer cardiovascular health in individuals with CAD.
- This study introduces a novel exercise model by adapting the traditional 4×4 HIIT into a 3×4 format plus 2 rounds of 8 resistance exercises. The program is time-efficient (3 sessions/week of 45 minutes) and aligns with the World Health Organization recommendations, the American and European clinical guidelines for CAD management, and American College of Sports Medicine recommendations on resistance training prescription.

*What are the clinical implications?:*

- HIIT alone was effective in increasing CBF only in individuals with low levels of CBF at baseline, and in reducing BMI in the whole sample.
- Both HIIT+RT and HIIT equally improved cardiorespiratory fitness, whereas the HIIT+RT additionally improved muscular fitness.
- Both HIIT+RT and HIIT were safe in this population with CAD, with no major adverse events or death occurring during roughly 2000 sessions.

## Introduction

Individuals with cardiovascular disease (CVD) are at higher risk of cognitive decline^1,2^ and all-cause dementia.^3–5^ Particularly, coronary artery disease (CAD), the most prevalent type of CVD,^6^ has been established as a risk factor for accelerated cognitive decline and dementia.^7,8^ Decreases in cerebral blood flow (CBF), that entail a reduced supply of oxygen and nutrients to brain tissue,^9,10^ have been associated with an increased risk of dementia^11^ and cognitive decline.^12–14^

High-Intensity Interval Training (HIIT) has exhibited countless benefits for individuals with CAD, including: reduced mortality^15,16^ and improvements in cardiorespiratory fitness (CRF),^17–20^ mental health^21–23^ and quality of life.^17,18,23^ HIIT has shown superior efficacy compared to moderate-intensity continuous training (MICT) in improving peak oxygen uptake (VO_2_peak), both in the general population^24^ and in patients with CVD.^19,20,25^ In addition to aerobic exercise, emerging evidence and guidelines^26–31^ recommend including resistance training (RT) to optimize health benefits. RT has been demonstrated to be effective at improving hemodynamic outcomes such as blood pressure, arterial stiffness, and endothelial function.^28^ Based on this evidence, it is of particular interest to investigate the effect of modified HIIT protocols to integrate RT within the same session, as a novel and time-efficient method to meet guidelines and optimize benefits.

Exercise has been widely proposed as a promising strategy to slow down or prevent dementia and age-related cognitive decline.^32^ Nonetheless, a recent meta-analysis^23^ revealed that evidence on the effects of exercise on brain and cognitive outcomes in individuals with CAD is nearly non-existent, and to our knowledge, only one single-arm longitudinal study^33^ has examined its impact on CBF. To address this gap, the overall aim of the Heart-Brain trial is to investigate the effects of HIIT+RT and HIIT alone, compared to usual care, on global gray matter CBF (hereafter named just CBF) as the primary outcome, and other secondary health outcomes: regional CBF, cognition, CRF, muscular fitness and body composition. Additionally, we assessed changes in weekly moderate-vigorous physical activity (MVPA) and number of muscle-strengthening sessions in all the study groups as control for potential contamination and compensation effects.

## Methods

### Study design, participants, protocol registration and deviations

The Heart-Brain trial is a single-center, three-arm, single-blinded randomized controlled trial (RCT) including a total of 105 individuals with CAD (50-75 years old) from Granada (Spain). All data were collected between April 2022 and June 2024. The trial protocol was in accordance with the principles of the Declaration of Helsinki and was approved by the Research Ethics Board of the Andalusian Health Service (CEIM/CEI Provincial de Granada; #1776-N-21 on December 21st, 2021). All participants provided written informed consent. All research processes were conducted by the premises of the Singapore Statement on Research Integrity.^34^ The trial was registered on Clinicaltrials.gov (Identifier: NCT06214624; Submission date: December 22, 2023 url: https://clinicaltrials.gov/study/NCT06214624). The standard operating procedures for assessments and exercise intervention have been uploaded to public repositories.^35,36^ The statistical analysis plan (SAP) and the study protocol were uploaded to Clinicaltrials.gov before the last patient was enrolled (submission date: January 29th, 2024). There were no deviations in the study design, intervention, or methods used to assess the primary or secondary outcomes reported in this manuscript during the total duration of the study. There was only a slight deviation in the way the cognitive composite scores were computed for analyses to align with new evidence provided by a large RCT.^37^

The reporting of the results adheres to the 2025 Consolidated Standards of Reporting Trials Extension (CONSORT extension) guidelines^38^ (Table S1). A more extensive description of the methodology and protocols of the Heart-Brain project has been published elsewhere.^39^ Patient and public involvement actions have been integrated throughout the different phases of the study (Table S2).

### Eligibility criteria

In brief, eligibility criteria included: (i) individuals with chronic CAD and stable medical treatment, aged between 50-75 years, (ii) with left ventricular ejection fraction ≥45%, (iii) physically inactive,^26^ and (iv) with no major cognitive impairments according to the Spanish version of the modified Telephone Interview of Cognitive Status (STICS-m) (≥ 26 points).^40,41^ The full list of eligibility criteria has been published elsewhere^39^ and is provided in Table S3.

### Interventions and usual care

A full description of the Heart-Brain exercise interventions, following the Consensus on Exercise Reporting Template (CERT), can be found as a preprint.^42^ Briefly, participants were randomized using a 1:1:1 ratio and stratified by age (<65 or ≥65 years) and sex (female or male), to one of the following three groups: HIIT+RT, HIIT and usual care. Participants assigned to one of the intervention groups were asked to come to the Sport and Health University Research Institute (iMUDS) in Granada (Spain) for 45-min training sessions, 3 times per week, for 12 weeks. Both interventions were designed to be isotemporal and isocaloric.^42^ The HIIT parts were designed following the HIIT guidelines for clinical populations^43^ and were conducted preferably on a treadmill.

#### HIIT+RT Intervention

This intervention combined treadmill-based HIIT with RT using elastic bands and bodyweight exercises. Each session followed this structure: (i) a 5-minute warm-up involving walking with gradual increases in speed and/or incline; (ii) a HIIT phase consisting of 3 intervals of 4 minutes at high-intensity (85–95% of HRmax), each followed by a 3-minute recovery period (70% of HRmax); (iii) a 4-minute cool-down phase, followed by a 1-minute and 20-second transition period to get off the treadmill and walk slowly around the room (to mitigate any dizziness post-treadmill); (iv) two rounds of an 8-exercise RT circuit. Each exercise was performed for 20 seconds, with a 40-second rest between exercises and a 1-minute rest between circuits.

#### HIIT Intervention

The HIIT sessions followed this structure: (i) a 10-minute warm-up consisting of walking with a gradual increase in speed and/or incline, aiming to reach and maintain 70% of maximum heart rate (HRmax); (ii) a main phase of 4 high-intensity intervals (85–95% of HRmax), each lasting 4 minutes, with 3-minute active recovery periods (70% of HRmax) in between; (iii) a 10-minute cool-down phase.

#### Usual care group

The usual care group was treated as usual in outpatient Phase III, which in Spain includes periodic medical revisions where the clinician may provide patients with lifestyle and clinical advice and medication control. Participants were offered the supervised HIIT program described above after all pre-and post-intervention assessments were completed, i.e. waiting-list control group.

### Statistical Power and Sample Size

Power calculations were performed using G*Power (version 3.1.9.7. Universität Düsseldorf, Germany).^44^ The Heart-Brain RCT was designed to detect a low-to-medium effect size in the primary outcome, CBF (i.e., Cohen’s d = 0.35)^33^ for repeated-measures, within-between interaction, with a two-sided alpha of 5% and power of 80%. The sample size was adjusted for an expected 7% dropout during the intervention (based on 3-month dropout rates of our previous RCTs).^45^ The estimated sample size was 90 participants (30 in each study arm) for primary outcome analyses (MRI-based CBF). We aimed to enroll additional participants anticipating that some might feel claustrophobic during the MRI scanning and that some imaging data might be discarded after quality control (15% loss estimated based on experience in previous studies), so that 105 participants was the final recruitment target.

### Outcome measurements

The primary and secondary outcomes were assessed at baseline and post-intervention. Weekly MVPA and number of muscle-strengthening sessions were assessed to control changes in these intervention-related behaviors across the three study arms.

#### Primary outcome: change in CBF

MRI data were acquired with a 3.0 Tesla Siemens Magnetom Prisma scanner (Siemens Medical Solutions, Erlangen, Germany). Background-suppressed four-delay pseudo-continuous arterial spin labelling (pCASL), based on the protocol by Wang et al.,^46^ was performed to obtain CBF (ml/100g/min) measurements. A 3D T1 weighted image was acquired and processed for registration and segmentation purposes.

First, T1 weighted scans were preprocessed using FSL’s ANAT pipeline.^47^ Then, T1 weighted and arterial spin labelling (ASL) images were corregistered.^48–51^ Finally, ASL data and FSL’s ANAT outputs were entered into the *oxford_asl* command, which estimates CBF.^52,53^ Partial volume correction was used to report CBF.^54^ Further details on MRI acquisition, processing and visual inspection and quality control procedures can be found in Supplemental methods. *Regional CBF*.

Bilateral parcellations from FreeSurfer’s analyses^55^ were used to determine regional CBF (Supplementary material 1, section 4). Due to high correlation between hemispheres (r Pearson correlation values between 0.8 and 0.9, p<0.001) for the regions included (hippocampus, posterior cingulate cortex, anterior cingulate cortex and precuneus), values from both hemispheres were averaged.

#### General cognition and cognitive domains: executive function/attentional control, episodic memory, processing speed, and working memory

The participants completed a comprehensive neuropsychological test battery assessing different cognitive domains: (i) general cognition,^56^ (ii) episodic memory,^56,57^ (iii) processing speed,^58,59^ (iv) working memory,^60,61^ and (v) executive function / attentional control.^57,59^ Further details are provided in Supplemental methods.

#### Cardiorespiratory fitness

The participants performed a standardized ramp incremental cardiopulmonary exercise test (CPET) on a treadmill (h/p/cosmos, Nussdorf, Germany), according to the American College of Sports Medicine (ACSM) guidelines.^62^ Respiratory gases were registered with a dilution flow system (Omnical, Maastricht Instruments, Maastricht, the Netherlands). Relative VO_2_peak (i.e. body mass normalized - mL/kg/min) was used as CRF indicator. Further details are provided in Supplemental methods and have been previously described elsewhere.^63^

#### Muscular fitness

The 30-s sit-to-stand test was performed as described in the Senior Fitness Test battery procedures.^64^ This test assesses lower-body muscular endurance and is widely recognized as an indicator of overall muscular fitness, functional capacity, balance and coordination status, as well as CRF.^65^

#### Body composition

A SECA 225 (Seca, Hamburg, Germany) stadiometer and scale was used to measure height and weight, and a bioimpedance scale (Tanita MC-980MA-N PLUS, Tanita Europe B.V., Amsterdam, The Netherlands) was employed for fat and muscle mass assessments. Body mass index (BMI), fat mass index (FMI) and muscle mass index (MMI) were later computed, dividing the corresponding mass in kilograms by the squared height in meters.

#### MVPA and muscle-strengthening training sessions

For measuring MVPA (at baseline and mid-point-6^th^ week) participants were asked to wear an Axivity AX3 accelerometer (Axivity Ltd., New Castle Upon Tyne, United Kingdom) for 10 consecutive days. Weekly MVPA was calculated using the GGIR package^66^ (version 3.1-2) in R (version 4.4.1) and defined as the daily time accumulated above 100m*g* of acceleration,^67,68^ considering only bouts longer than 1 minute to remove acceleration noise. Further details on accelerometer measures can be found elsewhere.^69^ Muscle-strengthening sessions were defined as weekly frequency of self-reported weightlifting or body weight strength training sessions and assessed at baseline and post-assessments using the questionnaire employed in the IMPACT cohort (https://cohorte-impact.es/en)^70^

#### Moderation outcomes

Sex (male vs female), age (<65 vs ≥ 65 years), education (primary and secondary vs university studies), and baseline level of the outcome (below/above median), were used as moderators as predefined in our SAP. Further details are provided in Supplemental methods.

### Attendance and compliance

Attendance was evaluated as the proportion of sessions completed out of the total sessions offered to the participant. Participants with ≥70% attendance were included in the per-protocol (PP) analysis.^39^

Compliance was defined as the percentage of exercise sessions in which participants reached the required duration at the target heart rate intensity (≥85% of HRmax, derived from CPET), specifically >4.5 minutes for the HIIT+RT group, and >6.5 minutes for the HIIT group in accordance with the Guidelines for HIIT Prescription and Monitoring in clinical populations.^43^ For that, participants wore a chest strap (Polar H10, Kempele, Finland) connected to a sports watch (Polar Ignite 2, Kempele, Finland), we monitored second by second heart rate (HR) in the training sessions (+3.5 million data points analyzed).^42^

### Adverse events

All adverse events were recorded in REDCap,^71,72^ and classified according to the Common Terminology Criteria for Adverse Events (CTCAE)^73^ by an experienced cardiologist (EME). The 3-point major adverse cardiovascular events (MACE) definition was applied.^74^ An event was considered likely related to the exercise program when it occurred during the training session or within the 3 hours thereafter.^75^

### Statistical analyses

The analysis conducted is based on the predefined SAP published in Clinicaltrials.gov before completion of the study, and all analysis scripts and code used in this study will be deposited in a public GitHub repository.^35^ The primary analytical approach to test the intervention effect was based on intention-to-treat (ITT) analyses performed on available cases, which include all randomized participants with valid data for at least one time point. As a secondary analysis, PP analyses were conducted for the primary and secondary outcomes, in participants attending at least 70% of sessions. Moreover, additional sensitivity analyses were performed based on the PP data by excluding: (i) participants who did not achieve ≥70% of sessions based on time in heart rate target zones, and (ii) participants who experienced adverse events at post-assessments, had deviations from the exercise protocol, or underwent interruption periods due to adverse events.

Treatment effects were evaluated by examining between-group differences (HIIT+RT vs usual care; HIIT vs usual care; HIIT+RT vs HIIT) in the change from baseline to 12-weeks in primary and secondary outcomes (except for MVPA, evaluated from baseline to mid-point). For this purpose, we used Constrained (i.e., baseline-adjusted) Linear Mixed Models (CLMM), specifically with *lme4*^76^ *and lmerTest* packages.^77^ The models included time as a categorical fixed effect, and group-by-time interaction, with the intercept specified as a random effect. Statistical significance was set at P<0.05. The intervention effect was represented by the time x group interaction coefficient, along with 95% confidence intervals (CIs) and p-values. Estimated marginal means and the change over time within each arm were calculated by the *emmeans* package.^78^ Exploratory subgroup and interaction analyses (i.e., moderation analyses) were prespecified in the SAP and performed to examine whether the key variables age, sex, education level and baseline level of the study outcome modified the effect of the intervention. All analyses were performed using R version 4.5.1. Further details on statistical analyses can be found in Supplemental methods.

## Results

A total of 252 participants were initially invited to participate based on medical records meeting clinical eligibility criteria. After additional eligibility checks during baseline evaluations, 105 participants were randomized. A flowchart with specific numbers and reason for eligibility exclusions is shown in Figure 1. The participants’ baseline characteristics are shown in Table 1. The mean age at baseline was 62.1±6.6, 21% were female. No significant differences were found between dropout and non-dropout participants for the primary outcome (Table S5). The intervention was well accepted by the participants, as shown by a high attendance (92%), compliance (90%) and reported enjoyment (5.4/7), as described in the exercise protocol article.^42^ Median attendance was 6% higher in the HIIT+RT than in the HIIT group, without differences in compliance or enjoyment between exercise groups.^42^

**Figure 1.**
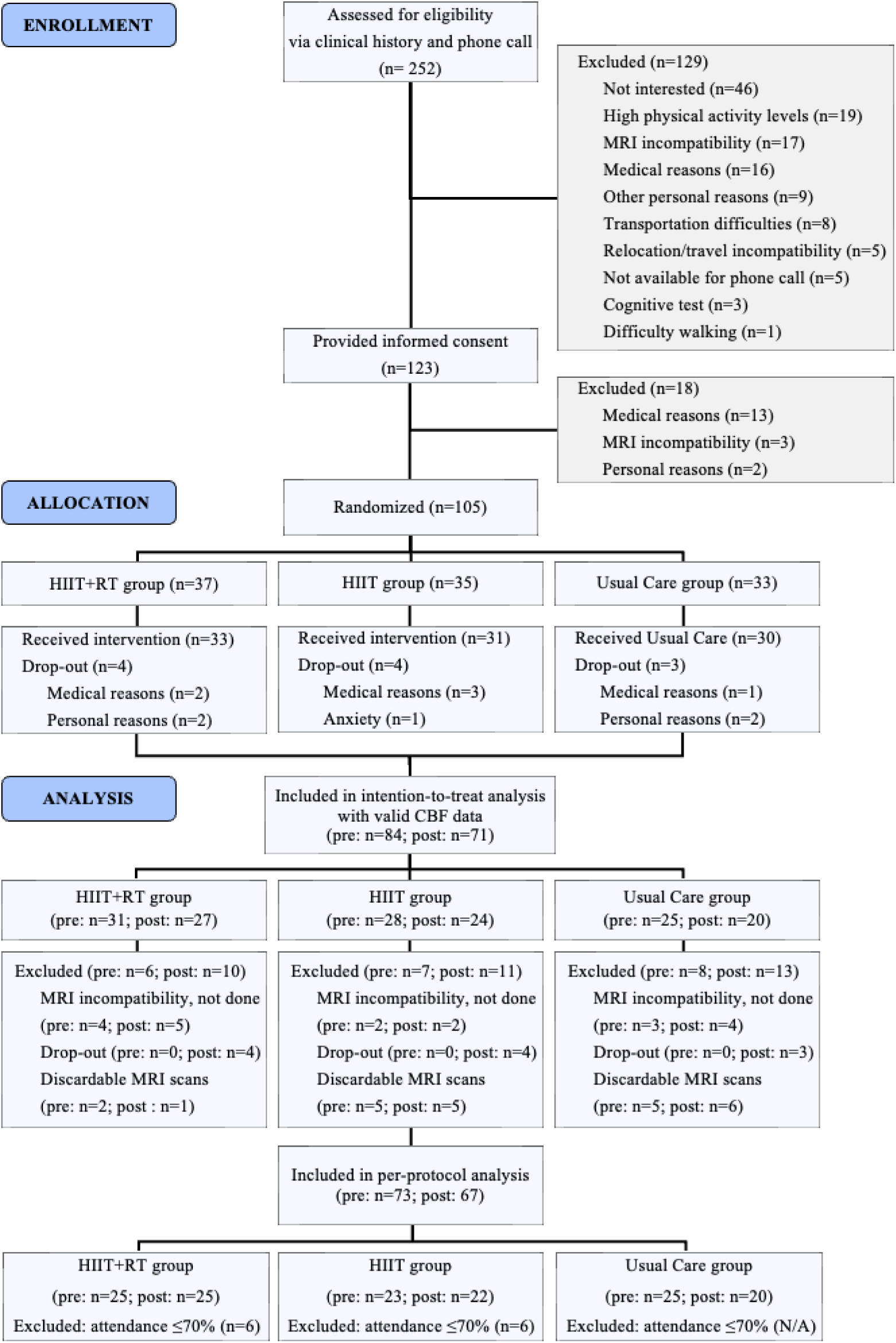
CONSORT flow diagram from the Heart-Brain trial. N/A: not applicable. Analysis numbers correspond to the primary outcome (cerebral blood flow). Drop-out participants were inherently excluded from the per-protocol analysis considered as not meeting attendance. Abbreviations: CBF, cerebral blood flow; HIIT, high intensity interval training; HIIT+RT, high intensity interval training + resistance training; MRI, magnetic resonance image; SMD, standardized mean difference.

**Table 1.** Baseline characteristics of the Heart-Brain sample.

| Characteristic | Overall N = 105<sup>I</sup> | HIIT+RT N = 37<sup>I</sup> | HIIT N = 35<sup>I</sup> | Usual care N = 33<sup>I</sup> |
| --- | --- | --- | --- | --- |
| Age | 62.13 (6.59) | 62.00 (6.99) | 61.91 (6.55) | 62.52 (6.34) |
| Sex, n (%) |  |  |  |  |
| Male | 83 (79%) | 29 (78%) | 27 (77%) | 27 (82%) |
| Female | 22 (21%) | 8 (22%) | 8 (23%) | 6 (18%) |
| Education level, n (%) |  |  |  |  |
| Primary and secondary school | 63 (60%) | 23 (62%) | 23 (66%) | 17 (52%) |
| University | 42 (40%) | 14 (38%) | 12 (34%) | 16 (48%) |
| Weight (kg) | 82.88 (12.78) | 84.74 (12.59) | 83.06 (13.33) | 82.84 (12.61) |
| Height (cm) | 168.66 (8.64) | 168.79 (8.65) | 169.10 (10.03) | 168.05 (6.98) |
| Body mass index (kg/m2) | 29.1 (3.9) | 29.0 (3.6) | 29.1 (4.3) | 29.2 (3.7) |
| Body mass index categories, n (%) |  |  |  |  |
| Normal weight | 9 (8.6%) | 3 (8.1%) | 4 (11%) | 2 (6.1%) |
| Overweight | 59 (56%) | 22 (59%) | 18 (51%) | 19 (58%) |
| Obesity | 37 (35%) | 12 (32%) | 13 (37%) | 12 (36%) |
| Muscle mass (kg) | 55.53 (8.52) | 54.92 (8.53) | 56.00 (9.28) | 55.71 (7.87) |
| Fat mass (kg) | 24.13 (6.79) | 24.02 (6.82) | 24.15 (7.03) | 24.22 (6.72) |
| Muscle mass index (kg/m2) | 19.40 (1.79) | 19.19 (1.59) | 19.49 (2.03) | 19.55 (1.77) |
| Fat mass index (kg/m2) | 8.54 (2.62) | 8.48 (2.59) | 8.57 (2.83) | 8.56 (2.49) |
| Systolic blood pressure (mmHg) | 125.25 (14.84) | 125.19 (14.83) | 127.88 (14.24) | 122.56(15.20) |
| Diastolic blood pressure (mmHg) | 77.70 (9.05) | 77.93 (9.14) | 78.82(10.38) | 76.29 (7.23) |
| Anti-hypertensive treatment, n (%) | 84 (80%) | 28 (76%) | 28 (80%) | 28 (85%) |
| Lipid-lowering treatment, n (%) | 104 (99%) | 37 (100%) | 35 (100%) | 32 (97%) |
| Betablockers treatment, n (%) | 80 (76%) | 28 (76%) | 26 (74%) | 26 (79%) |
| APOE4 carrier, n (%) | 19 (18%) | 8 (22%) | 5 (14%) | 6 (18%) |
| VO<sub>2</sub>peak(ml/kg/min)<sup>2</sup> | 25.23 (5.01) | 25.18 (4.91) | 26.00 (5.16) | 24.51 (5.02) |
| 30s Sit-to-stand (repetitions) | 16.80 (4.32) | 16.22 (3.99) | 17.31 (4.26) | 16.91 (4.75) |
| Moderate-vigorous physical activity (minutes/week), median (Q1, Q3) | 320.76 (193.76, 452.28) | 326.46 (200.16, 437.63) | 373.46 (237.27, 477.02) | 298.82 (156.94, 362.63) |
| Self-reported muscle-strengthening (sessions/week), median (Q1, Q3) | 0.00 (0.00, 3.00) | 0.00 (0.00, 3.00) | 0.00 (0.00, 4.00) | 0.00 (0.00, 3.00) |
| CBF (ml/100g/min) | 54.87 (10.42) | 54.06 (10.12) | 58.04 (9.93) | 52.31 (10.80) |
| Hippocampus CBF (ml/100g/min) | 42.78 (8.40) | 43.99 (8.21) | 42.20 (8.50) | 42.12 (8.68) |
| Precuneus CBF (ml/100g/min) | 48.32 (10.47) | 51.34 (9.90) | 47.07 (9.97) | 46.47 (11.34) |
| Posterior cingulate cortex CBF (ml/100g/min) | 56.94 (11.29) | 59.83 (10.88) | 54.39 (11.66) | 56.88 (10.93) |
| Anterior cingulate cortex CBF (ml/100g/min) | 47.59 (9.74) | 49.37 (9.75) | 46.50 (10.21) | 46.94 (9.23) |
| General cognition - Montreal Cognitive Assessment (MoCA) Total score | 24.79 (3.25) | 24.59 (3.63) | 24.49 (3.41) | 25.33 (2.60) |
| Episodic memory (z-score) | 0.00 (1.00) | 0.11 (1.01) | -0.19 (1.04) | 0.08 (0.95) |
| Processing speed (z-score) | 0.00 (1.00) | -0.05 (1.02) | -0.06 (1.17) | 0.12 (0.77) |

| Characteristic | Overall N = 105<sup>1</sup> | HIIT+RT N = 37<sup>1</sup> | HIIT N = 35<sup>1</sup> | Usual care N = 33<sup>1</sup> |
| --- | --- | --- | --- | --- |
| Working memory (z-score)<sup>2</sup> | 0.00 (1.00) | -0.08 (0.98) | -0.02 (1.15) | 0.12 (0.86) |
| Executive function & attentional control (z-score) | 0.00 (1.00) | -0.01 (0.96) | -0.12 (1.26) | 0.13 (0.71) |

Data is presented as mean (SD) unless otherwise indicated.
<sup>1</sup>Due to missing data, working memory: n = 103 (HIIT+RT= 37, HIIT= 34, Usual care= 32), VO<sub>2</sub>peak: n = 103 (HIIT+RT= 37, HIIT= 33, Usual care= 33), CBF related variables: n = 84 (HIIT+RT= 31, HIIT= 28, Usual care= 25), Moderate-vigorous physical activity: n = 104 (HIIT+RT= 36, HIIT= 35, Usual care= 33), Muscle-strengthening: n = 98 (HIIT+RT= 33, HIIT= 35, Usual care= 30); Fat mass, muscle mass, fat mass index and muscle mass index: n = 104 ( HIIT+RT= 36, HIIT= 35, Usual care= 33).
Abbreviations: APOE, apolipoprotein E; CBF, cerebral blood flow; HIIT, High Intensity Interval Training; MoCA, Montreal Cognitive Assessment; RT, Resistance training; VO<sub>2</sub>peak, peak oxygen uptake

### Effects on primary outcome

We did not observe between-group differences in CBF after the 12-week intervention (HIIT+RT vs usual care: SMD= -0.02(-0.48,0.45), P=0.95; HIIT vs usual care: SMD= 0.02(-0.47,0.51), P=0.95; HIIT+RT vs HIIT: SMD= -0.03(-0.47, 0.41), P=0.89; Figure 2A, Table S6). PP and sensitivity analysis (PP+adverse events, PP+compliance) were in line with ITT primary analyses (Table S7, Figure S1).

**Figure 2.**
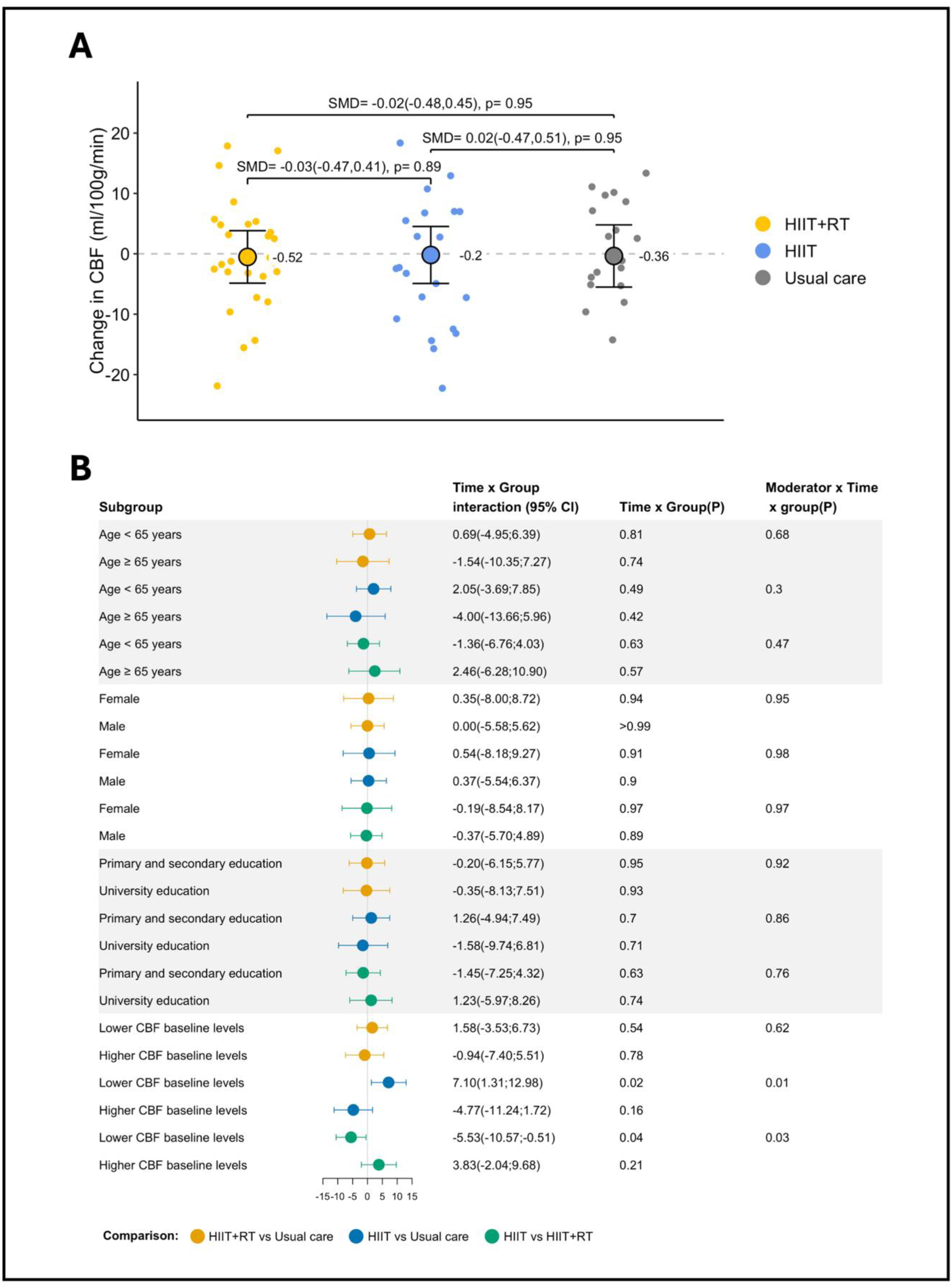
Intention-to-treat effects of the 12-week interventions on CBF by intervention group (A) and for subgroups (B). A: shows adjusted mean change (value shown to the right of the dot) over time and 95% Confidence Intervals by intervention groups, dots cloud indicates participant individual raw change values, and standardized mean differences and p-values are shown above the plots, connecting the corresponding study groups. B: dots indicate adjusted mean difference of marginal means and 95% confidence intervals between HIIT, HIIT+RT and usual care in each subgroup. Abbreviations: CBF, cerebral blood flow; CI, confidence intervals; HIIT, high intensity interval training; HIIT+RT, high intensity interval training + resistance training; SMD, standardized mean difference.

The predefined moderation analyses showed a significant interaction between CBF baseline levels, intervention group and time (P<0.05), such that participants assigned to HIIT demonstrated a greater increase in CBF in the subsample with low baseline CBF (vs usual care: 7.10 ml/100g/min, P=0.02; vs HIIT+RT: 5.53 ml/100g/min, P=0.04). There were no significant interactions with age, sex, or education (Figure 2B).

### Effects on secondary outcomes

No group differences were observed for the change in CBF at specific regions of interests (i.e., hippocampus, precuneus and posterior cingulate cortex; all P>0.3 Figure 3, Table S6). Exploratory analyses in all brain regions computed (a total of 42 regions by 2 hemispheres, n=84 measures), white matter CBF and arterial transit time, revealed no significant intervention effects. No intervention effects were observed either for CBF when using a voxel-wise whole brain approach (data not shown). There were no significant differences among study groups for measures of general cognition or any of the other cognitive domains (all P≥0.05; Figure 4, Table S8).

**Figure 3.**
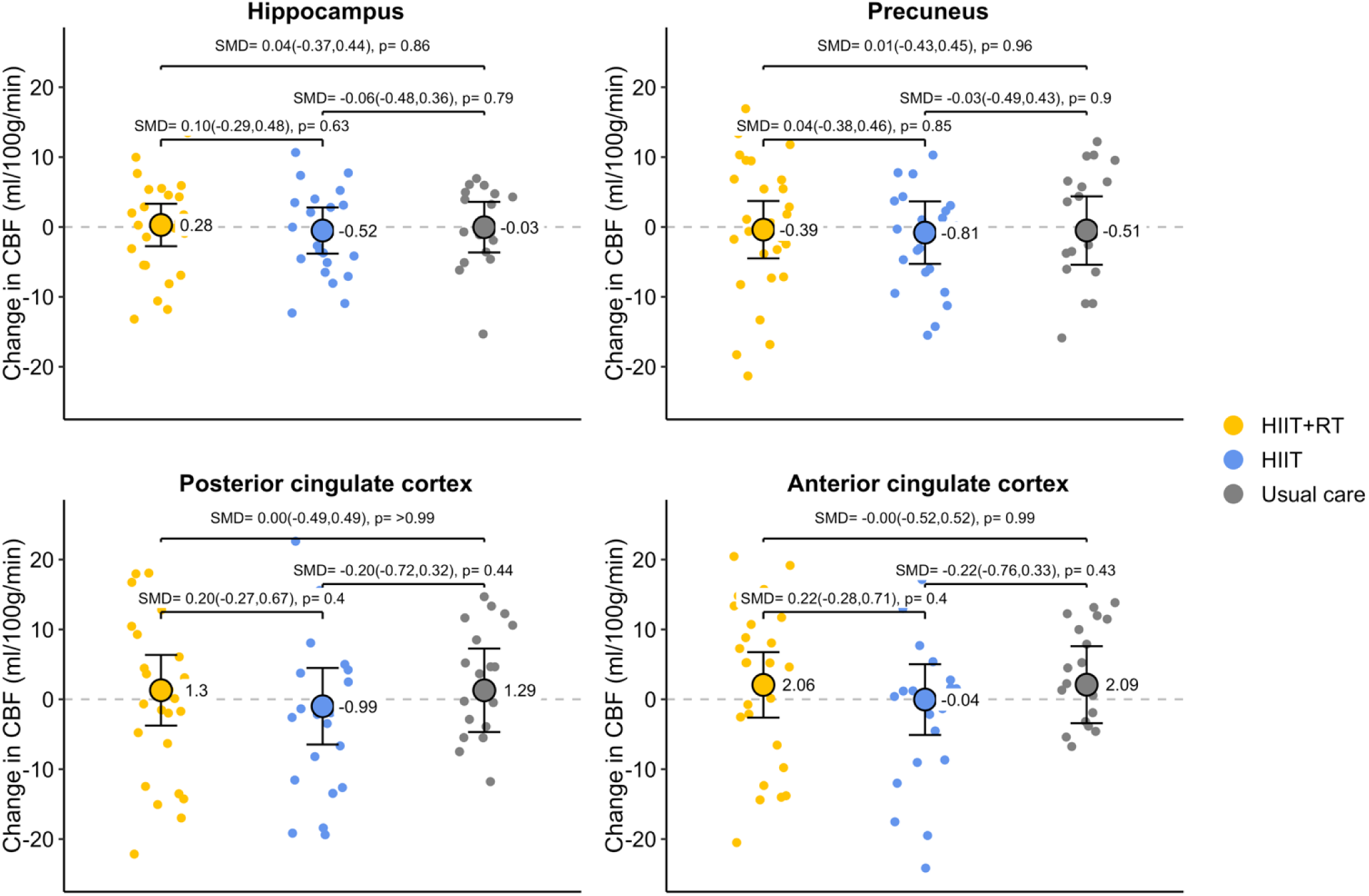
Intention-to-treat effects of the 12-week interventions on regional CBF by intervention group (HIIT+RT, HIIT, usual care). The figure shows adjusted mean change (value shown to the right of the dot) over time and 95% Confidence Intervals by intervention groups, and dots cloud indicate participants individual raw change values. Standardized mean differences and p-values are shown above the plots, connecting the corresponding study groups. Abbreviations: CBF, cerebral blood flow; HIIT, high intensity interval training; HIIT+RT, high intensity interval training + resistance training; SMD, standardized mean difference.

**Figure 4.**
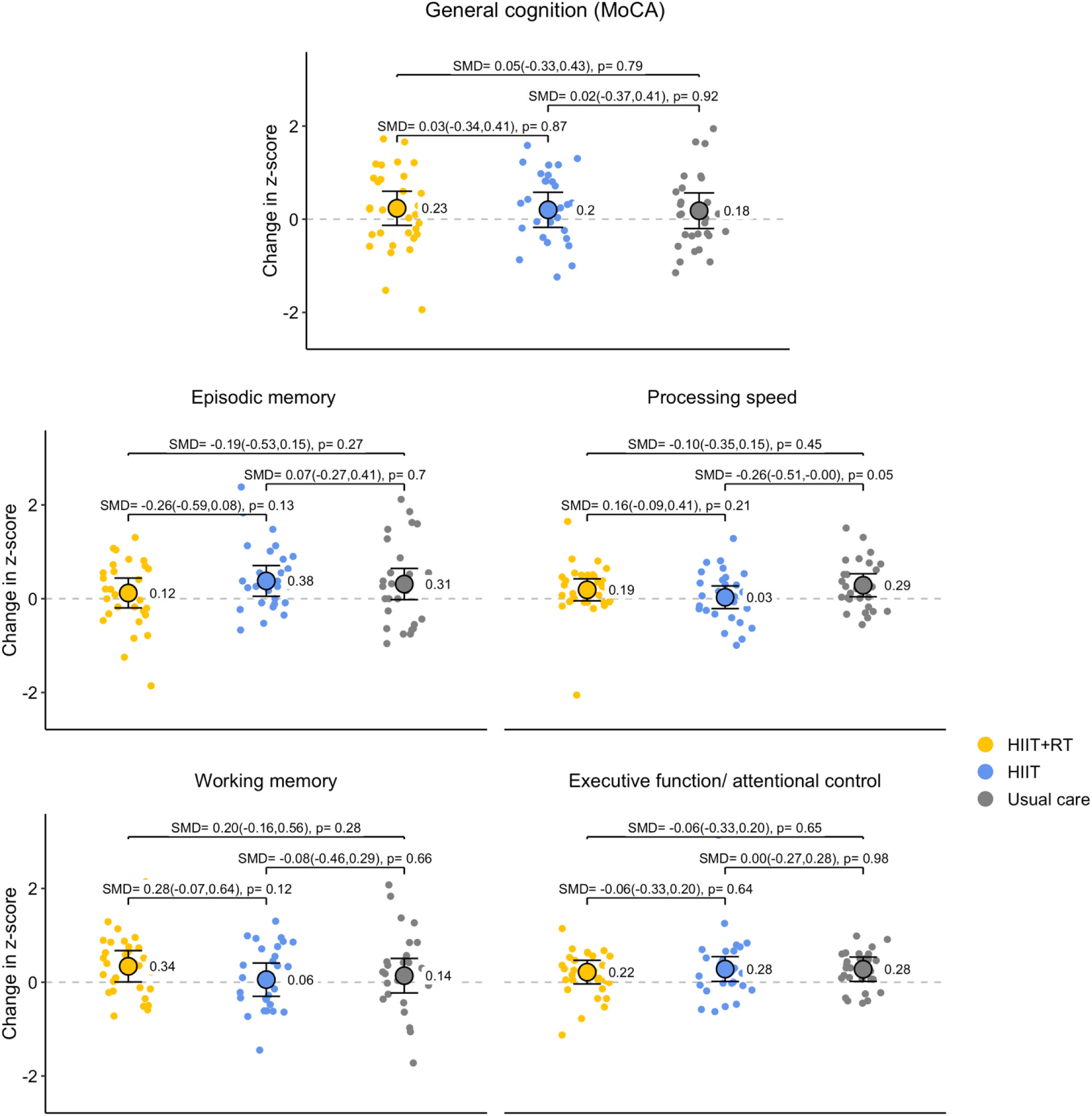
Intention-to-treat effects of the 12-week exercise interventions on general cognition (Montreal cognitive assessment – MoCA score) and cognitive sub-domains (episodic memory, processing speed, working memory, executive function/attentional control) z-scores by intervention group (HIIT+RT, HIIT, usual care). The figure shows adjusted mean change (value shown to the right of the dot) over time and 95% Confidence Intervals by intervention groups, and dots cloud indicate participants individual z-score change values. Standardized mean differences and p-values are shown above the plots, connecting the corresponding study groups. Abbreviations: HIIT, high intensity interval training; HIIT+RT, high intensity interval training + resistance training; MoCA, Montreal cognitive assessment; SMD, standardized mean difference.

On the other hand, both exercise groups significantly improved VO_2_peak (HIIT+RT: +2.6; HIIT:+2.5 mL/kg/min) as compared with usual care (HIIT+RT: SMD= 0.52, P<0.001; HIIT: SMD= 0.50, P<0.001), with no differences between HIIT+RT and HIIT exercise groups (Figure 5, Table S8). Moreover, only the HIIT+RT group significantly improved performance in the 30s sit-to-stand test compared to the usual care group (+3.3 repetitions, SMD= 0.77, P<0.001) and the HIIT group (+3.1 repetitions, SMD= 0.72, P<0.001) (Figure 5, Table S8). On the other hand, BMI only decreased in the HIIT group (-0.4 kg/m^2^) compared to usual care (SMD= - 0.12, P=0.02) and the HIIT+RT (SMD= 0.11, P=0.03) groups. No other significant effects were found for fat or muscle mass, with or without accounting for body size (i.e., FMI and MMI) (Figure 6, Table S8).

**Figure 5.**
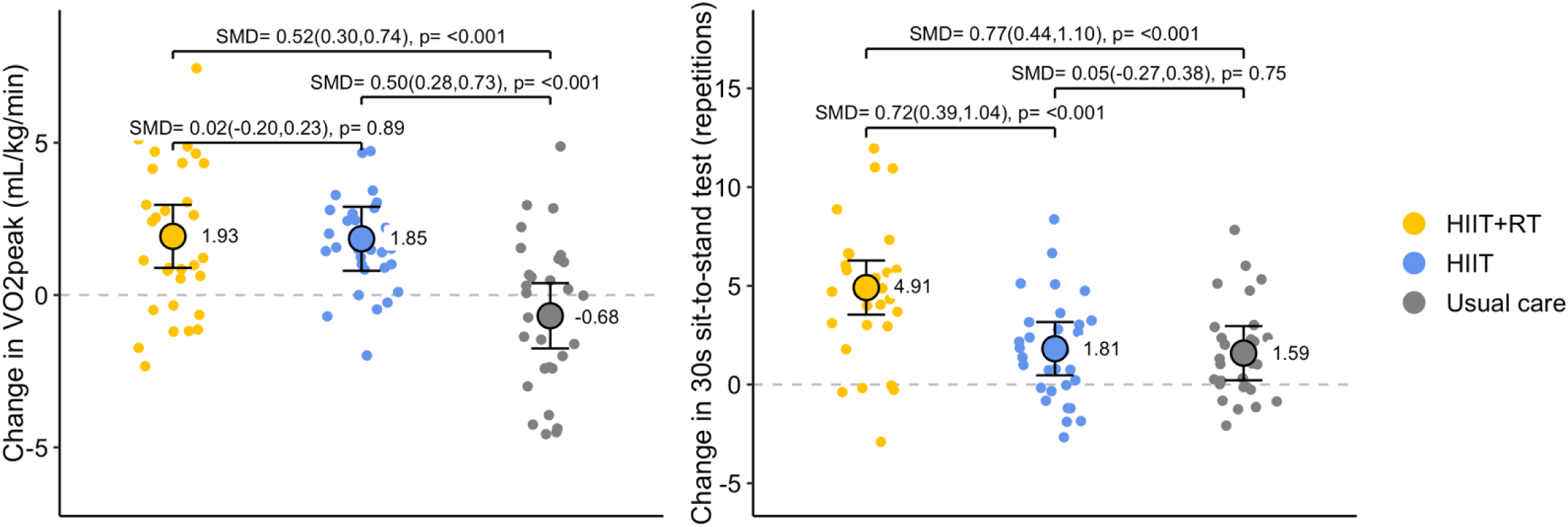
Intention-to-treat effects of the 12-week exercise interventions on peak oxygen uptake (VO_2_peak) and 30s sit-to-stand repetitions, by intervention group (HIIT+RT, HIIT, usual care). The figure shows adjusted mean change (value shown to the right of the dot) over time and 95% Confidence Intervals by intervention groups, and dots cloud indicate participant individual raw change values. Standardized mean differences and p-values are shown above the plots, connecting the corresponding study groups. Abbreviations: HIIT, high intensity interval training; HIIT+RT, high intensity interval training + resistance training; SMD, standardized mean difference; VO2peak, peak oxygen uptake.

**Figure 6.**
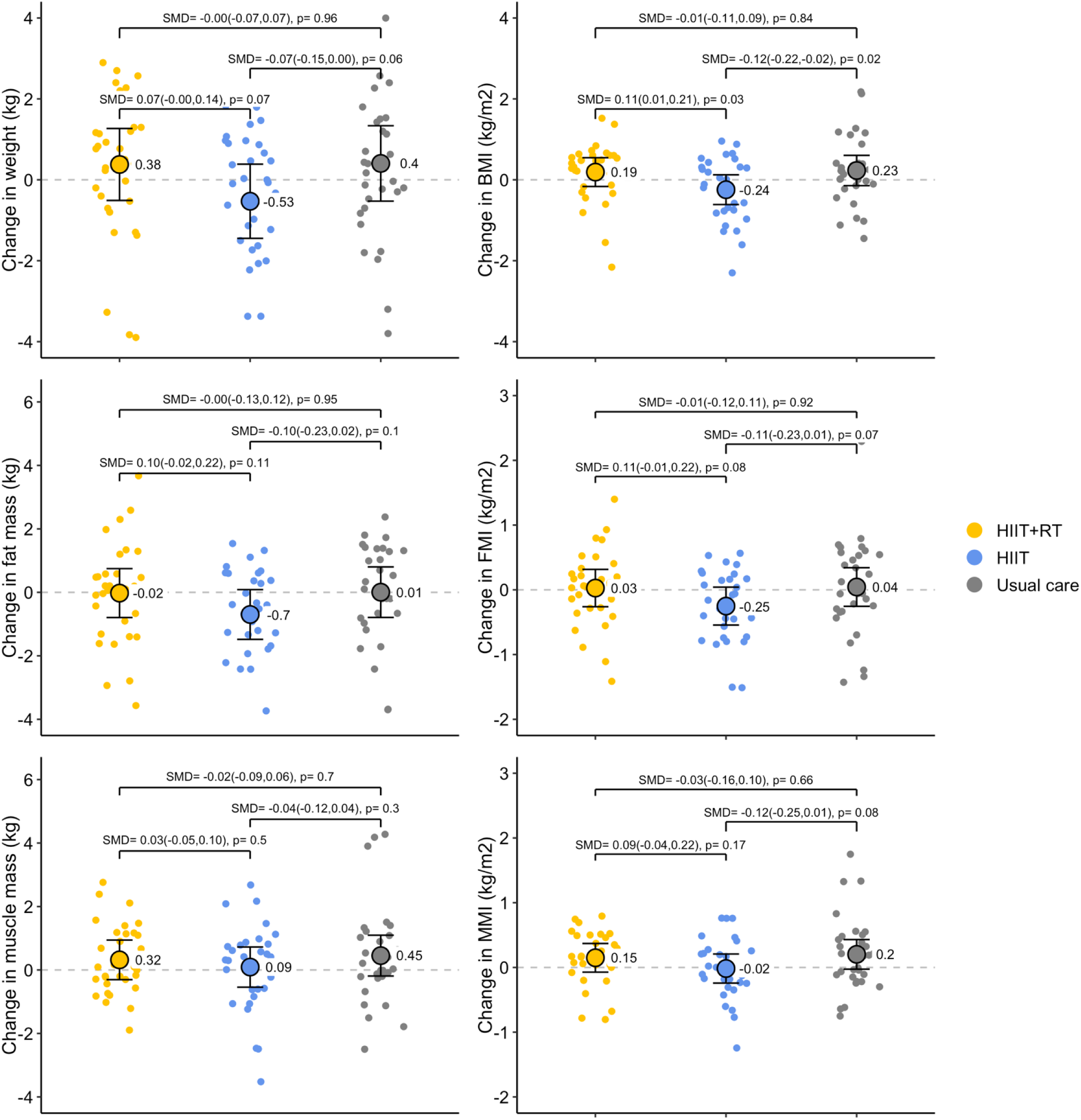
Intention-to-treat effects of the 12-week exercise interventions on body weight, body mass index (BMI), fat mass, fat mass index (FMI), muscle mass and muscle mass index (MMI) by intervention group (HIIT+RT, HIIT, usual care). The figure shows adjusted mean change (value shown to the right of the dot) over time and 95% Confidence Intervals by intervention groups, and dots cloud indicate participant individual raw change values. Standardized mean differences and p-values are shown above the plots, connecting the corresponding study groups. Abbreviations: HIIT, high intensity interval training; HIIT+RT, high intensity interval training + resistance training; SMD, standardized mean difference.

PP analyses conducted for secondary outcomes were consistent with the ITT primary analyses (Table S7). Further, there was no evidence of interaction between the intervention and the moderators studied (Table S10).

### Changes observed in MVPA and self-reported muscle-strengthening sessions during the intervention

The HIIT+RT group increased MVPA from baseline to mid-intervention in 40 min/week (P=0.28) and the HIIT group in 124 min/week (P=0.001) compared to usual care (Figure 7, Table S8). HIIT+RT reported an increase in weekly muscle-strengthening sessions when compared with the other two arms (vs usual care: +3.08 and vs HIIT: +2.34 sessions/week; both P<0.001; Figure 7, Table S8).

**Figure 7.**
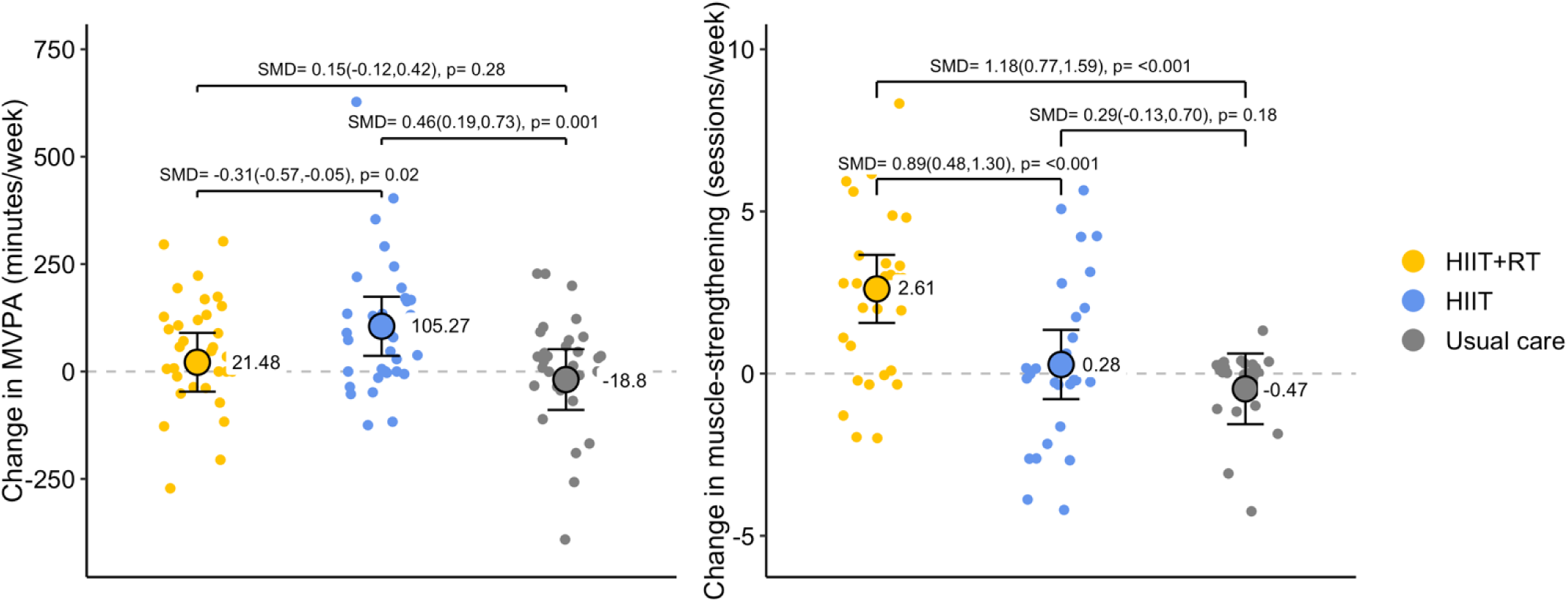
Changes observed (intention-to-treat analyses) on moderate to vigorous physical activity (MVPA) at 6^th^ week and muscle-strengthening sessions at post-assessments, by intervention group (HIIT+RT, HIIT, usual care). The figure shows adjusted mean change (value shown to the right of the dot) over time and 95% Confidence Intervals by intervention groups, and dots cloud indicate participant individual raw change values. Standardized mean differences and p-values are shown above the plots, connecting the corresponding study groups. Abbreviations: HIIT, high intensity interval training; HIIT+RT, high intensity interval training + resistance training; MVPA, moderate-vigorous physical activity; SMD, standardized mean difference.

### Correlation between changes in CBF, cognitive function, fitness, body composition, MVPA and muscle-strengthening sessions

The changes in CBF (both global gray matter and region-specific) were not significantly correlated with changes in cognitive function domains, fitness indicators, MVPA, muscle-strengthening sessions or body composition (Figure S2).

### Adverse events

No death or life-threatening event occurred, and no MACE was recorded in the usual care group or in the exercise group during a total of 1995 sessions of 45 min each (1496 hours of training). There were 3 serious adverse events (grade III out of V) recorded during the study period. These 3 events occurred in the HIIT group, they were CVD related events (i.e. chest pain reported), but only one of them was considered related to the exercise sessions and chest discomfort had started before the training session. There were three additional participants from the HIIT+RT arm that experienced exercise-related dizziness and hypotension at the beginning of the exercise program (i.e. weeks 1-3), with no need for medical intervention. Twelve additional non-CVD events (grade I or II) were reported in the two exercise arms and only one of them was reported in the usual care arm. Noteworthy, adverse events attributable to the exercise intervention (i.e. those occur during training sessions or within 3h of training) represented a 44.4% of the events. The complete list of events is reported in Table S11.

## Discussion

The Heart-Brain RCT evaluated the effects of HIIT+RT and HIIT on CBF and other health outcomes in individuals with CAD. These 12-week exercise interventions did not lead to significant changes in the primary outcome, CBF, in the whole sample. However, CBF baseline levels moderated the effect of the intervention, such that among participants with lower CBF at baseline, HIIT was effective at increasing CBF compared to usual care and HIIT+RT. Intervention effects were not detected on regional CBF or cognition. On the other hand, both exercise interventions were effective at improving CRF (VO_2_peak), HIIT+RT was effective for improving muscular fitness, and HIIT significantly reduced BMI. Noteworthy, both exercise programs were well accepted by the participants, as indicated by the high attendance, compliance and enjoyment previously reported.^42^

In line with these results, a prior single-arm study showed that a 6-month cardiac rehabilitation (CR), based on MICT and RT, did not induce significant changes in global CBF in 17 individuals with CAD.^33^ By contrast, they reported significant CBF increments bilaterally in the anterior cingulate, using a voxel-wise analysis.^33^ Notably, they lacked a peer-matched control group, so they were unable to report between-group differences.

Previous results in general aging populations also remain inconclusive. Several studies^79–85^ found no significant changes in CBF in healthy older adults following aerobic exercise interventions lasting from 8 to 26 weeks. Similarly, aerobic exercise did not induce changes in CBF in individuals with vascular cognitive impairment^86^ or in whole brain CBF (including both white matter and gray matter) in individuals with Alzheimer’s Disease.^87^ However, a recent review, which included studies in healthy and clinical populations, found that exercise can effectively increase region-specific CBF, particularly in the hippocampus and anterior cingulate cortex.^88^ Interestingly, behavioral interventions with longer durations may have a greater impact on traditional cardiovascular risk factors, which can translate into enhanced brain vascular health as reflected by CBF increases. A 12-month aerobic exercise intervention in individuals with mild cognitive impairment found increases in CBF accompanied with improved arterial stiffness.^89^ Similarly, 2 diet and aerobic exercise combined interventions in adults with overweight/obesity (1 year duration) and with type 2 diabetes (8 and 11 years duration) found weight loss related to CBF increases.^90,91^ Regarding the weight-loss-related improvements in CBF mentioned above, it is noteworthy that, in our study, the only group that reduced BMI was the HIIT group, and that was also the only one that improved CBF among participants who had lower baseline CBF. Moreover, in our sample, BMI was inversely correlated with CBF at baseline,^92^ which together with the previous studies cited above, could point to a potential mediating role of BMI. However, we failed to demonstrate support for this mediation hypothesis, since no correlation was observed between changes in BMI and changes in CBF, and formal mediation models (supplemental methods), with changes in BMI as potential mediator, were not significant (data not shown). Furthermore, the HIIT group had a larger increase than the HIIT+RT in weekly MVPA (124 vs. 40 min/week, compared to the usual care group) as objectively assessed by accelerometers, which is known to have multiple benefits^26^ and could have potentially contributed to the larger positive effects observed on both BMI and CBF.

Importantly, the pathways through which regular exercise training may induce CBF changes are yet to be fully understood. CBF is affected and controlled by multiple factors (e.g. cardiac output, endothelial function, intracranial pressure, metabolism, neural activity, blood gases, and neurovascular innervation), which are likely involved in mechanisms activated by exercise and could potentially lead to differential effects depending on exercise intensity.^93,94^ However, as prior interventions were mostly based on MICT,^80,81,83–85^ it is difficult to understand how different training methods may differently affect CBF by inducing distinct vascular mechanisms. One recent study,^79^ which used a 26-week HIIT+RT intervention in older adults, found significant CBF reductions in participants that exhibited larger gains in CRF following the intervention. The opposite was found by Maass et al.^82^, who reported an increase of CBF in the hippocampus associated with improved CRF (ventilatory anaerobic threshold) in healthy older adults after a 3-month aerobic exercise intervention. In our study, we similarly explored whether CBF changes were correlated to cognition, VO_2_peak and 30s sit-to-stand performance changes, but no significant correlations were observed.

What seems more conclusive is that CBF decreases with age and differs between sex across the lifespan.^95–97^ A previous study^82^ showed that, whereas exercise elicited CBF increases in individuals aged between 60 and 70 years, it tended towards decreases in older participants. It is noteworthy that, in our study, both exercise groups showed increases in CBF in individuals aged below 65 years and decreases in those aged 65 or above, yet these effects and interaction were not significant. Besides, a recent RCT in older adults showed that those with higher subjective cognitive decline and larger self-reported memory complaints had larger improvements in cognitive outcomes after a 24-week RT intervention.^98^ These findings suggest that participants with poorer baseline status have more room to improve and may therefore rapidly benefit from behavioral interventions. Likewise, we observed that CBF increased significantly only in participants with lower baseline levels after the HIIT intervention. Nevertheless, these results should be interpreted with caution and contrasted in future studies with larger sample size, more sufficiently powered for subgroup analyses.

Focusing on cognitive function, exercise has been demonstrated to elicit improvements in general cognitive performance as well as in specific cognitive domains, with low and moderate-intensity interventions showing larger effects.^99^ For individuals with CVD, exercise-based CR has consistently been found to improve global and attention-psychomotor cognitive functions, whereas the effect on executive function and memory remains unclear.^100^ To our knowledge, there is only one previous study that examined the effects of exercise on cognitive function in 18 individuals with CAD, which reported improvements in psychomotor speed and complex attention after a 12-week CR program.^101^ In our study, we did not find between-group effects in general cognition or any of the cognitive domains studied. However, findings from CR based studies are difficult to compare with ours as they do not have a control group.^100,101^

On the other hand, our findings are consistent with results from previous research, which demonstrated that both HIIT^102–105^ and combined aerobic exercise with RT^106,107^ are effective for increasing VO_2_peak in patients with CAD. We observed improvements of 2.61 mL/kg/min for the HIIT+RT and 2.53 mL/kg/min for the HIIT, when comparing with usual care, an effect size considered clinically relevant in previous RCTs in a CVD population.^75^ These results indicate that condensing traditional 4x4 HIIT interval into 3x4 intervals plus a RT circuit, in line with the recent position stand on RT prescription from the American College of Sports Medicine,^31^ can elicit similar improvements in VO_2_peak, while meeting the WHO recommendations^26^ and the American and European clinical guidelines for CAD management.^27–30^ Furthermore, we compared the effects of two different exercise trainings on the effect of 30s sit-to-stand performance. This is a widely used method to assess muscular fitness (muscle strength and endurance), functional capacity (crucial for daily life), indirect risk of falls, fractures and frailty, balance, coordination and cardiorespiratory fitness.^65^ We detected that HIIT+RT significantly improved muscle fitness (vs usual care: 3.32 repetitions; vs HIIT: 3.10 repetitions), making it a more comprehensive and time-efficient approach for improving physical function, CRF and muscular fitness, in individuals with CAD. Remarkably, these effect sizes are larger than the minimum clinically important difference reported for the 30s sit-to-stand test (i.e., 2 repetitions).^108^

The two types of HIIT interventions investigated in this RCT can be considered safe, as no MACE, life threatening event or death occurred in roughly 2000 supervised exercise sessions (≈1500 hours of training). It should be emphasized that we strictly followed the guidelines to implement HIIT in clinical populations,^43^ the list of contraindications and a cardiologist-supervised incremental maximal test was conducted at baseline excluding participants when ineligible for high intensity exercise. This cautious approach reduced the probability of major adverse events and death in this trial and should also be considered in future studies or clinical practice. Nevertheless, some serious (severity grade III out of V) events occurred during the study. Three participants in the HIIT group reported chest pain during the study, yet two of them occurred outside the window of the exercise sessions. Of note, these three participants had a history of repeated chest pain symptoms before enrolling in the study. It was therefore expected that similar symptoms could arise during the study. Indeed, it could be argued that exercise at high intensity could reveal (not cause) supply-demand related ischemia in individuals with CAD. Additionally, three exercise-related hypotension events occurred in the HIIT+RT group, in three different individuals, during the RT section of the session. Eighty percent of the sample were taking anti-hypertensive drugs, potentially affecting the hemodynamic ability to quickly adjust to the change of positions (some including exercises on the floor immediately followed by upright standing^42^). Interestingly, these three events occurred in the very beginning of the exercise program (i.e. first training session, week 2 and 3), all the participants continued with the program seamlessly, suggesting an adaptation to this type of effort and change of positions. In addition, participants were advised to properly hydrate before and during the training sessions, and were instructed to breathe continuously during resistance exercise, avoiding the Valsalva maneuver, which together can reduce these types of events. Finally, other milder events were reported by the participants during the study, mostly from the two exercise groups. We recorded 11 events like headaches and pain, infections, COVID-19 in the exercise groups, whereas only 1 event of this type was reported by the usual care group. It is highly probable that the intense interaction between the participants in the exercise groups and the intervention staff during the 3 sessions/week (1:1 ratio trainer-participant) for 12 weeks led to more communication and event reporting than in the usual care group.

### Limitations and Strengths

The major strength of this study is addressing the knowledge gap by testing the effect of exercise on CBF and cognitive function in individuals with CAD, which is of upmost importance given their elevated cognitive burden and dementia risk, and limited prior investigation (see our meta-analysis^23^). Besides, we analyzed other relevant health outcomes, such as CRF, muscular fitness and body composition. For the measurement of the primary outcome, we employed a state-of-the-art, accurate, and reproducible non-invasive technique.^109^ Moreover, the use of a comprenhensive battery of cognitive tests allowed us to examine different cognitive domains at a greater depth than prior studies. Further, we have tested two different exercise programs, the well-known 4x4 HIIT, and a variation of it, that included 2 rounds of 8 RT exercise. This new exercise model was able to improve CRF and muscular/functional fitness in a time-efficient manner. Additionally, we have assesed changes in accelerometer based MVPA and self-reported muscle-strengthening sessions, to control for potential contamination and compensatory effects.

Nevertheless, several limitations were present. First, some conditions that may have affected CBF (time of the MRI, medication, fasting status or exercising 24 hours before the scan, although no intervention session was performed the previous day) were not controlled before the MRI scan. Second, the largest post-labelling delay, defined as the time between blood labelling and the image acquisition, in the ASL sequence was 2 seconds, potentially not capturing the signal from those regions where the blood takes longer to arrive. A 3-month length intervention might not have been long enough for inducing brain health changes in this study, and a larger sample size would have allowed for more powered subgroup analyses.

## Conclusion

This RCT showed that a 12-week HIIT+RT and HIIT intervention did not increase CBF in a sample of individuals with CAD, but HIIT led to significant CBF improvement only in the subgroup with lower baseline CBF (predefined analysis), i.e. those who potentially need it the most. Interestingly, only the HIIT program was effective on eliciting BMI reductions, and previous literature has shown that weight-loss might play a role in CBF improvements. No intervention effects were observed on regional CBF or cognitive performance. On the other hand, both HIIT+RT and HIIT improved VO_2_peak, and only HIIT+RT additionally enhanced muscular fitness, making it a more complete option to induce clinically significant improvements in cardiorespiratory and muscular fitness in individuals with CAD in a time-efficient manner (3 sessions/week of 45min). The cognitive burden and the elevated risk of dementia in CAD population warrant further investigation, specifically focusing on individuals with lower CBF, as well as longer interventions.

## Acknowledgments

We thank Dr. Veronica Cabanas-Sanchez for performing the randomization of the study. The authors are grateful to the participants for taking part in this study. Additionally, we acknowledge Kai Valiant de Geus for his contribution to the visual quality control of pCASL images.

## Data sharing statement

Metadata (e.g. study protocols, data dictionaries, data processing scripts, statistical analyses plan) of the project will be open access available when the final script is ready after the peer-review process. All metadata will be hosted in the on a GitHub repository for version control and ease of reuse (https://github.com/Heart-Brain/Heart-Brain) and archived on Zenodo (https://doi.org/10.5281/zenodo.17865709) to ensure long-term preservation and to provide digital object identifiers (DOIs). The individual patient data of the Heart-Brain trial will not be made open access, due to privacy concerns and violation of the General Data Protection Regulation (due to the low number of cases in certain specific characteristics). Individual data will be available under restricted access following the "as open as possible, as closed as necessary" principle. The data files will include pseudonymized identification codes and only contain participants who provided informed consent for data sharing. The procedure for data sharing can be requested via the PI of the study (Prof. Ortega). Following this procedure, this study fully complies with the Open Science principles including FAIR data management, reproducibility, and inclusive, collaborative practices.

## Supplemental methods

### 1. Consolidated Standards of Reporting Trials adherence

**Table S1.**
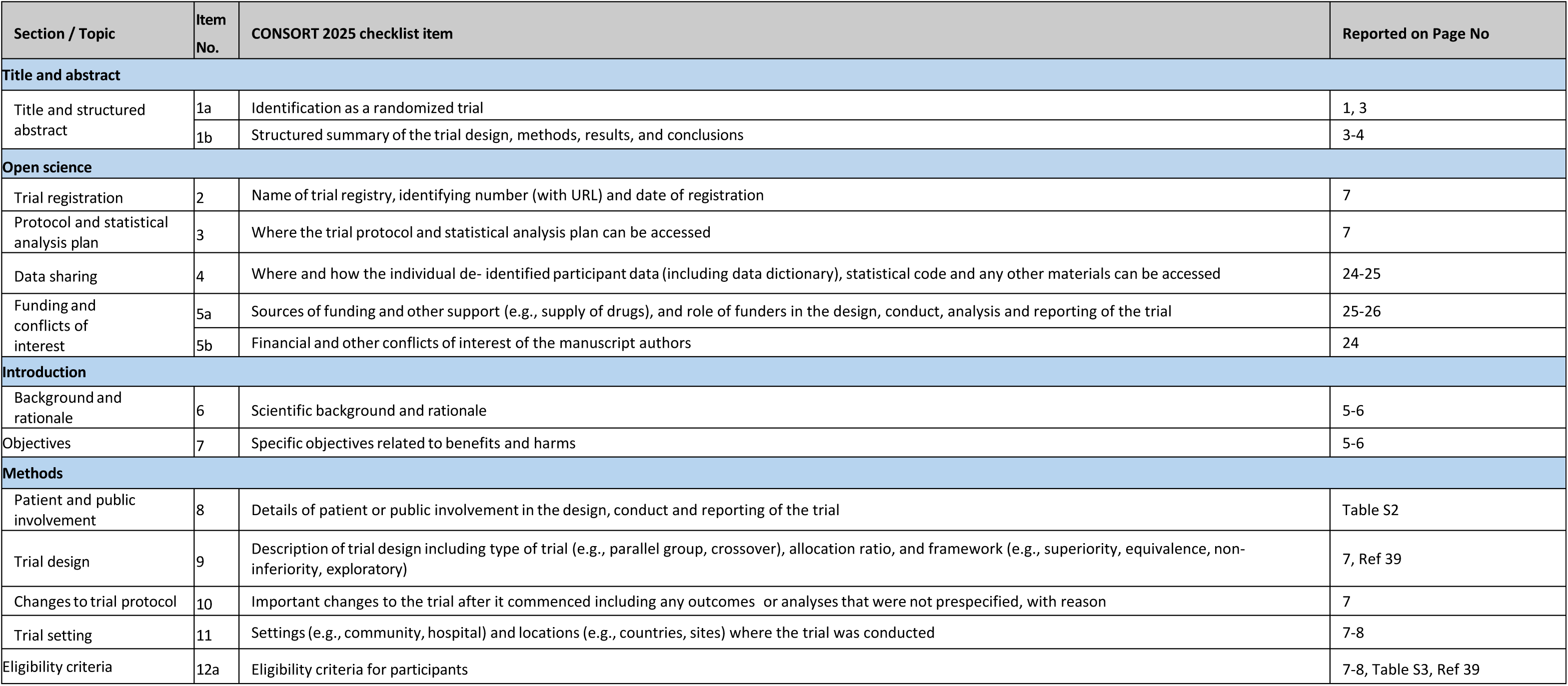
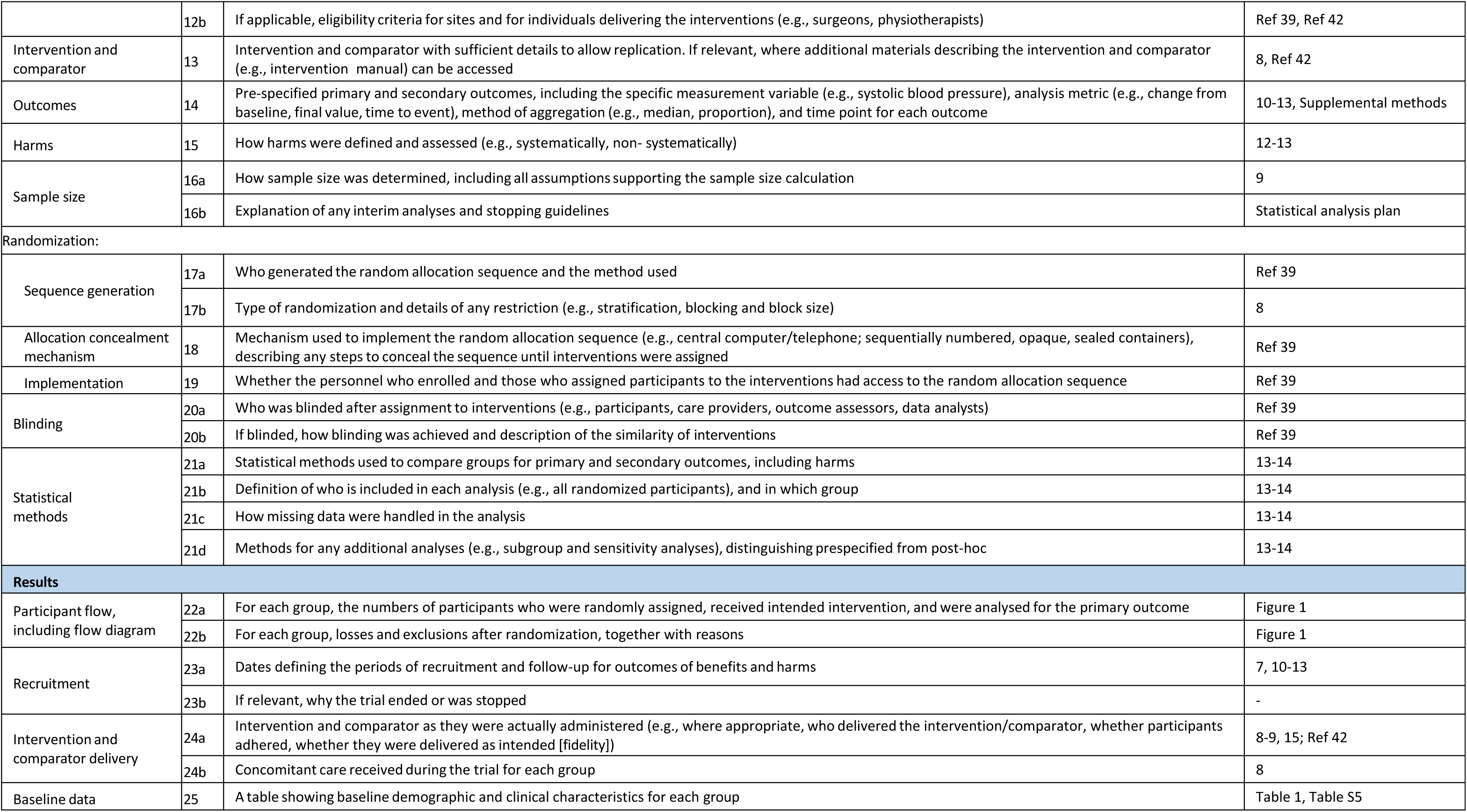
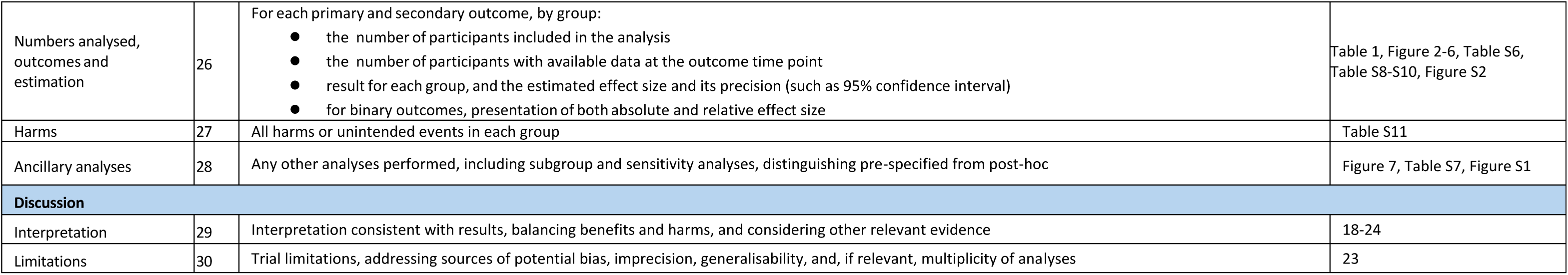
CONSORT (Consolidated Standards of Reporting Trials) checklist.

### 2. Patient and public involvement

**Table S2.** Patient and public involvement actions taken, split by phase of the study and groups of people involved.

| Phase of the study | Group involved | Action description |
| --- | --- | --- |
| Design | Patients' representatives | The principal investigator (FBO) visited the Cardiac Patients' Association of Granada to disseminate information about the study and gather feedback from patients' representatives. |
|  | Cardiologists | A team of cardiologists was actively involved in the planning of the project as part of the research team |
| Implementation | Patients' representatives | The Cardiac Patients' Association of Granada helped to identify and refer potential participants for the screening process. |
|  | Cardiologists | Cardiologists took part in the evaluations, including echocardiograms and cardiopulmonary exercise test, and were involved in clinical decision-making in the case of adverse events. |
|  | Research team and participants | A final evaluation questionnaire was administered to all participants to gather their feedback on the different phases of their participation in the study. |
| Dissemination | Research team and participants | At the end of the study, participants received a personalized report with some health-related outcomes at post-assessments, and a brochure from the Cardiac Patients' Association containing information about the associations' aims and actions. |
|  | Research team | A comprehensive description of the exercise interventions developed in the Heart-Brain trial has been published in open access.<sup>42</sup> In addition, detailed multilingual manuals and instructional videos have been uploaded to YouTube/Zenodo/Github, providing accessible resources for public health professionals, patients and general public interested in implementing a feasible, low-cost HIIT and resistance-based exercise program. |

### 3. Inclusion and exclusion criteria

**Table S3.**
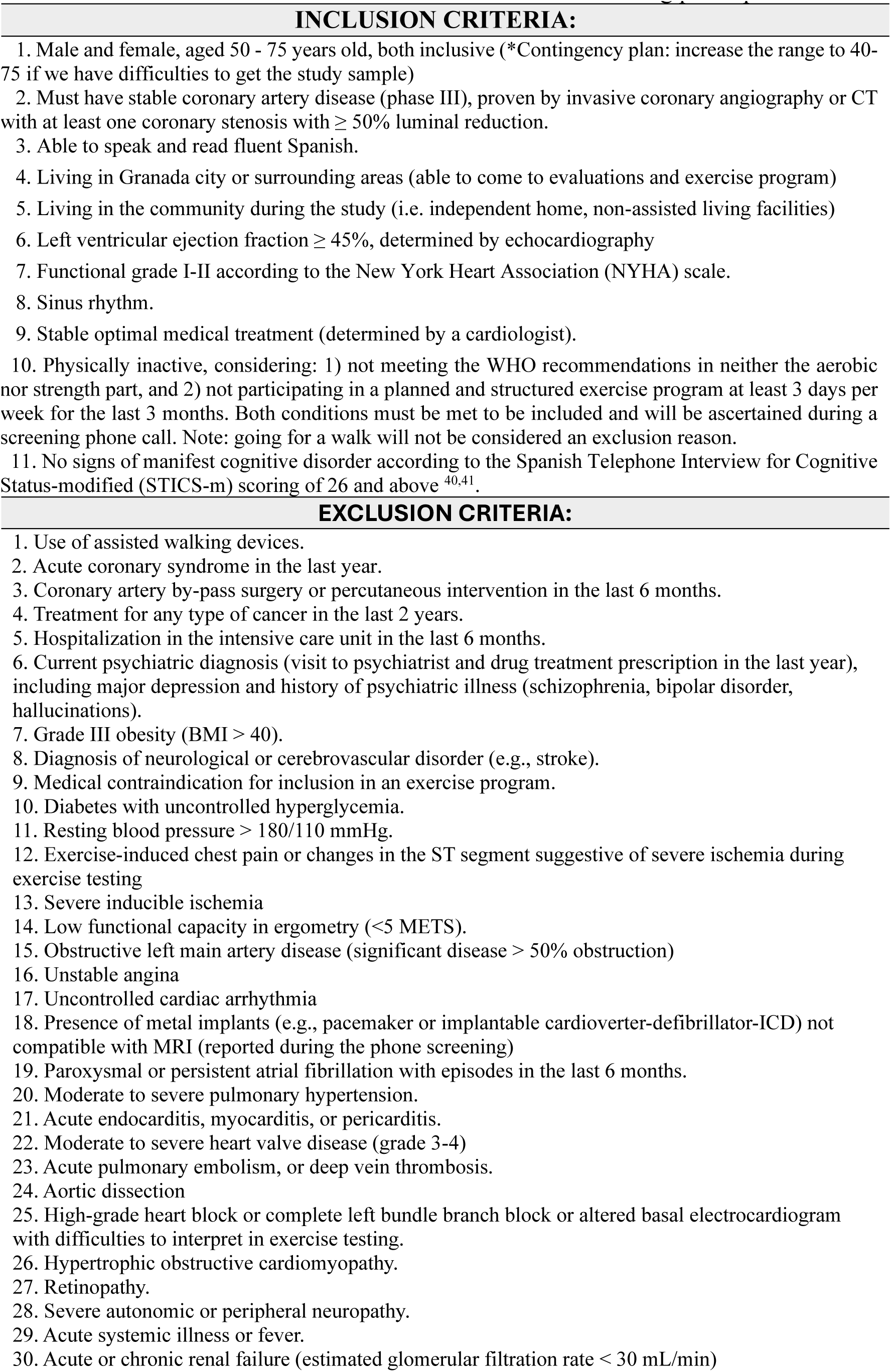
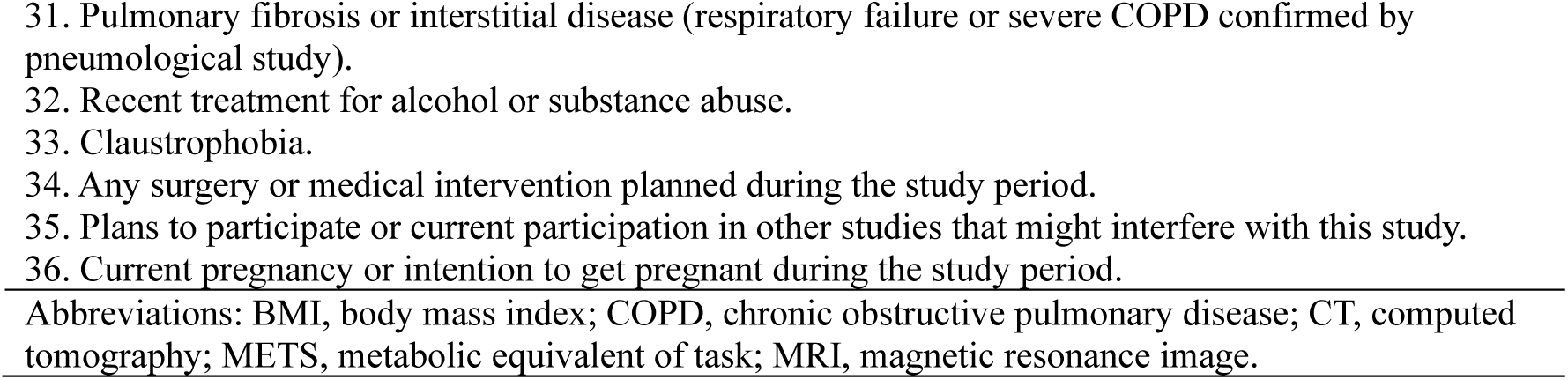
Heart-Brain inclusion and exclusion criteria for selecting participants.

### 4. MRI

#### 4.1. MRI acquisition

Background-suppressed four-delay pCASL, including a proton density calibration image (M0) and 9 label-control pairs, based on the protocol by Wang et al.,^46^ was performed to obtain cerebral blood flow (CBF, hereafter refers to gray matter only unless otherwise indicated) measurements. Participants were advised to remain awake during the scan. Sequence parameters were: labeling duration= 1500 ms, post-labeling delays: 2x500, 2x1000, 2x1500, 3x2000 ms, TR= 4300ms, TE= 23.64 ms, flip angle= 120, FOV= 300x300 mm, slice thickness= 2.5 mm, voxel resolution= 3.1×3.1×2.5 mm.

A sagittal 3D T1 weighted image (TR = 2400, TE = 2.31, number of slices = 224, voxel resolution = 0.8x0.8x0.8 mm) was acquired and processed for registration and segmentation purposes.

#### 4.2. MRI processing

T1 weighted scans were preprocessed using FSL’s ANAT pipeline,^47^ which includes reorientation, cropping, bias-field correction, registration to standard space, brain extraction, tissue segmentation, and subcortical structure segmentation.

Then, two stepwise coregistration was performed to align the T1 weighted image and arterial spin labelling (ASL) M0. *Mri_coreg* from Freesurfer (version 7.4.1)^48^ performed a linear registration between the two volumes. The resultant transformation matrix was then used as initial transformation matrix for running FSL’s FLIRT^49–51^ boundary-based coregistration, with six degrees of freedom rigid body transformation.

Finally, ASL to structural transformation matrix coming from FSL’s FLIRT was entered into *oxford_asl* command, together with ASL data and FSL’s ANAT outputs to get CBF maps. *Oxford_asl* uses the FSL’s FABBER ASL package and Bayesian Inference to invert the kinetic model for ASL MRI (BASIL).^52,53^ Previously described sequence parameters (post-labeling delays, TR and TE) and labeling efficiency (set at 0.7) were specified as input to the kinetic models for estimating CBF. Additional input parameters were kept with default settings corresponding to pCASL acquisition. Given tissue segmentations [gray matter (GM), white matter (WM) and cerebrospinal fluid (CSF)], BASIL calculates CBF in GM and WM. Partial volume correction was used to report CBF.^54^

##### 4.2.1.Regional CBF analyses

For the calculation of regional CBF, FreeSurfer’s brain image was coregistered to the ASL image using *epi_reg,*^110^ and the resulting transformation matrix was applied to the aparc+aseg segmentation image. Then, cortical and subcortical gray matter regions were extracted with *fslmaths* command, and the mean CBF for each region of interest (ROI) was calculated separately for each hemisphere by averaging CBF values of all voxels within the ROI using the *fslstats* command. The parcellation included the following gray matter brain areas for both the left and right hemisphere: banks of the superior temporal sulcus, caudal anterior cingulate cortex, caudal middle frontal gyrus, cuneus, entorhinal cortex, fusiform gyrus, inferior parietal lobule, inferior temporal gyrus, isthmus of the cingulate gyrus, lateral occipital cortex, lateral orbitofrontal cortex, lingual gyrus, medial orbitofrontal cortex, middle temporal gyrus, paracentral lobule, parahippocampal gyrus, pars opercularis, pars orbitalis, pars triangularis, pericalcarine cortex, postcentral gyrus, posterior cingulate cortex, precentral gyrus, precuneus, rostral anterior cingulate cortex, rostral middle frontal gyrus, superior frontal gyrus, superior parietal lobule, superior temporal gyrus, supramarginal gyrus, temporal pole, transverse temporal gyrus, insular cortex, frontal pole, thalamus proper, caudate nucleus, putamen, globus pallidus, hippocampus, amygdala, nucleus accumbens, and cerebellar cortex. Anterior cingulate cortex CBF was calculated as the mean of caudal anterior cingulate and rostral anterior cingulate CBF. Due to high correlation between hemispheres (r Pearson correlation values between 0.8 and 0.9, p<0.001) for the regions (hippocampus, posterior cingulate cortex, anterior cingulate cortex and precuneus) included in this study, values from both hemispheres were later averaged.

##### 4.2.2 Whole brain CBF analysis

To perform the whole-brain analyses, perfusion maps transformed to the Montreal National Imaging (MNI) space, coming from the BASIL processing,^52,53^ were used. Then, MNI perfusion maps from both time-points were subtracted to get the difference between baseline and post-assessment (i.e. post-assessment MNI CBF map – baseline MNI CBF map), and the individuals’ subtracted maps were smoothed using a Gaussian filter with a Full Width at Half Maximum (FWHM) of 6 mm. The smoothed subtracted maps were introduced into randomise command from FSL.^111^

#### 4.3. Visual inspection and quality control

Mean native space difference maps for each participant were visually inspected,^112,113^ by two independent raters (LSA and KdG), to ensure data quality. Attention was paid to ensure there were no: i) excessive motion resulting in spurious effects in difference map; ii) maps with brain territories which did not appear to be perfused; iii) maps with perfusion difference between hemispheres; and iv) faulty scans due to wrong placement of the ASL imaging slab. Further quality control procedures included the calculation of CBF spatial coefficient of variation (CoV), as previously described by Mutsaerts et al.^114^ CBF maps were ranked into 3 levels according to their CBF CoV: ‘good’ (CoV < 0.6, CBF flow signal predominates artefacts); (ii) ‘acceptable’ (0.6 ≤ CoV ≤ 0.8, both CBF signal and artefacts visible); or (iii) ‘bad’ (CoV > 0.8, artefacts predominate CBF signal). CBF maps were later reviewed, and CoV-based categorization of the images were corrected if necessary. Mean native space difference maps that met at least one of the previously defined criteria, and CBF maps ranked with CoV acceptable or bad, were excluded from the analyses.

### 4. Cognitive outcomes

#### ***4.1.*** Description of the cognitive tasks

Participants completed a comprehensive neuropsychological evaluation using both paper-based and computer-based assessments. All computerized cognitive tests were administered via PC or iPAD with a standard monitor and keyboard. A full description of each is provided below.

##### 4.1.1 Montreal Cognitive Assessment

The Montreal Cognitive Assessment (MoCA) is a widely used and validated screening instrument for detecting cognitive impairment.^56^ The MoCA involves brief assessments of short-term memory, visuospatial abilities, orientation, attention, language, and executive functions, each of which is scored separately. These scores can be summed to calculate a total score or used as individual scores for each section. Participants can receive up to 30 points on the MoCA. The Clock Drawing Test is part of the MoCA and assesses visuospatial and planning abilities. Participants are instructed to draw a clock, including placing the hands to represent a specific time (e.g., 11:10).

##### 4.1.2 Trail Making Test

The Trail Making Test (TMT) is a paper-and-pencil test that measures cognitive flexibility and set-shifting.^59^ TMT Part A is a measure of psychomotor processing speed that requires participants to connect numbers displayed on a page in ascending order. TMT Part B measures set-shifting and requires participants to alternate between numbers and letters to connect them in ascending order. Time to completion (seconds) was measured, and reverted (multiplied by -1) with higher values reflecting better performance.

##### 4.1.3 Digit Symbol Substitution Test

The Digit Symbol Substitution Test (DSST) is a paper-and-pencil test designed to assess psychomotor processing speed and basic attention.^58^ The DSST requires copying as many novel symbols corresponding to numbers as possible in 120 seconds. Total number of correct responses was measured. Higher scores reflect better cognitive performance.

##### 4.1.4 Picture Sequence Memory Test

Picture Sequence Memory Test (PSMT) is a computer-based task from NIH Toolbox Cognition Battery (version 1.17), that evaluates episodic memory.^57^ Participants were presented with a series of pictures in a specific order, and their task was to remember the order in which the pictures were shown. The number of pictures the participants were required to order ranged from 15-picture sequences to 18-picture sequences. Raw scores reflect the cumulative number of adjacent pairs of pictures remembered correctly over the three trials. Higher scores reflect better cognitive performance.

##### 4.1.5 Dimensional Change Card Sort Test

Dimensional Change Card Sort Test (DCCST) is a computer-based task from NIH Toolbox Cognition Battery (version 1.17) which assesses cognitive flexibility and the ability to shift attention between different dimensions or rules.^57^ Participants were trained on one rule and then required to shift their attention and adapt to a new sorting rule during the shift phase. In test trials, participants sort cards based on the current rule presented on the screen, which can switch between shape and color. Accuracy and response times were recorded (e.g., higher scores reflect better cognitive performance).

##### 4.1.6 List Sorting Working Memory Test

List Sorting Working Memory (LSWMT) is a computer-based task from NIH Toolbox Cognition Battery (version 1.17) that measures working memory.^60^ Participants were asked to recall and sequence by size different stimuli that were presented visually and via audio. If the participant sorted the list correctly, another longer list of familiar stimuli (foods and/or animals) was presented. On 1-list trials, all stimuli were from the same category (foods or animals). On 2-list trials, the stimuli were from two different categories (foods and animals). The participant was asked to sequence the food stimuli followed by the animal stimuli. Total items correct across the 1-list and 2-list conditions (maximum 28) were recorded.

##### 4.1.7 Flanker test

The Flanker test is an arrow version of the computer-based task from NIH Toolbox Cognition Battery (version 1.17) that measures inhibition.^57^ During this task participants are presented with a series of five arrows presented in a row on the screen at the same time, each pointing left or right. Participants had to respond as quickly as possible the direction of the middle arrow (right or left, using corresponding arrows on the keyboard), ignoring the external arrows. Accuracy and response times were recorded (e.g., higher scores reflect better cognitive performance). Higher scores reflect better cognitive performance.

##### 4.1.8 Spatial Working Memory Test

Spatial Working Memory Test (SWMT) is a computer-based task, aimed to assess an individual’s ability to temporarily store and manipulate spatial information.^61^ The participants were asked to stare at a crosshair in the middle of the screen. Then, 2, 3 or 4 black dots appear at random locations on the screen and disappear afterwards. Then, a red dot appears on the screen, and the participant must respond if the dot is either placed in the same place as one of the black dots that appeared before, or not, as fast as possible. Accuracy and the time taken to respond were recorded. Higher scores reflect better cognitive performance.

#### ***4.2.*** Data processing

Z scores were calculated for general cognition and the four subdomains included in the study, to be used in the analyses. For both, baseline and post-assessment, z-scores were calculated by subtracting the mean and dividing by the standard deviation (SD) from baseline, to standardize effect sizes. When needed, the original indicator scoring was inverted so that a higher score means higher cognitive performance in all cognitive outcomes.

General cognition was evaluated with Montreal Cognitive Assessment (MoCA) test. Cognitive performance from each test were grouped into four different subdomains (Table S1).

**Table S4.**
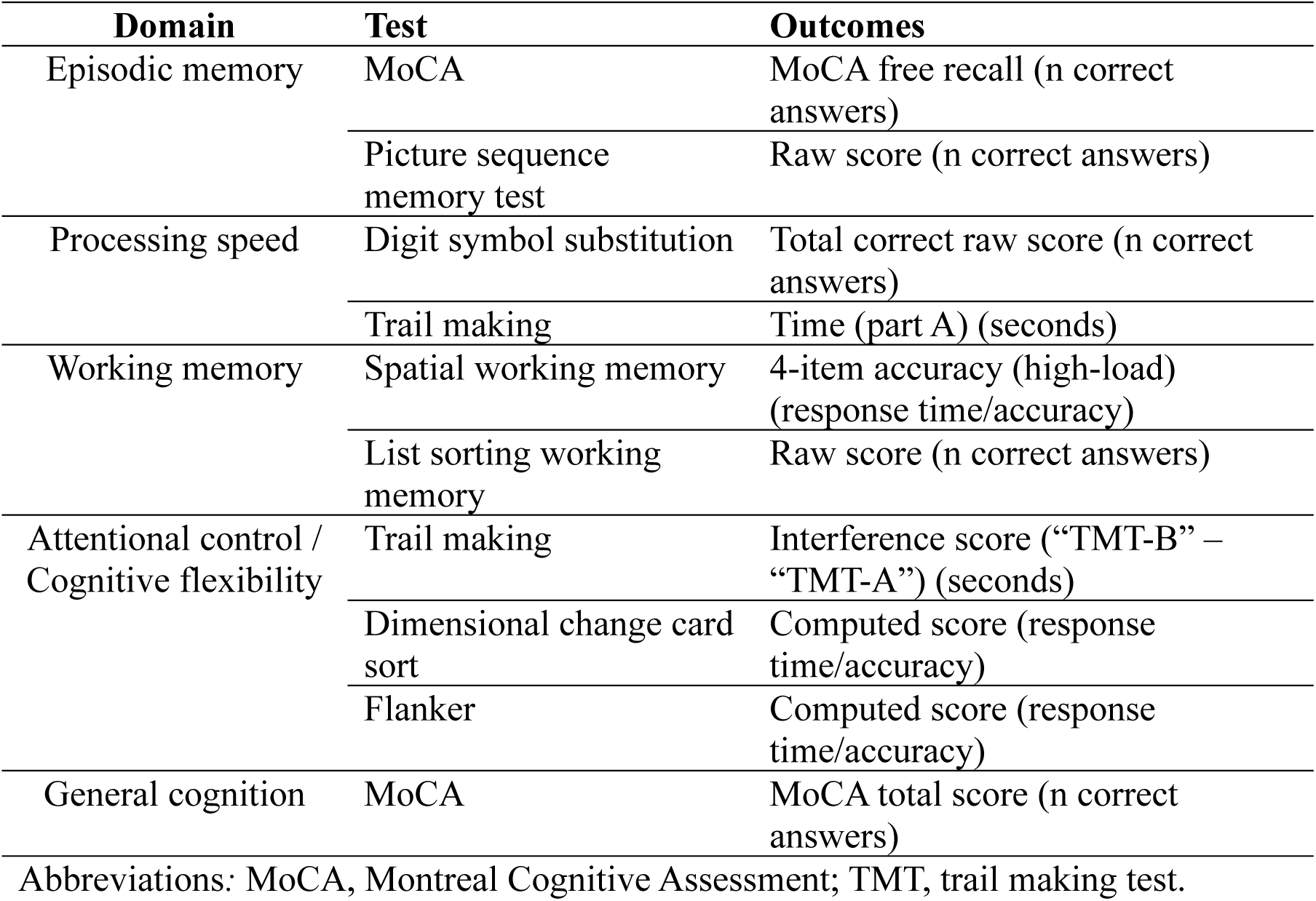
Neuropsychological tests and outcome variables in each cognitive domain.

| Domain | Test | Outcomes |
| --- | --- | --- |
| Episodic memory | MoCA | MoCA free recall (n correct answers) |
|  | Picture sequence memory test | Raw score (n correct answers) |
| Processing speed | Digit symbol substitution | Total correct raw score (n correct answers) |
|  | Trail making | Time (part A) (seconds) |
| Working memory | Spatial working memory | 4-item accuracy (high-load) (response time/accuracy) |
|  | List sorting working memory | Raw score (n correct answers) |
| Attentional control / Cognitive flexibility | Trail making | Interference score (“TMT-B” – “TMT-A”) (seconds) |
|  | Dimensional change card sort | Computed score (response time/accuracy) |
|  | Flanker | Computed score (response time/accuracy) |
| General cognition | MoCA | MoCA total score (n correct answers) |

Abbreviations: MoCA, Montreal Cognitive Assessment; TMT, trail making test.

### 5. CPET protocol

A standardized incremental CPET on a treadmill (h/p/cosmos, Nussdorf, Germany), according to the American College of Sports Medicine (ACSM) guidelines,^62^ was included in both pre- and post-assessments, and served as a tool for both clinical evaluation and performance measurement. The test had the following structure: (i) a 3-minute warm-up with progressively increasing speed until 4.8 km/h; (ii) a main part, characterized by 1% increase of the inclination every 30 s until volitional exhaustion or other indication for terminating the test according to guidelines; and (iii) a standardized 90-s active recovery at 0% inclination and gradually reduced speed. The maximal inclination of the treadmill was 25% and thus, the maximal exercise time including warm-up was 15.5 min. Respiratory gases were registered with a dilution flow system (Omnical, Maastricht Instruments, Maastricht, the Netherlands), generating output data with a time resolution of 5 s. A 12-lead electrocardiogram was continuously registered (AMEDTEC ECGpro, GmbH, Aue, Germany) and monitored by a cardiologist throughout the test. In addition, second by second heart rate (HR) was also monitored with a chest strap connected to a sports watch (Polar H10, Kempele, Finland). Blood pressure (BP) was measured during the test every 3 minutes with an automatic device using a microphone over the brachial artery for sound detection and R-wave gating from the ECG (Tango® M2 ECG-gated Automated BP Monitor, Suntech Medical Inc, NC, USA). Every 3 minutes, the participant was also asked to indicate the rate of perceived exertion (RPE) on a visual 0-10 scale.

#### 5.1 Data processing

Respiratory gases data (sampling rate of 5 seconds) and ECG data (sampling rate of 30 seconds) were matched according to registered exercise time. After visual assessment of each test, dilution flow system and ECG data were adjusted manually to match at the end of exercise. Then, Polar data for each test was resampled to 5 second averages and matched to the other test data by exercise duration. Each test was individually examined visually to verify the validity of variables at the end of exercise. Non valid variables due to for example leaking of gas at the end of exercise [falling ventilation (VE), oxygen consumption (VO_2_) and carbon dioxide production (VCO_2_) values], were noted in separate columns for each variable to be used in the later analyses where only valid test data should be included. Furthermore, to avoid respiratory gases resting values from the short acclimatization phase, we applied an arbitrary VO_2_ cutoff of 0.35 L/min. In case VO_2_ values were lower than the cutoff, VO_2_, VCO_2_ and respiratory exchange ratio (RER) data were excluded. Any negative values were also removed.

All data processing was performed using R (version 4.3.1).

#### 5.2 VO_2_peak determination

VO_2_peak, expressed in L/min, was calculated as the highest 30 second average of absolute oxygen uptake during the test. As there may be a slight time delay from VO_2_peak to registration of peak values due to intrinsic inertia in the gas registration mixing chamber type system, and as there may be a matching error of +/- 10 seconds in the timing between dilution flow system data and ECG test data (the latter indicating end of exercise), the 30 sec rolling average was not restricted only to the exercise phase, but could be extended to early recovery if needed. To express VO₂peak in relative terms, values were normalized to body mass by dividing by each participant’s body weight, yielding VO₂peak in mL/kg/min.

### 6. Moderation outcomes

#### 6.1. Moderators criteria

To evaluate the moderating effects of individual characteristics, participants were categorized based on various key variables. The following sections outline the criteria and methods used to define each category.

##### 6.1.1 Age

Age was dichotomized according to the median sample age as youngers (< 65 years) or elders (≥ 65 years).

##### 6.1.2 Sex

Sex was categorized as male or female based on self-reported information.

##### 6.1.3 Education level

Education was grouped in two levels: 1 (primary studies and secondary studies), 2 (university studies).

##### 6.1.4 Baseline levels of the outcomes studied

Participants were categorized into two groups: low (<median) and high (≥median), based on the median from values assessed at baseline.

### 7. Exploratory statistical analyses

Mediation analyses following AGReMA (A Guideline for Reporting Mediation Analyses) were conducted using the lme4 and bruceR packages in R. Database was split accordingly for each of the comparisons (i.e. HIIT+RT vs usual care, HIIT+RT vs usual care, HIIT vs HIIT+RT) to facilitate interpretation. BMI and VO2peak were introduced as potential mediators of the effects of exercise on CBF, and the intervention group was entered as predictor. We included baseline CBF as a covariate.

## Supplemental results

**Table S5.** Baseline characteristics of the Heart-Brain sample split by non-dropout and dropout participants.

| Characteristic | Non-dropouts N = 94<sup>I</sup> | Dropouts N = 11<sup>I</sup> | p-value<sup>2</sup> |
| --- | --- | --- | --- |
| Age | 62.22 (6.65) | 61.34 (6.27) | 0.7 |
| Sex |  |  | 0.3 |
| Male | 76 (81%) | 7 (64%) |  |
| Female | 18 (19%) | 4 (36%) |  |
| Education level |  |  | >0.9 |
| Primary and secondary school | 56 (60%) | 7 (64%) |  |
| University | 38 (40%) | 4 (36%) |  |
| Weight (kg) | 82.72 (12.79) | 86.97 (16.13) | 0.4 |
| Height (cm) | 168.73 (8.23) | 170.13 (13.31) | 0.7 |
| Body mass index (kg/m<sup>2</sup>) | 29.02 (3.81) | 29.99 (4.34) | 0.5 |
| BMI category |  |  | 0.7 |
| Normal weight | 8 (8.5%) | 1 (9.1%) |  |
| Overweight | 54 (57%) | 5 (45%) |  |
| Obesity | 32 (34%) | 5 (45%) |  |
| Muscle mass (kg) | 55.43 (8.11) | 56.37 (11.93) | 0.8 |
| Fat mass (kg) | 23.73 (6.51) | 27.51 (8.46) | 0.2 |
| Muscle mass index (kg/m<sup>2</sup>) | 19.42 (1.78) | 19.25 (2.00) | 0.8 |
| Fat mass index (kg/m<sup>2</sup>) | 8.40 (2.48) | 9.68 (3.50) | 0.3 |
| Systolic blood pressure (mmHg) | 126.26 (14.05) | 131.13 (17.62) | 0.4 |
| Diastolic blood pressure (mmHg) | 78.11 (8.93) | 80.49 (10.00) | 0.5 |
| Anti-hypertensive treatment | 76 (81%) | 8 (73%) | 0.8 |
| Lipid-lowering treatment | 93 (99%) | 11 (100%) | >0.9 |
| Betablockers treatment | 72 (77%) | 8 (73%) | >0.9 |
| APOE4 carrier | 19 (20%) | 0 (0%) | 0.2 |
| VO<sub>2</sub>peak (mL/kg/min) | 25.32 (5.10) | 24.35 (4.21) | 0.5 |
| 30s Sit-to-stand (repetitions) | 16.93 (4.36) | 15.73 (3.90) | 0.4 |
| Physical activity (minutes/week) | 312.81 (191.72, 447.07) | 421.67 (265.25, 477.02) | 0.4 |
| Strength training (sessions/week) | 0.00 (0.00, 3.00) | 0.00 (0.00, 5.00) | 0.4 |
| CBF (ml/100g/min) | 55.16 (10.66) | 52.68 (8.53) | 0.4 |
| Hippocampus CBF (ml/100g/min) | 43.12 (8.67) | 40.21 (5.65) | 0.2 |
| Precuneus CBF (ml/100g/min) | 48.53 (10.71) | 46.69 (8.85) | 0.6 |
| Posterior cingulate cortex CBF (ml/100g/min) | 57.65 (11.57) | 51.72 (7.45) | 0.044 |
| Anterior cingulate cortex CBF (ml/100g/min) | 47.97 (10.11) | 44.81 (6.08) | 0.2 |
| General cognition (MoCA) score | 24.91 (3.23) | 23.73 (3.41) | 0.3 |
| Episodic memory (z-score) | -0.03 (1.00) | 0.27 (1.02) | 0.4 |
| Processing speed (z-score) | -0.01 (1.01) | 0.13 (0.98) | 0.7 |
| Working memory (z-score) | 0.00 (0.98) | 0.01 (1.22) | >0.9 |

| Characteristic | Non-dropouts N = 94<sup>1</sup> | Dropouts N = 11<sup>1</sup> | p-value<sup>2</sup> |
| --- | --- | --- | --- |
| Executive function & attentional control (z-score) | 0.02 (0.99) | -0.21 (1.12) | 0.5 |
| Data is presented as mean (SD) unless otherwise indicated. |  |  |  |
| <sup>1</sup>Due to missing data sample size varies for: Working memory: Non-dropouts = 92, Dropouts = 11; VO<sub>2</sub>peak: Non-dropouts = 93, Dropouts = 10; CBF related variables: Non-dropouts = 74, Dropouts = 10; Moderate-vigorous physical activity: Non-dropouts = 93, Dropouts = 11; Muscle-strengthening: Non-dropouts = 89, Dropouts = 9. |  |  |  |
| <sup>2</sup>Baseline differences between groups were determined by t-student, wilcoxon rank sum test and chi squared tests for continuous and categorical variables, respectively. |  |  |  |
| Abbreviations: APOE, apolipoprotein E; CBF, cerebral blood flow; HIIT, High Intensity Interval Training; MoCA, Montreal Cognitive Assessment; RT, Resistance training; VO<sub>2</sub>peak, peak oxygen uptake |  |  |  |

**Table S6.** Intention-to-treat effects of the 12-week intervention on global and regional CBF.

| Outcome | Intervention | Baseline mean (95% CI) | Post-assessment mean (95% CI) | Change over time; mean (95% CI) | Interaction (ref = Usual care) | P for interaction (ref = Usual care) | Interaction (ref = HIIT) | P for interaction (ref = HIIT) |
| --- | --- | --- | --- | --- | --- | --- | --- | --- |
| CBF (ml/100g/min) | HIIT+RT | 54.71 (52.50; 56.91) | 54.18 (50.79; 57.57) | -0.52 (-4.87; 3.82) | -0.16 (-4.96; 4.65) | 0.95 | -0.32 (-4.94; 4.25) | 0.89 |
|  | HIIT | 54.71 (52.50; 56.91) | 54.51 (50.87; 58.15) | -0.20 (-4.91; 4.52) | 0.17 (-4.88; 5.26) | 0.95 |  |  |
|  | Usual care | 54.71 (52.50; 56.91) | 54.34 (50.38; 58.30) | -0.36 (-5.52; 4.79) |  |  |  |  |
| Hippocampus CBF (ml/100g/min) | HIIT+RT | 42.71 (41.00; 44.43) | 43.00 (40.50; 45.49) | 0.28 (-2.75; 3.31) | 0.32 (-3.07; 3.71) | 0.86 | 0.80 (-2.43; 4.01) | 0.63 |
|  | HIIT | 42.71 (41.00; 44.43) | 42.20 (39.53; 44.86) | -0.52 (-3.82; 2.79) | -0.48 (-4.00; 3.05) | 0.79 |  |  |
|  | Usual care | 42.71 (41.00; 44.43) | 42.68 (39.79; 45.56) | -0.03 (-3.66; 3.59) |  |  |  |  |
| Precuneus CBF (ml/100g/min) | HIIT+RT | 48.02 (45.73; 50.30) | 47.63 (44.28; 50.98) | -0.39 (-4.49; 3.72) | 0.12 (-4.46; 4.71) | 0.96 | 0.42 (-3.96; 4.78) | 0.85 |
|  | HIIT | 48.02 (45.73; 50.30) | 47.20 (43.62; 50.79) | -0.81 (-5.29; 3.66) | -0.30 (-5.09; 4.53) | 0.9 |  |  |
|  | Usual care | 48.02 (45.73; 50.30) | 47.50 (43.62; 51.39) | -0.51 (-5.41; 4.39) |  |  |  |  |
| Posterior cingulate cortex CBF (ml/100g/min) | HIIT+RT | 56.67 (54.22; 59.12) | 57.97 (54.11; 61.82) | 1.30 (-3.76; 6.35) | 0.01 (-5.53; 5.56) | >0.99 | 2.28 (-3.03; 7.57) | 0.4 |
|  | HIIT | 56.67 (54.22; 59.12) | 55.68 (51.53; 59.83) | -0.99 (-6.47; 4.49) | -2.28 (-8.07; 3.56) | 0.44 |  |  |
|  | Usual care | 56.67 (54.22; 59.12) | 57.96 (53.44; 62.48) | 1.29 (-4.69; 7.26) |  |  |  |  |
| Anterior cingulate cortex CBF (ml/100g/min) | HIIT+RT | 47.33 (45.19; 49.46) | 49.39 (45.92; 52.86) | 2.06 (-2.62; 6.75) | -0.03 (-5.11; 5.07) | 0.99 | 2.10 (-2.73; 6.91) | 0.4 |
|  | HIIT | 47.33 (45.19; 49.46) | 47.29 (43.55; 51.03) | -0.04 (-5.09; 5.02) | -2.13 (-7.42; 3.19) | 0.43 |  |  |
|  | Usual care | 47.33 (45.19; 49.46) | 49.42 (45.34; 53.50) | 2.09 (-3.41; 7.60) |  |  |  |  |

Values indicate estimated marginal means at each time point (and 95% Confidence Intervals).
Abbreviations: CBF, Cerebral blood flow; CI, confidence intervals; HIIT, High Intensity Interval Training; HIIT+RT, high intensity interval training + resistance training

**Table S7.** Per-protocol effects of the 12-week intervention on primary and secondary outcomes.

| Outcome | Intervention | Baseline mean (95% CI) | Post-assessment mean (95% CI) | Change over time; mean (95% CI) | Interaction (ref = Usual care) | P for interaction (ref = Usual care) | Interaction (ref = HIIT) | P for interaction (ref = HIIT) |
| --- | --- | --- | --- | --- | --- | --- | --- | --- |
| CBF (ml/100g/min) | HIIT+RT | 54.95 (52.52; 57.38) | 53.76 (50.14; 57.37) | -1.19 (-5.74; 3.35) | -0.83 (-5.69; 4.06) | 0.74 | 0.37 (-4.39; 5.09) | 0.88 |
|  | HIIT | 54.95 (52.52; 57.38) | 53.38 (49.56; 57.20) | -1.57 (-6.40; 3.26) | -1.20 (-6.26; 3.92) | 0.64 |  |  |
|  | Usual care | 54.95 (52.52; 57.38) | 54.59 (50.59; 58.58) | -0.36 (-5.48; 4.76) |  |  |  |  |
| Hippocampus CBF (ml/100g/min) | HIIT+RT | 43.15 (41.25; 45.04) | 42.98 (40.29; 45.66) | -0.17 (-3.38; 3.04) | 0.00 (-3.46; 3.47) | >0.99 | 1.10 (-2.26; 4.45) | 0.53 |
|  | HIIT | 43.15 (41.25; 45.04) | 41.88 (39.06; 44.70) | -1.27 (-4.68; 2.15) | -1.09 (-4.67; 2.49) | 0.55 |  |  |
|  | Usual care | 43.15 (41.25; 45.04) | 42.98 (40.02; 45.93) | -0.17 (-3.81; 3.46) |  |  |  |  |
| Precuneus CBF (ml/100g/min) | HIIT+RT | 48.27 (45.75; 50.78) | 47.00 (43.40; 50.59) | -1.27 (-5.61; 3.07) | -0.72 (-5.40; 3.98) | 0.76 | 0.36 (-4.19; 4.88) | 0.88 |
|  | HIIT | 48.27 (45.75; 50.78) | 46.64 (42.86; 50.42) | -1.63 (-6.24; 2.99) | -1.08 (-5.92; 3.81) | 0.66 |  |  |
|  | Usual care | 48.27 (45.75; 50.78) | 47.72 (43.76; 51.67) | -0.55 (-5.46; 4.36) |  |  |  |  |
| Posterior cingulate cortex CBF (ml/100g/min) | HIIT+RT | 57.66 (54.98; 60.34) | 57.44 (53.38; 61.49) | -0.22 (-5.40; 4.96) | -1.22 (-6.73; 4.32) | 0.67 | 2.41 (-2.99; 7.77) | 0.38 |
|  | HIIT | 57.66 (54.98; 60.34) | 55.03 (50.74; 59.32) | -2.63 (-8.13; 2.87) | -3.63 (-9.37; 2.18) | 0.22 |  |  |
|  | Usual care | 57.66 (54.98; 60.34) | 58.66 (54.16; 63.15) | 1.00 (-4.82; 6.82) |  |  |  |  |
| Anterior cingulate cortex CBF (ml/100g/min) | HIIT+RT | 47.89 (45.53; 50.24) | 48.85 (45.18; 52.53) | 0.97 (-3.87; 5.81) | -1.01 (-6.13; 4.13) | 0.7 | 2.39 (-2.58; 7.34) | 0.35 |
|  | HIIT | 47.89 (45.53; 50.24) | 46.46 (42.57; 50.36) | -1.42 (-6.56; 3.71) | -3.40 (-8.70; 1.94) | 0.21 |  |  |
|  | Usual care | 47.89 (45.53; 50.24) | 49.87 (45.78; 53.95) | 1.98 (-3.44; 7.40) |  |  |  |  |
| General cognition (MoCA) (z-score) | HIIT+RT | 0.04 (-0.15; 0.24) | 0.30 (0.00; 0.59) | 0.25 (-0.12; 0.63) | 0.09 (-0.29; 0.47) | 0.65 | 0.08 (-0.30; 0.47) | 0.67 |
|  | HIIT | 0.04 (-0.15; 0.24) | 0.22 (-0.09; 0.52) | 0.17 (-0.21; 0.56) | 0.01 (-0.38; 0.39) | 0.98 |  |  |
|  | Usual care | 0.04 (-0.15; 0.24) | 0.21 (-0.09; 0.51) | 0.17 (-0.21; 0.54) |  |  |  |  |
| Episodic memory (z-score) | HIIT+RT | 0.01 (-0.20; 0.22) | 0.12 (-0.17; 0.40) | 0.11 (-0.22; 0.44) | -0.20 (-0.54; 0.14) | 0.25 | -0.24 (-0.58; 0.10) | 0.17 |
|  | HIIT | 0.01 (-0.20; 0.22) | 0.36 (0.07; 0.65) | 0.35 (0.01; 0.69) | 0.04 (-0.31; 0.39) | 0.82 |  |  |
|  | Usual care | 0.01 (-0.20; 0.22) | 0.32 (0.03; 0.61) | 0.31 (-0.02; 0.64) |  |  |  |  |
| Processing speed (z-score) | HIIT+RT | -0.00 (-0.20; 0.20) | 0.28 (0.03; 0.52) | 0.28 (0.06; 0.50) | -0.01 (-0.24; 0.22) | 0.94 | 0.25 (0.02; 0.48) | 0.04 |
|  | HIIT | -0.00 (-0.20; 0.20) | 0.02 (-0.22; 0.27) | 0.03 (-0.20; 0.25) | -0.26 (-0.50; -0.03) | 0.03 |  |  |
|  | Usual care | -0.00 (-0.20; 0.20) | 0.28 (0.04; 0.53) | 0.29 (0.06; 0.51) |  |  |  |  |
| Working memory (z-score) | HIIT+RT | 0.05 (-0.14; 0.25) | 0.34 (0.05; 0.63) | 0.29 (-0.06; 0.64) | 0.17 (-0.20; 0.53) | 0.38 | 0.25 (-0.11; 0.62) | 0.18 |
|  | HIIT | 0.05 (-0.14; 0.25) | 0.09 (-0.21; 0.39) | 0.04, (-0.33; 0.41) | -0.09, (-0.47; 0.29) | 0.65 |  |  |
|  | Usual care | 0.05, (-0.14; 0.25) | 0.18, (-0.12; 0.48) | 0.12, (-0.25; 0.49) |  |  |  |  |
| Executive function and attentional control (z-score) | HIIT+RT | 0.03 (-0.17; 0.23) | 0.17 (-0.08; 0.42) | 0.14 (-0.11; 0.39) | -0.13 (-0.39; 0.12) | 0.32 | -0.11 (-0.37; 0.15) | 0.41 |
|  | HIIT | 0.03 (-0.17; 0.23) | 0.28 (0.02; 0.54) | 0.25 (-0.01; 0.51) | -0.02 (-0.28; 0.24) | 0.88 |  |  |
|  | Usual care | 0.03 (-0.17; 0.23) | 0.30 (0.05; 0.55) | 0.27 (0.02; 0.52) |  |  |  |  |
| VO<sub>2</sub>peak (mL/kg/min) | HIIT+RT | 25.36 (24.30; 26.41) | 27.40 (26.17; 28.64) | 2.05 (1.00; 3.09) | 2.74 (1.64; 3.84) | <0.001 | 0.14 (-0.96; 1.25) | 0.8 |
|  | HIIT | 25.36 (24.30; 26.41) | 27.26 (26.02; 28.50) | 1.90 (0.83; 2.98) | 2.59 (1.48; 3.71) | <0.001 |  |  |
|  | Usual care | 25.36 (24.30; 26.41) | 24.67 (23.43; 25.91) | -0.69 (-1.75; 0.38) |  |  |  |  |
| 30s sit-to-stand (repetitions) | HIIT+RT | 16.78 (15.82; 17.75) | 21.69 (20.41; 22.98) | 4.91 (3.53; 6.29) | 3.32 (1.89; 4.75) | <0.001 | 2.95 (1.51; 4.39) | <0.001 |
|  | HIIT | 16.78 (15.82; 17.75) | 18.74 (17.44; 20.04) | 1.95 (0.55; 3.36) | 0.36 (-1.08; 1.81) | 0.62 |  |  |
|  | Usual care | 16.78 (15.82; 17.75) | 18.38 (17.09; 19.66) | 1.59 (0.21; 2.97) |  |  |  |  |
| Weight (kg) | HIIT+RT | 82.89 (80.28; 85.51) | 83.30 (80.63; 85.98) | 0.41 (-0.51; 1.34) | 0.01 (-0.97; 0.98) | 0.99 | 0.89 (-0.09; 1.88) | 0.08 |
|  | HIIT | 82.89 (80.28; 85.51) | 82.41 (79.73; 85.09) | -0.48 (-1.43; 0.47) | -0.89 (-1.88; 0.11) | 0.09 |  |  |
|  | Usual care | 82.89 (80.28; 85.51) | 83.30 (80.62; 85.97) | 0.41 (-0.53; 1.34) |  |  |  |  |
| BMI (kg/m<sup>2</sup>) | HIIT+RT | 29.06 (28.27; 29.84) | 29.26 (28.44; 30.07) | 0.20 (-0.17; 0.57) | -0.03 (-0.42; 0.36) | 0.88 | 0.41 (0.02; 0.81) | 0.05 |
|  | HIIT | 29.06 (28.27; 29.84) | 28.84 (28.02; 29.66) | -0.21 (-0.60; 0.17) | -0.44 (-0.84; -0.04) | 0.03 |  |  |
|  | Usual care | 29.06 (28.27; 29.84) | 29.29 (28.47; 30.10) | 0.23 (-0.15; 0.61) |  |  |  |  |
| Fat mass (kg) | HIIT+RT | 23.86 (22.46; 25.25) | 23.82 (22.34; 25.30) | -0.04 (-0.85; 0.77) | -0.06 (-0.91; 0.79) | 0.90 | 0.73 (-0.13; 1.58) | 0.10 |
|  | HIIT | 23.86 (22.46; 25.25) | 23.09 (21.61; 24.58) | -0.77 (-1.59; 0.06) | -0.79 (-1.44; 0.07) | 0.08 |  |  |
|  | Usual care | 23.86 (22.46; 25.25) | 23.88 (22.40; 25.36) | 0.02 (-0.79; 0.83) |  |  |  |  |
| Muscle mass (kg) | HIIT+RT | 55.62 (53.93; 57.32) | 56.02 (54.29; 57.76) | 0.40 (-0.23; 1.03) | -0.05 (-0.71; 0.61) | 0.88 | 0.19 (-0.47; 0.86) | 0.57 |
|  | HIIT | 55.62 (53.93; 57.32) | 55.83 (54.09; 57.57) | 0.21 (-0.23; 0.84) | -0.24 (-0.91; 0.42) | 0.47 |  |  |
|  | Usual care | 55.62 (53.93; 57.32) | 56.08 (54.34; 57.81) | 0.45 (-0.18; 1.08) |  |  |  |  |
| FMI (kg/m<sup>2</sup>) | HIIT+RT | 8.54 (7.97; 9.12) | 8.27 (7.61; 8.93) | 0.02 (-0.28; 0.32) | -0.03 (-0.35; 0.29) | 0.86 | 0.29 (-0.03; 0.61) | 0.08 |
|  | HIIT | 8.54 (7.97; 9.12) | 8.28 (7.61; 8.95) | -0.27 (-0.58; 0.04) | -0.32 (-0.64; 0.00) | 0.06 |  |  |
|  | Usual care | 8.54 (7.97; 9.12) | 8.59 (7.93; 9.25) | 0.05 (-0.26; 0.35) |  |  |  |  |
| MMI (kg/m<sup>2</sup>) | HIIT+RT | 19.41 (19.05; 19.77) | 19.95 (19.46; 20.44) | 0.18 (-0.05; 0.40) | -0.02 (-0.26; 0.21) | 0.84 | 0.15 (-0.09; 0.38) | 0.23 |
|  | HIIT | 19.41 (19.05; 19.77) | 19.51 (19.01; 20.01) | 0.03 (-0.20; 0.26) | -0.17 (-0.41; 0.07) | 0.16 |  |  |
|  | Usual care | 19.41 (19.05; 19.77) | 19.70 (19.20; 20.19) | 0.20 (-0.02; 0.42) |  |  |  |  |

Values indicate estimated marginal means at each time point (and 95% Confidence Intervals).
Abbreviations: BMI, body mass index; CBF, Cerebral blood flow; CI, confidence intervals; FMI, fat mass index; HIIT, High Intensity Interval Training; HIIT+RT, high intensity interval training + resistance training; MMI, muscle mass index; MoCA, Montreal Cognitive Assessment; VO<sub>2</sub>peak, peak oxygen uptake.

**Figure S1.**
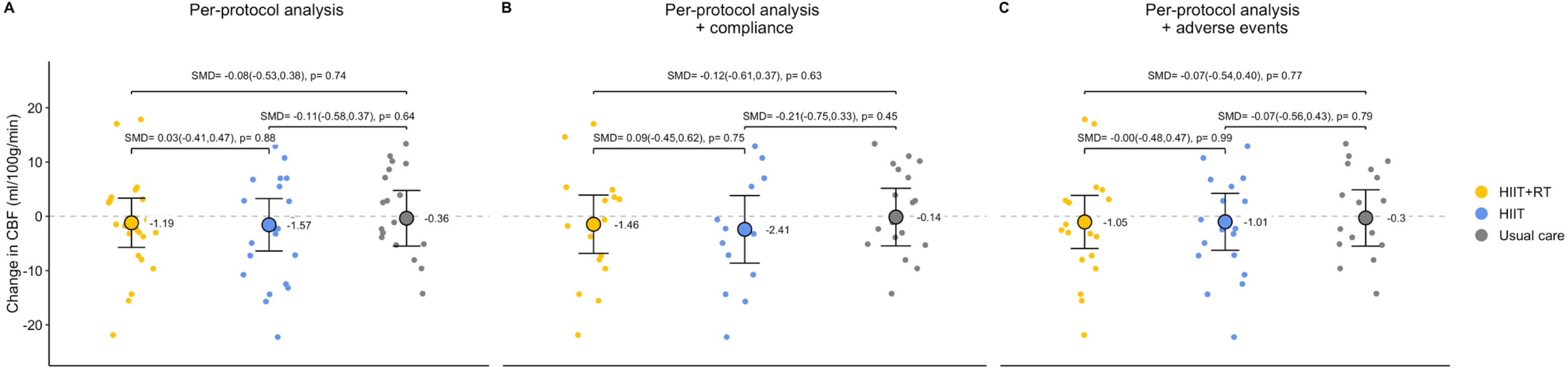
A. Per-protocol (participants meeting ≥70% attendance) effects of the 12-week interventions on CBF by intervention group (HIIT+RT, HIIT, Usual care). B. Sensitivity analyses based on per-protocol + compliance database (per-protocol plus excluding participants not achieving ≥70% compliant sessions) effects of the 12-week interventions on CBF by intervention group (HIIT+RT, HIIT, Usual care). C. Sensitivity analyses based on per-protocol + adverse events (per-protocol plus excluding participants who experienced adverse events at post-assessments, had deviations from the exercise protocol, or underwent interruption periods due to adverse events) effects of the 12-week interventions on CBF by intervention group (HIIT+RT, HIIT, Usual care). The figure shows adjusted mean change over time and 95% Confidence Intervals by intervention groups, and dots cloud indicate participants individual raw change values. P-values are shown above the plots, connecting the corresponding study groups. Abbreviations: CBF, cerebral blood flow; HIIT, high intensity interval training; HIIT+RT, high intensity interval training + resistance training; SMD, standardized mean difference.

**Table S8.**
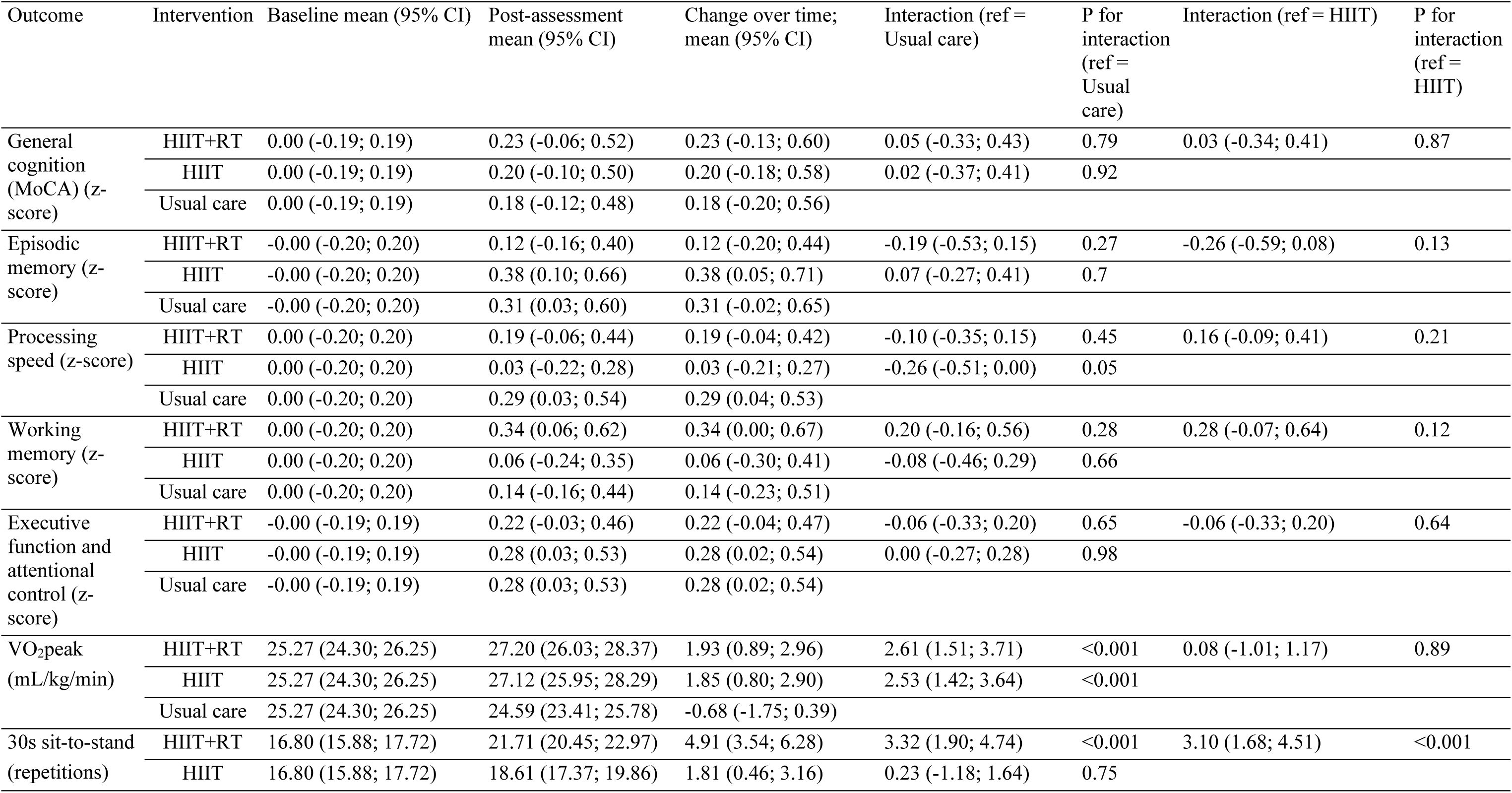
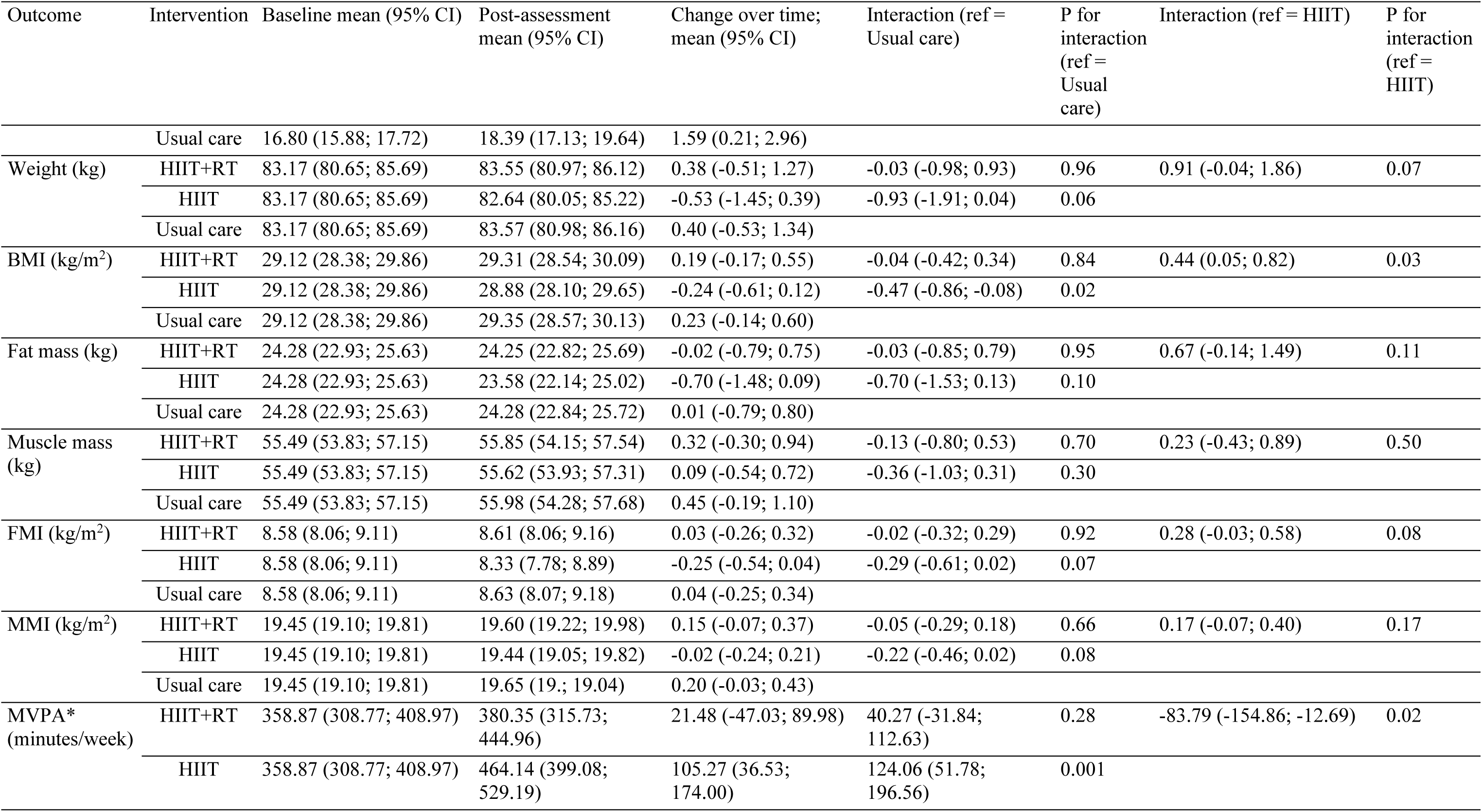
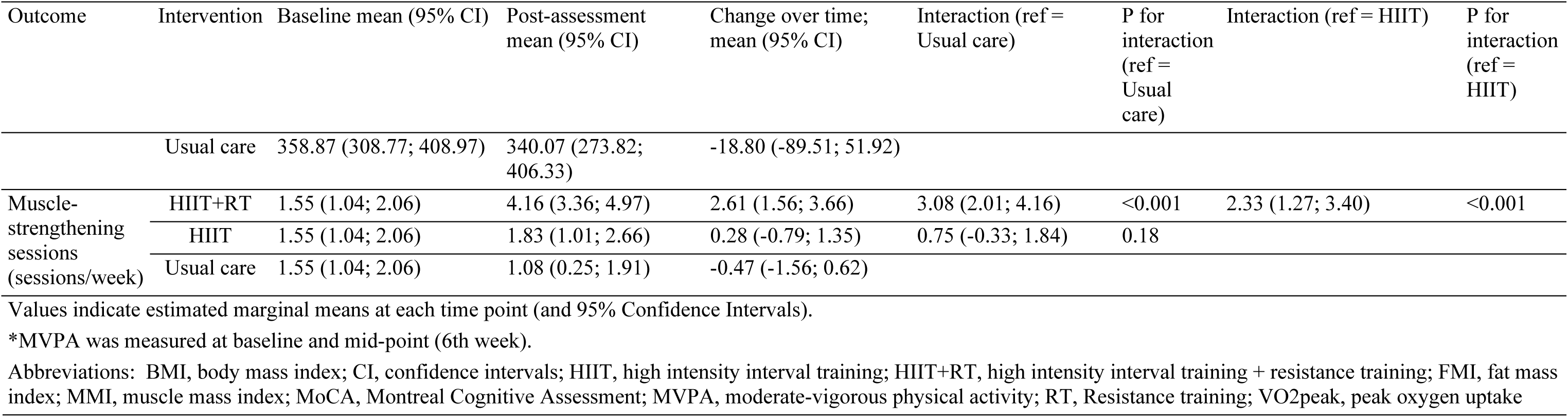
Intention-to-treat effects of the 12-week intervention on general cognition, cognitive domains, VO2peak, 30s sit-to-stand repetitions, body weight, BMI, fat mass, muscle mass, FMI, MMI, MVPA and muscle-strengthening sessions.

**Table S9.**
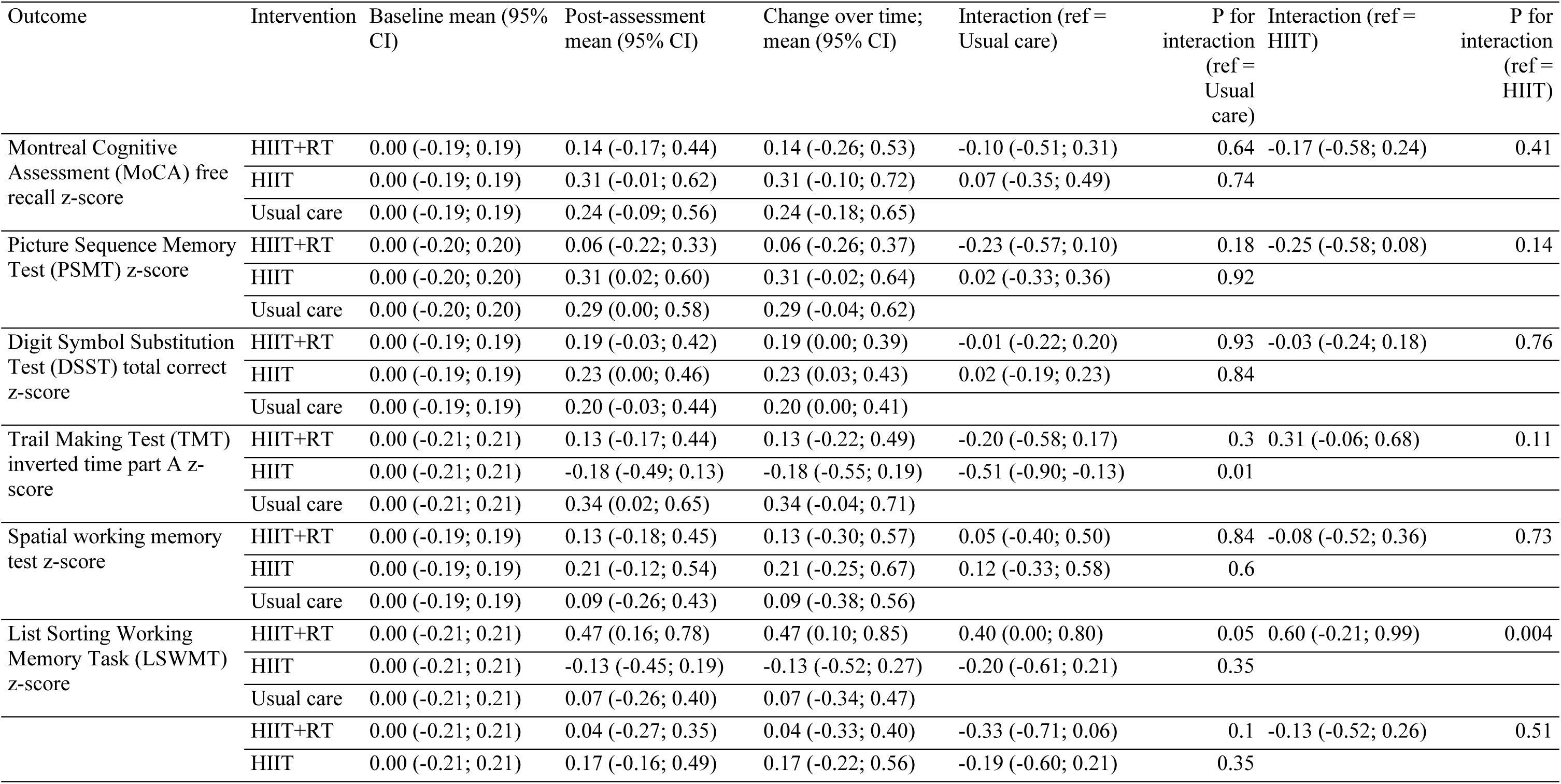
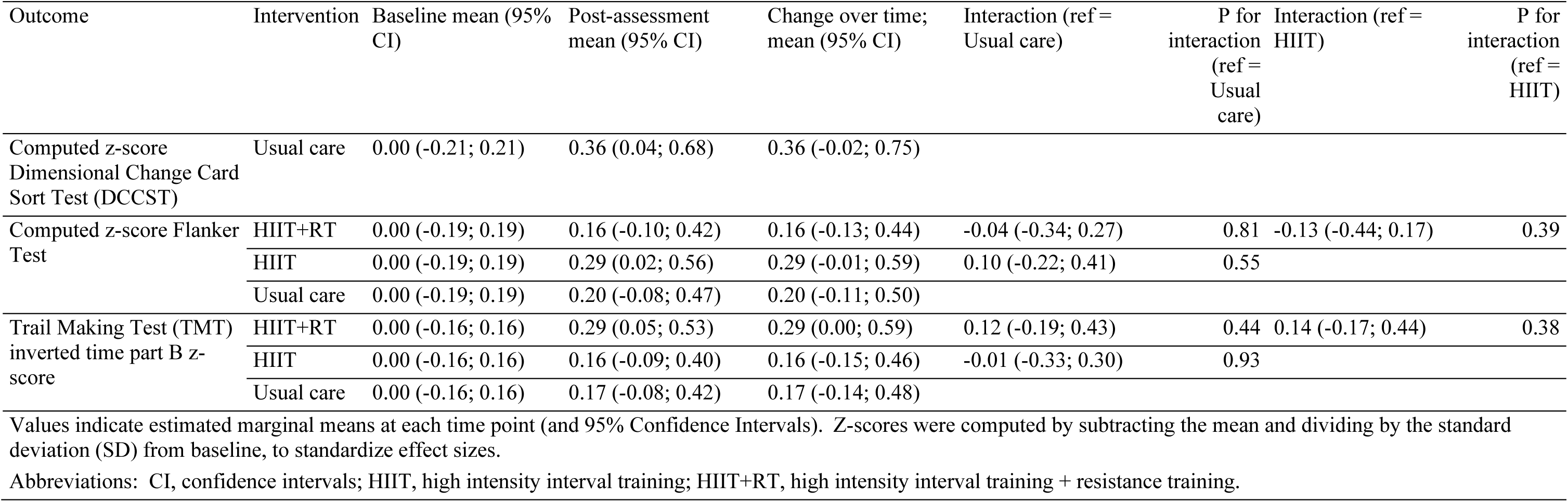
Intention-to-treat effects of the 12-week intervention on the individual cognitive tests used to compute the cognitive domains (z-scores)

**Table S10.**
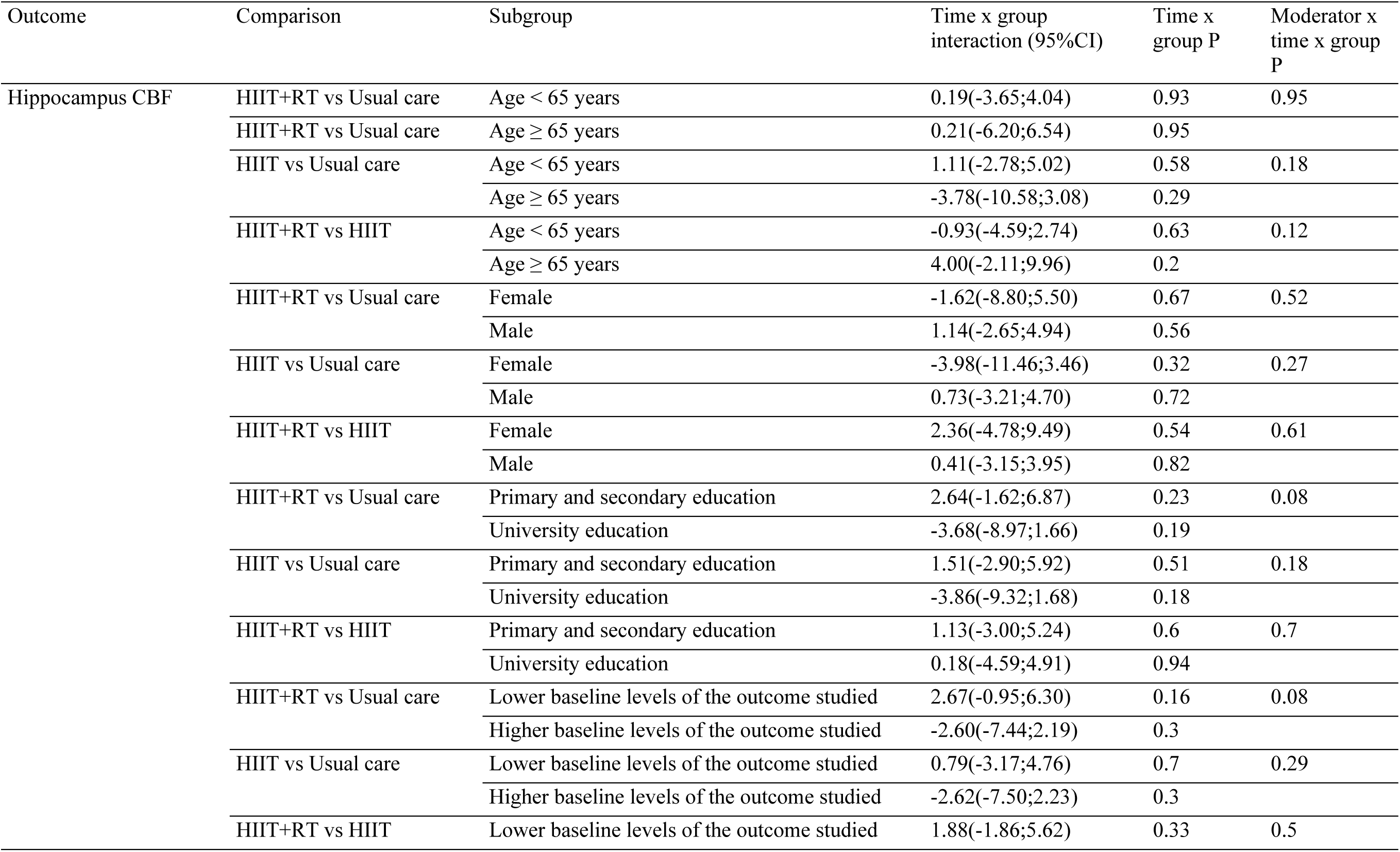
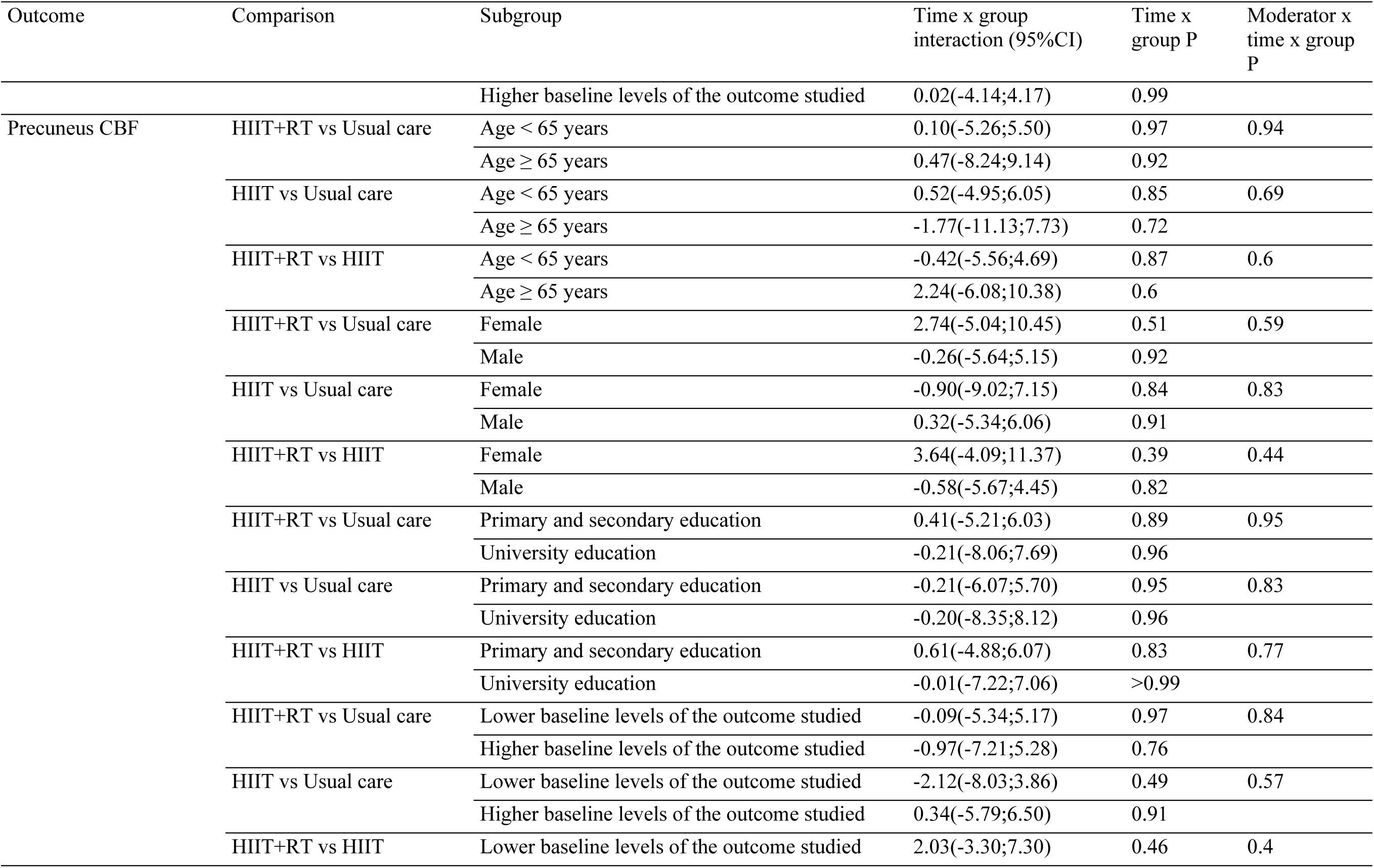
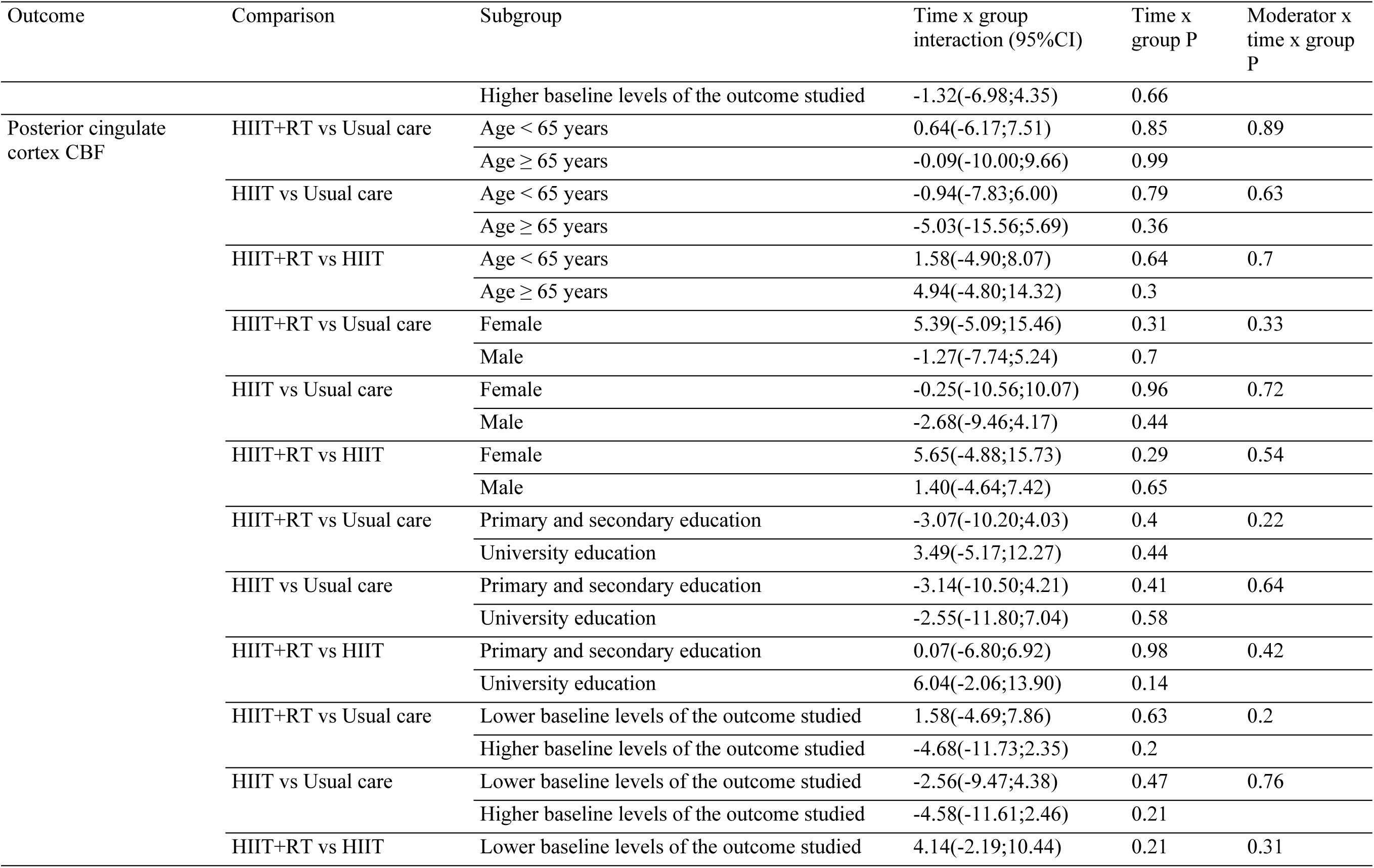
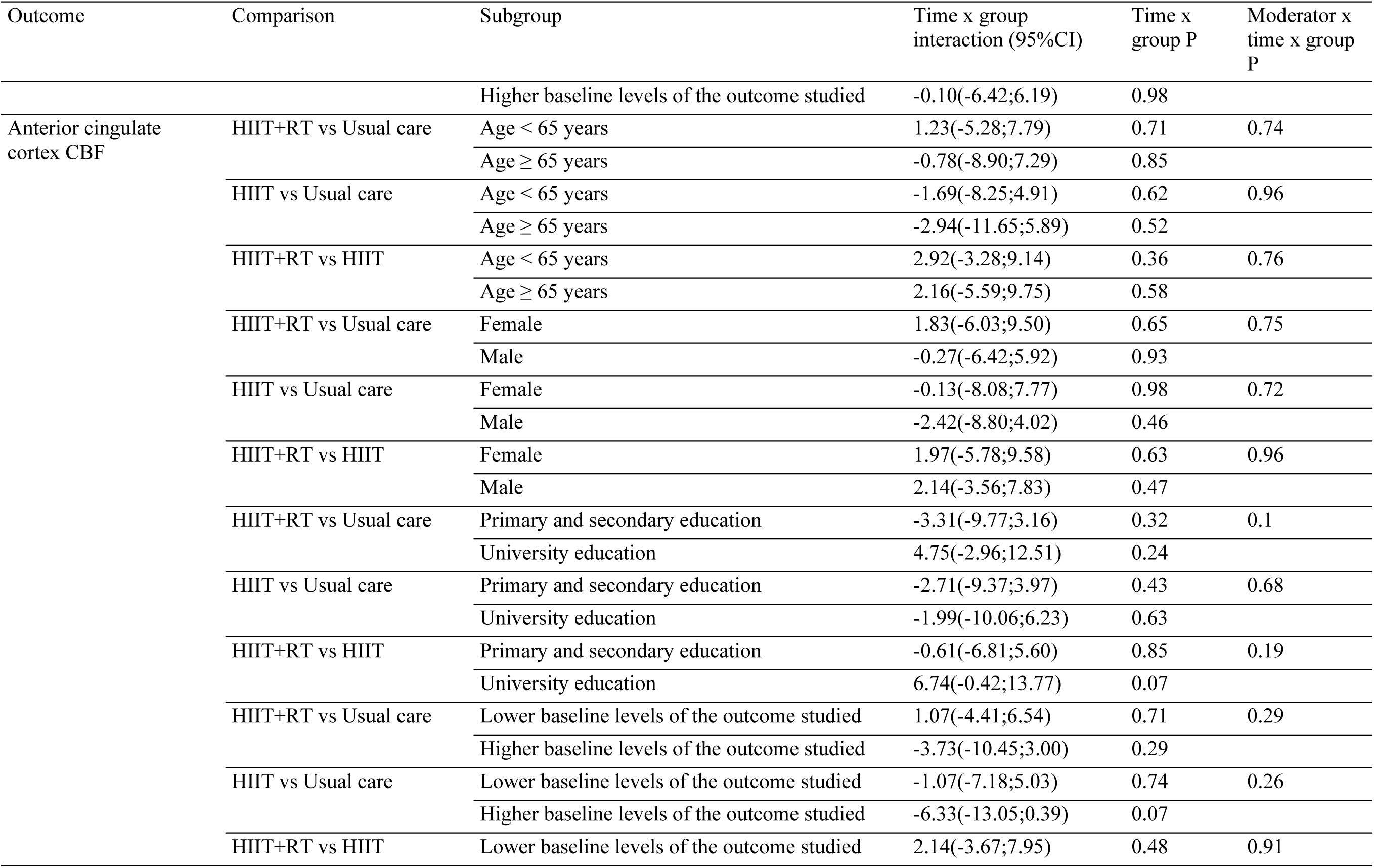
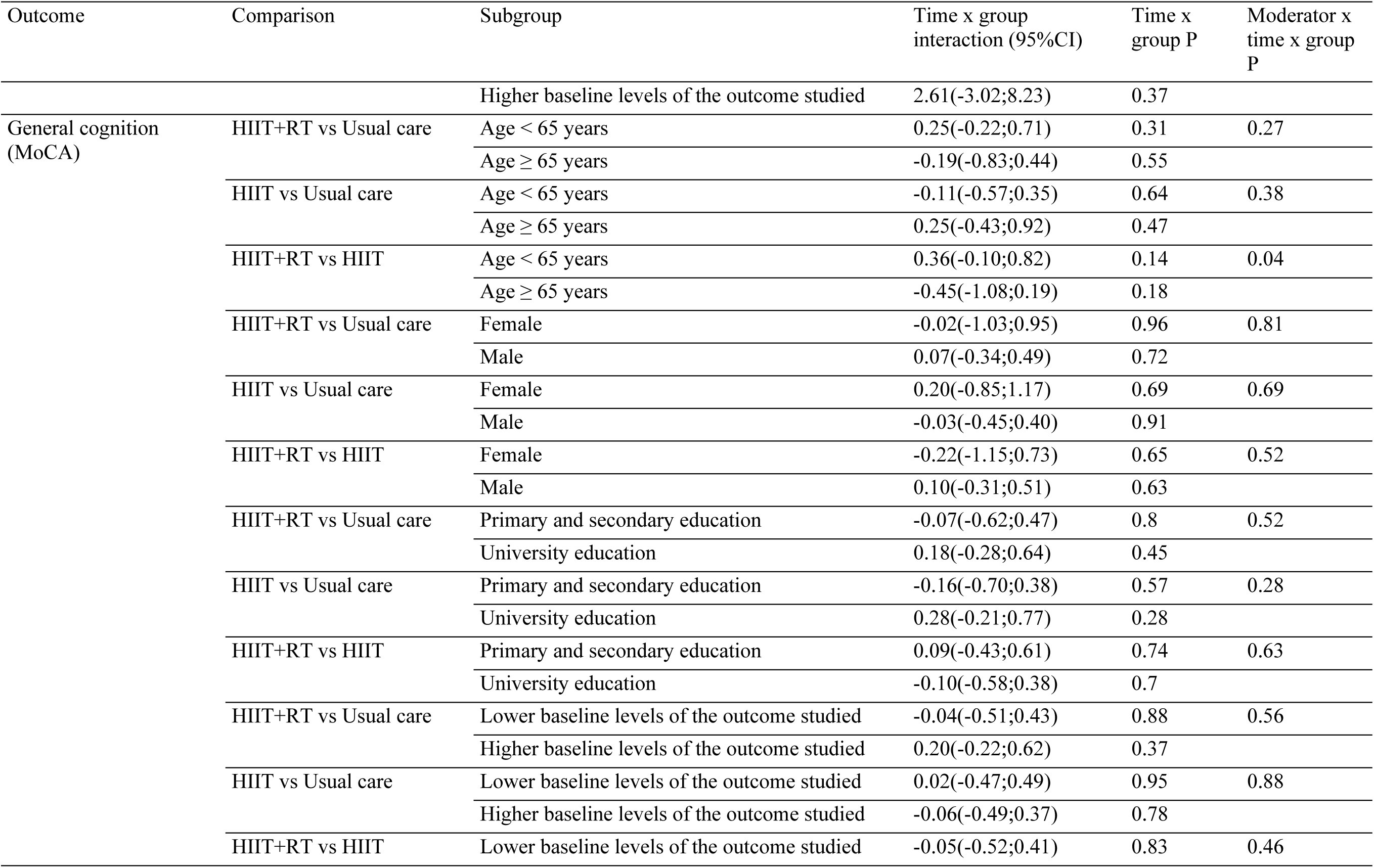
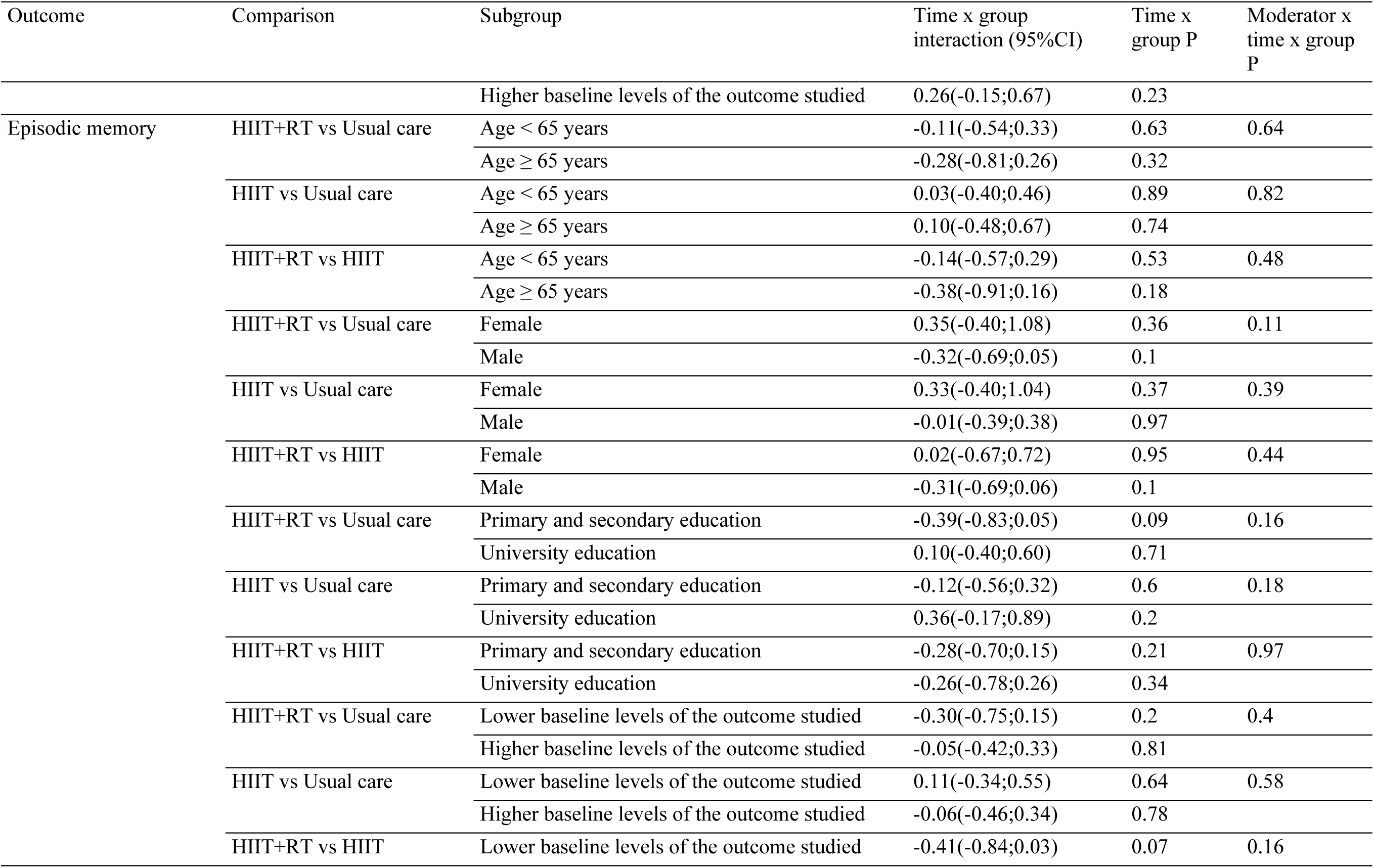
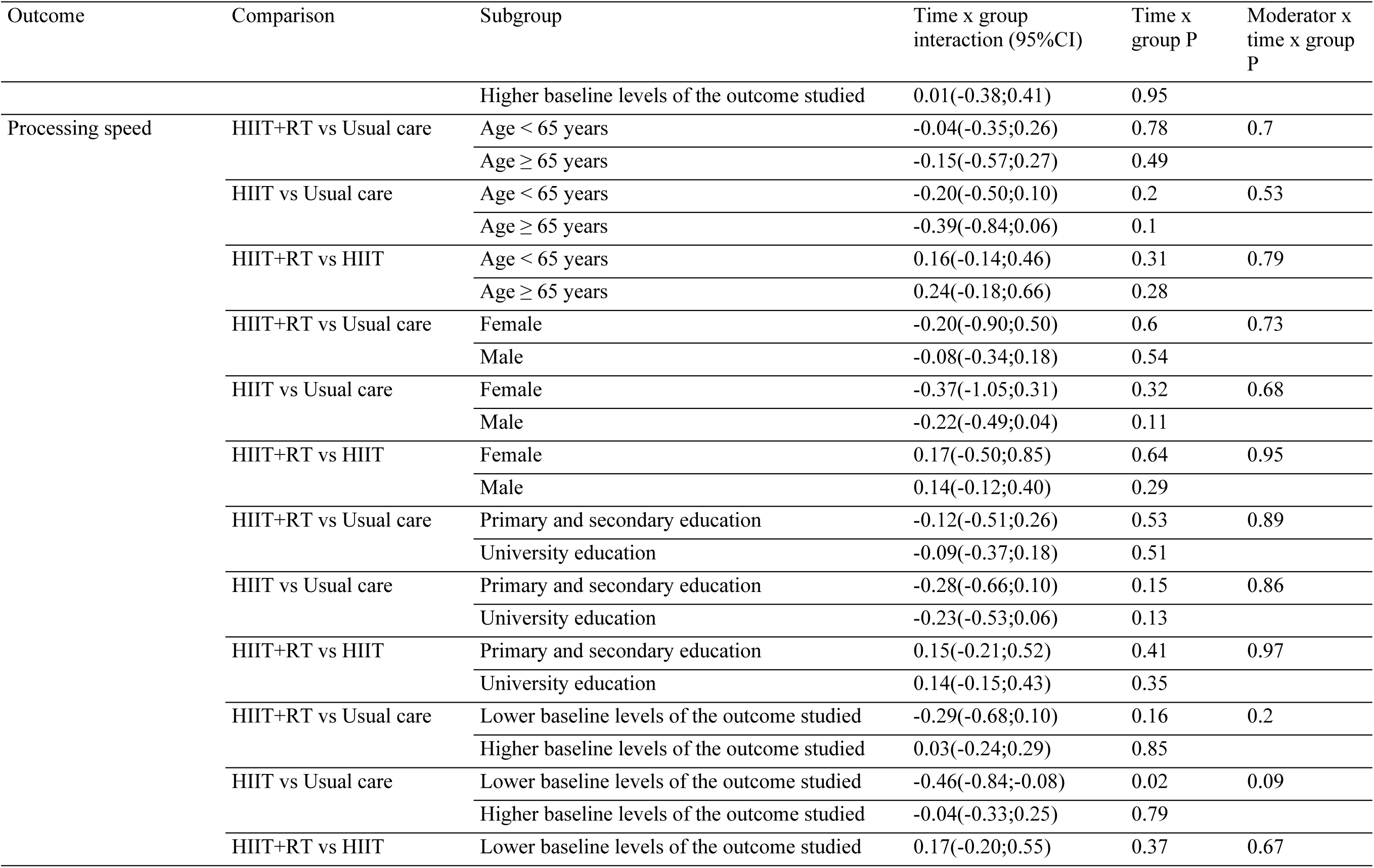
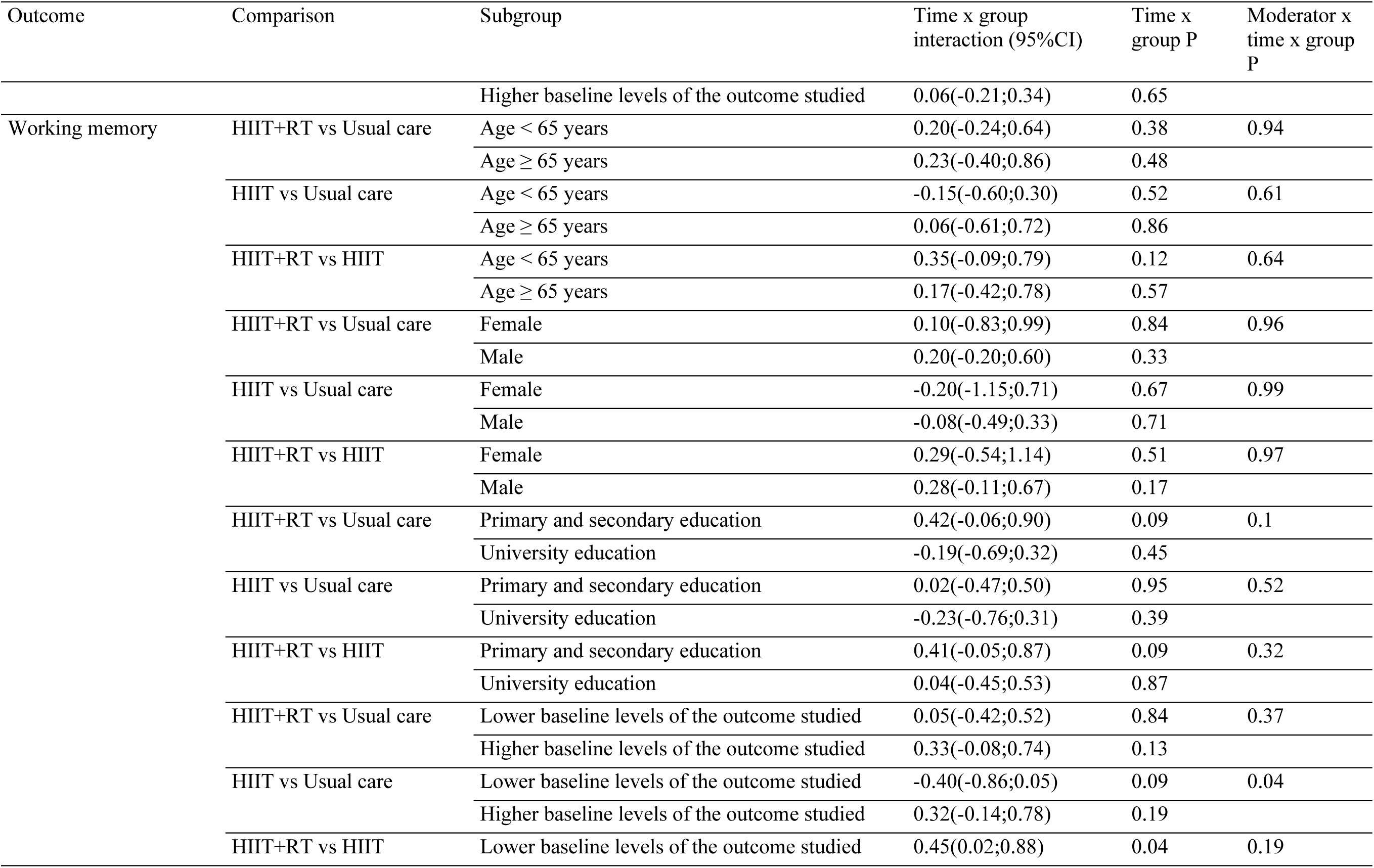
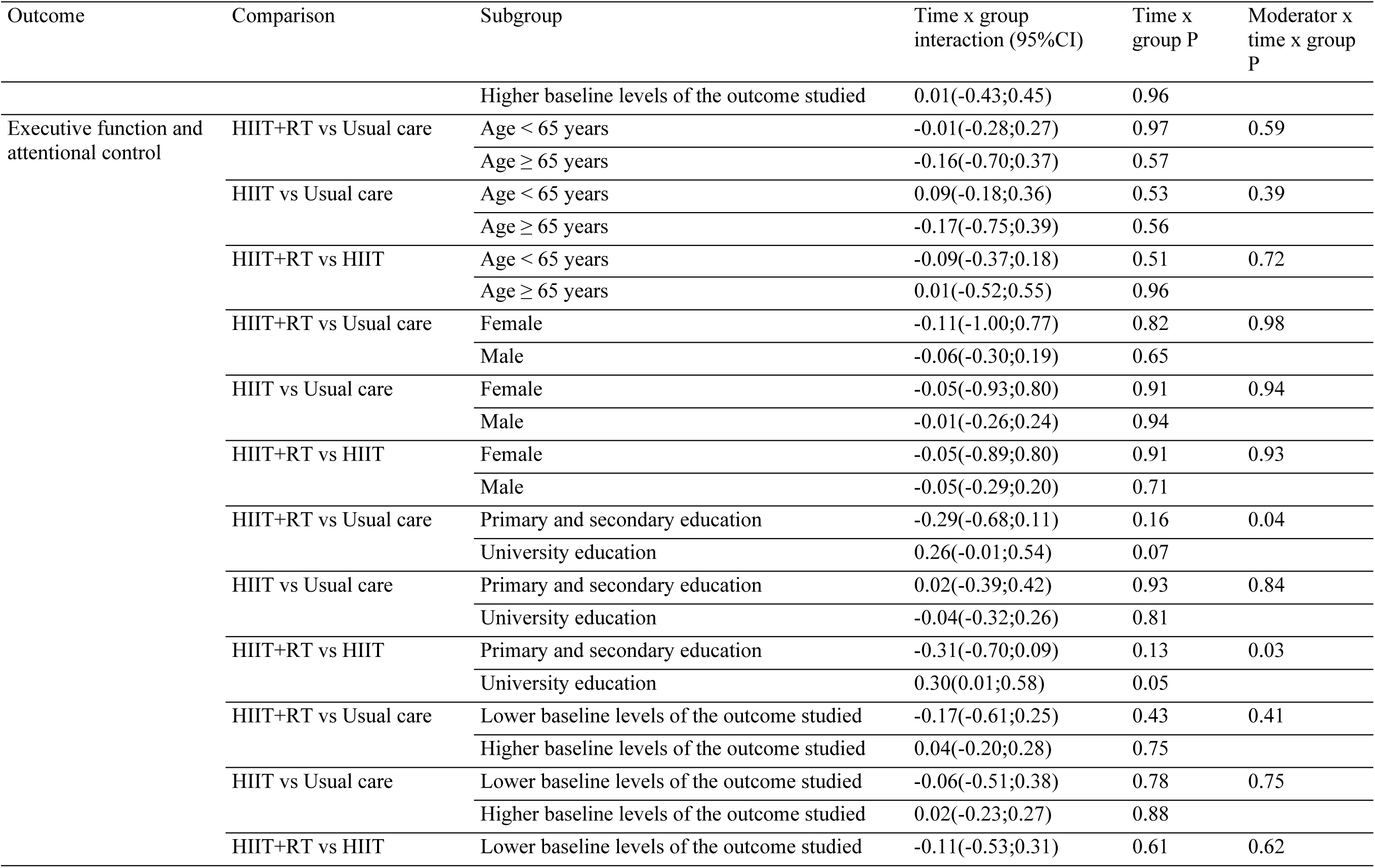
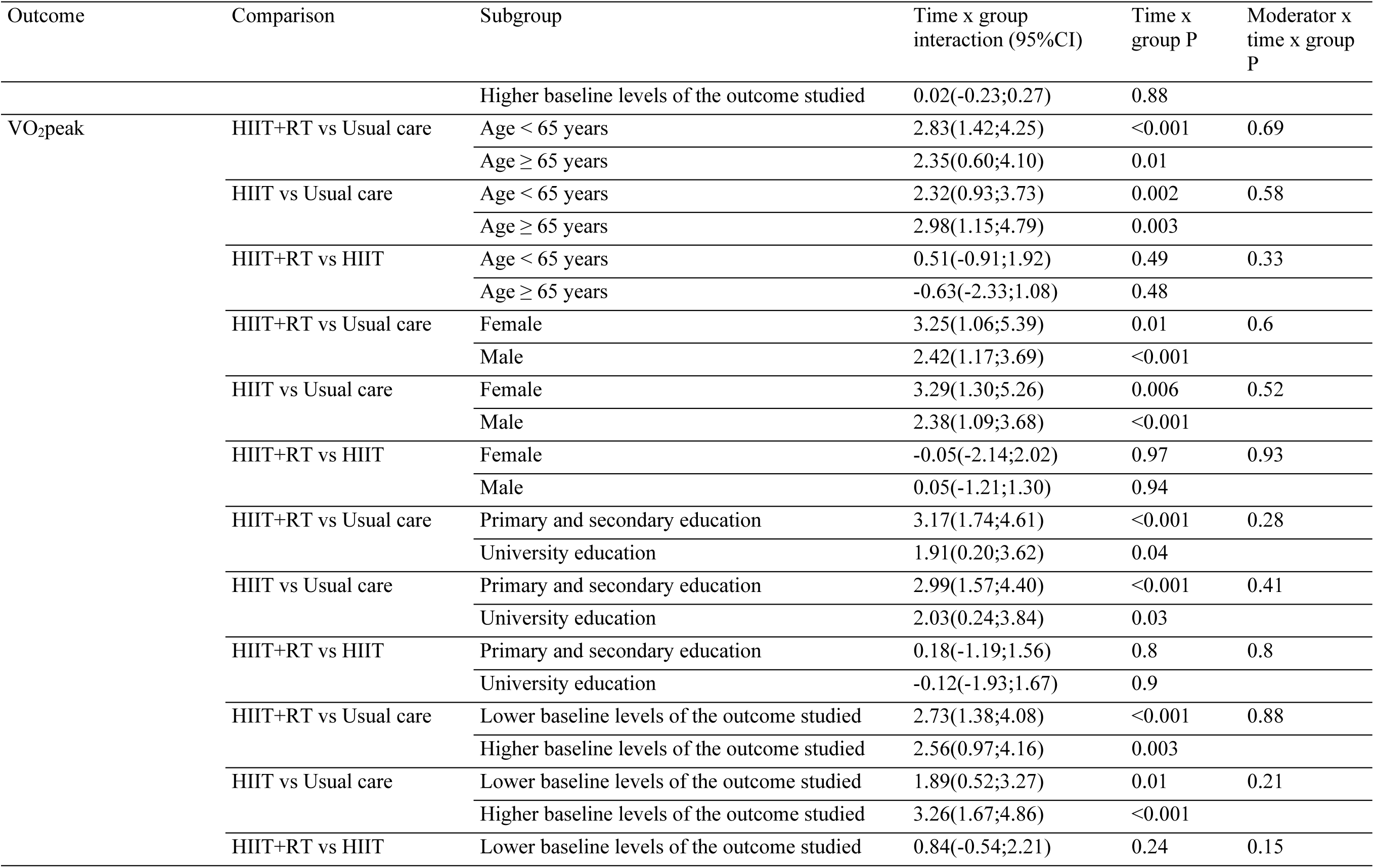
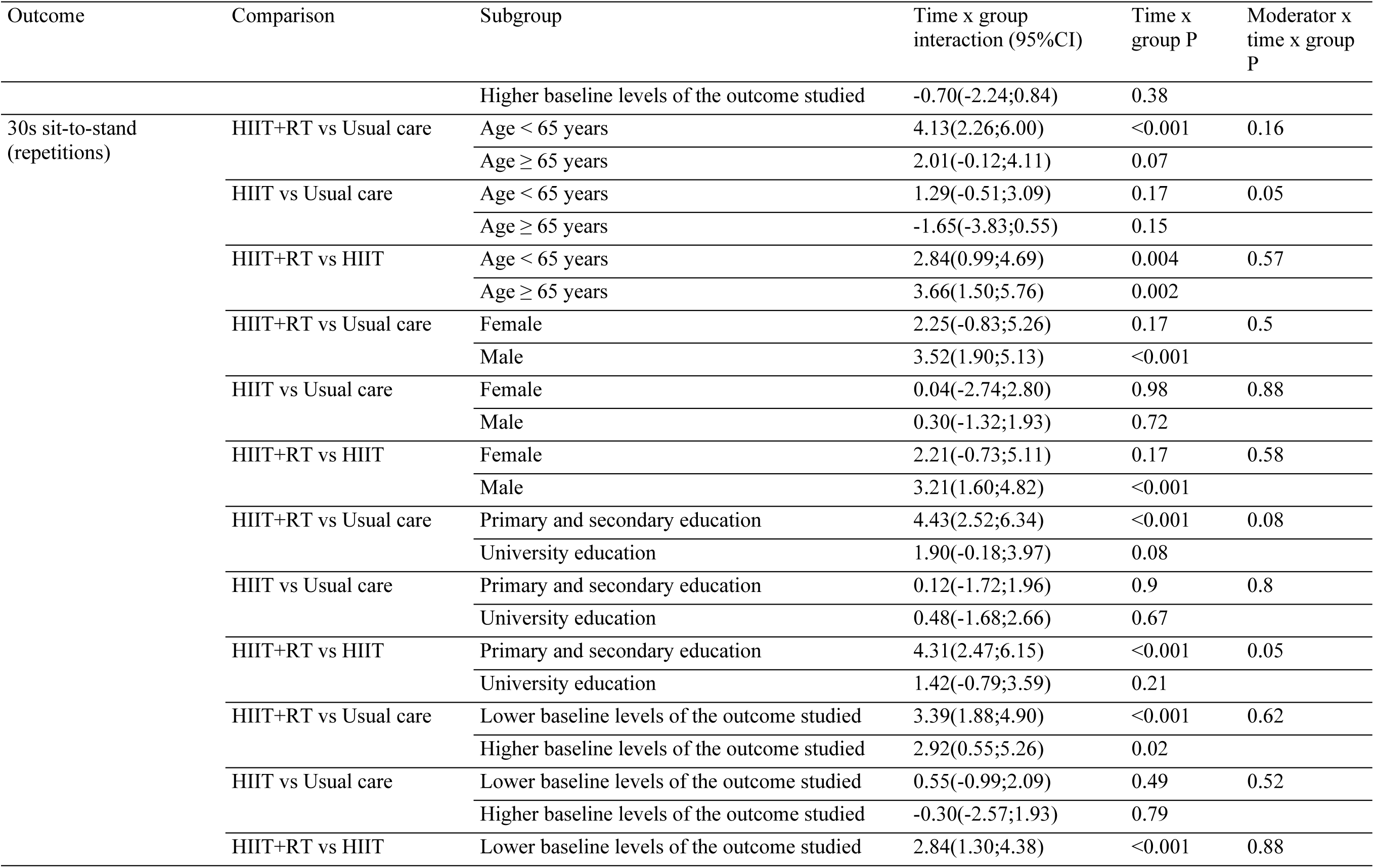
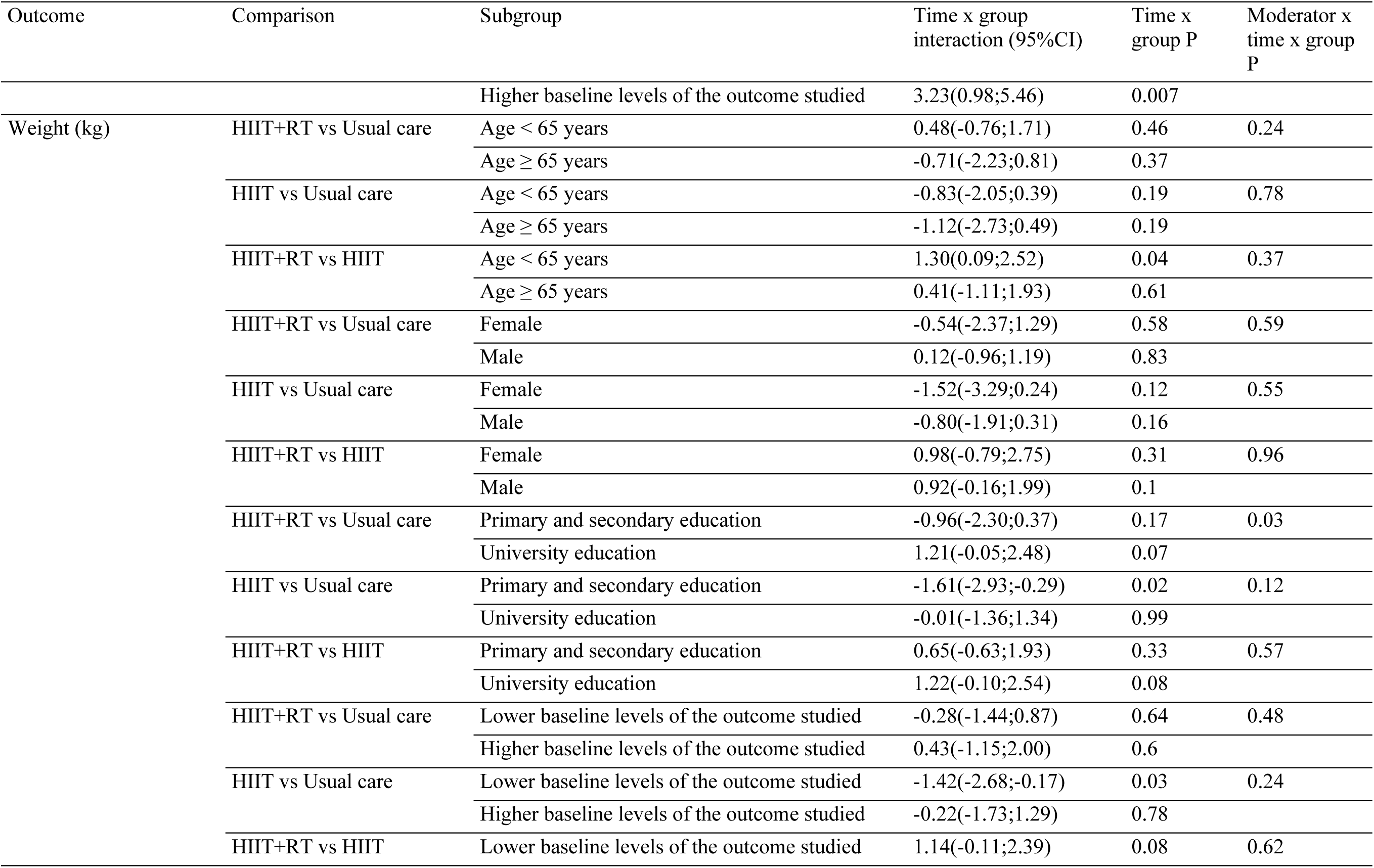
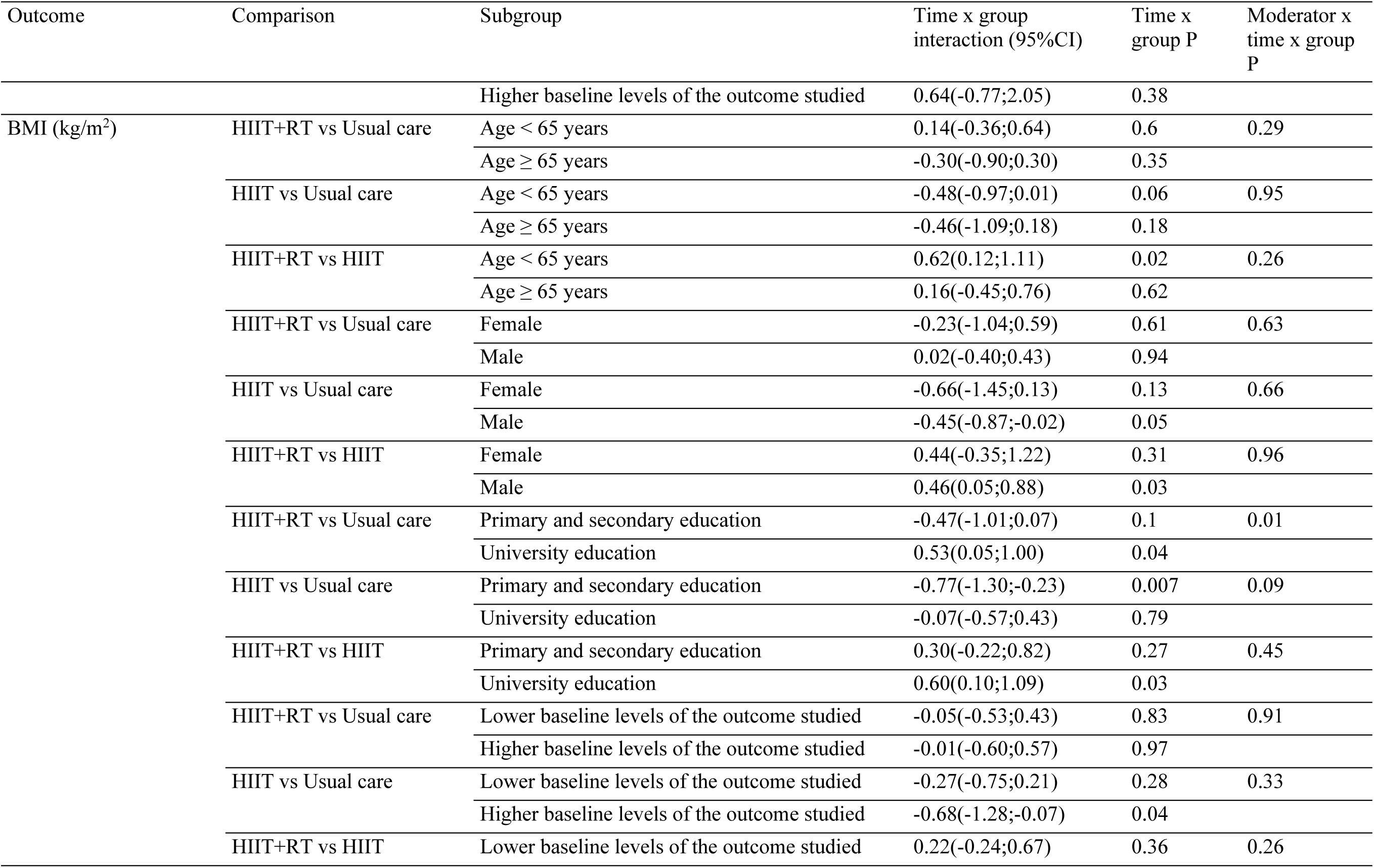
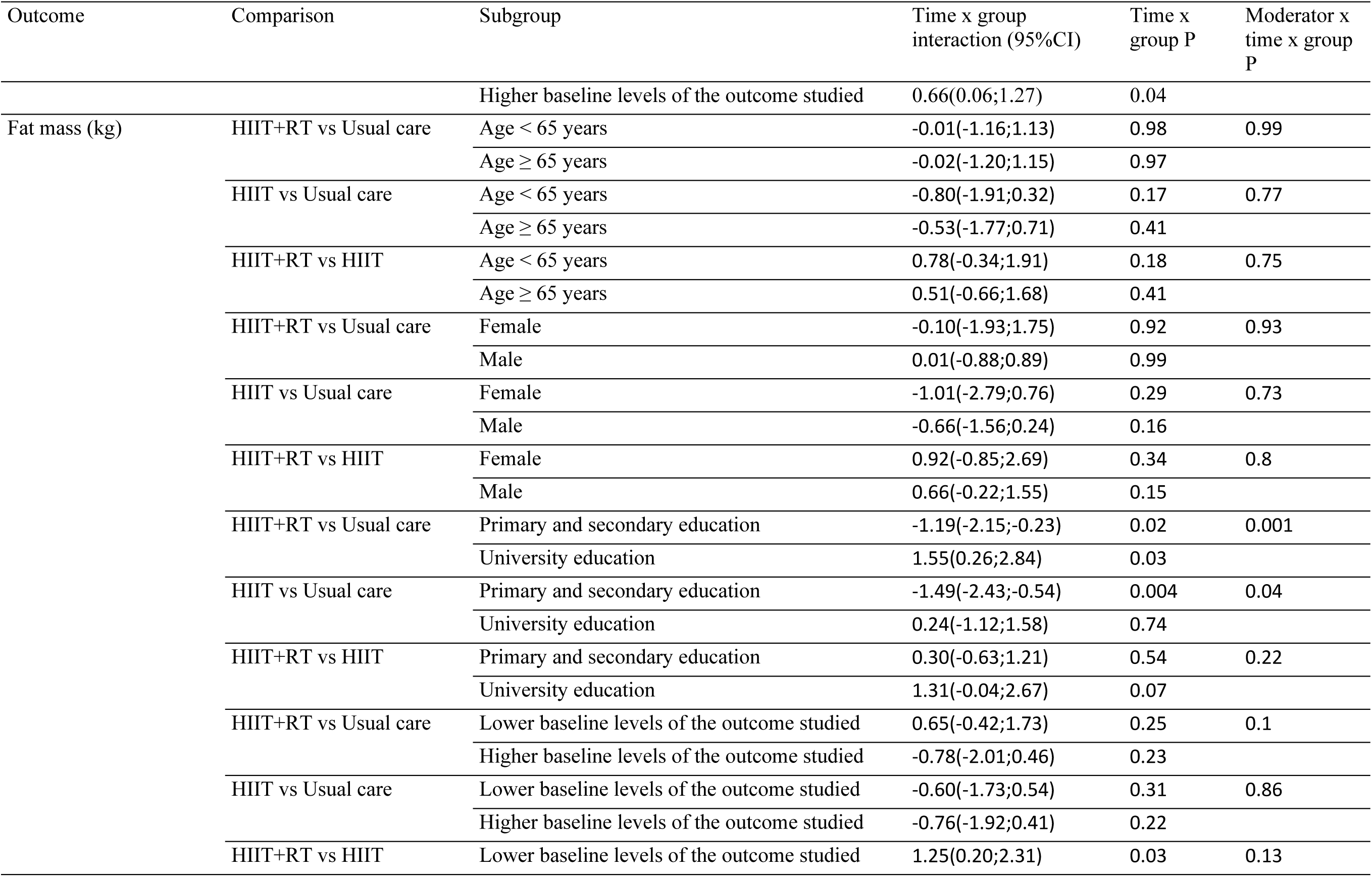
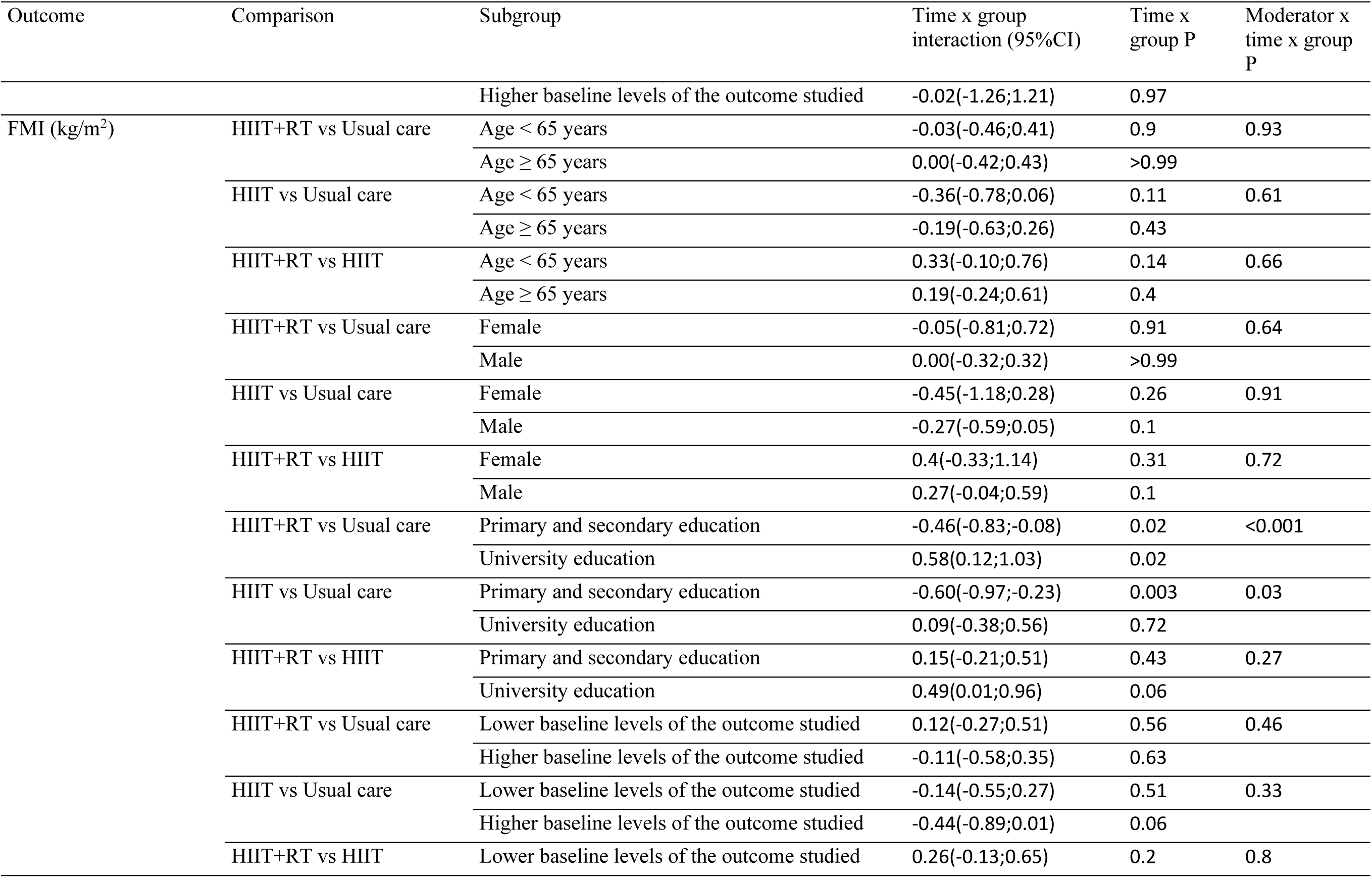
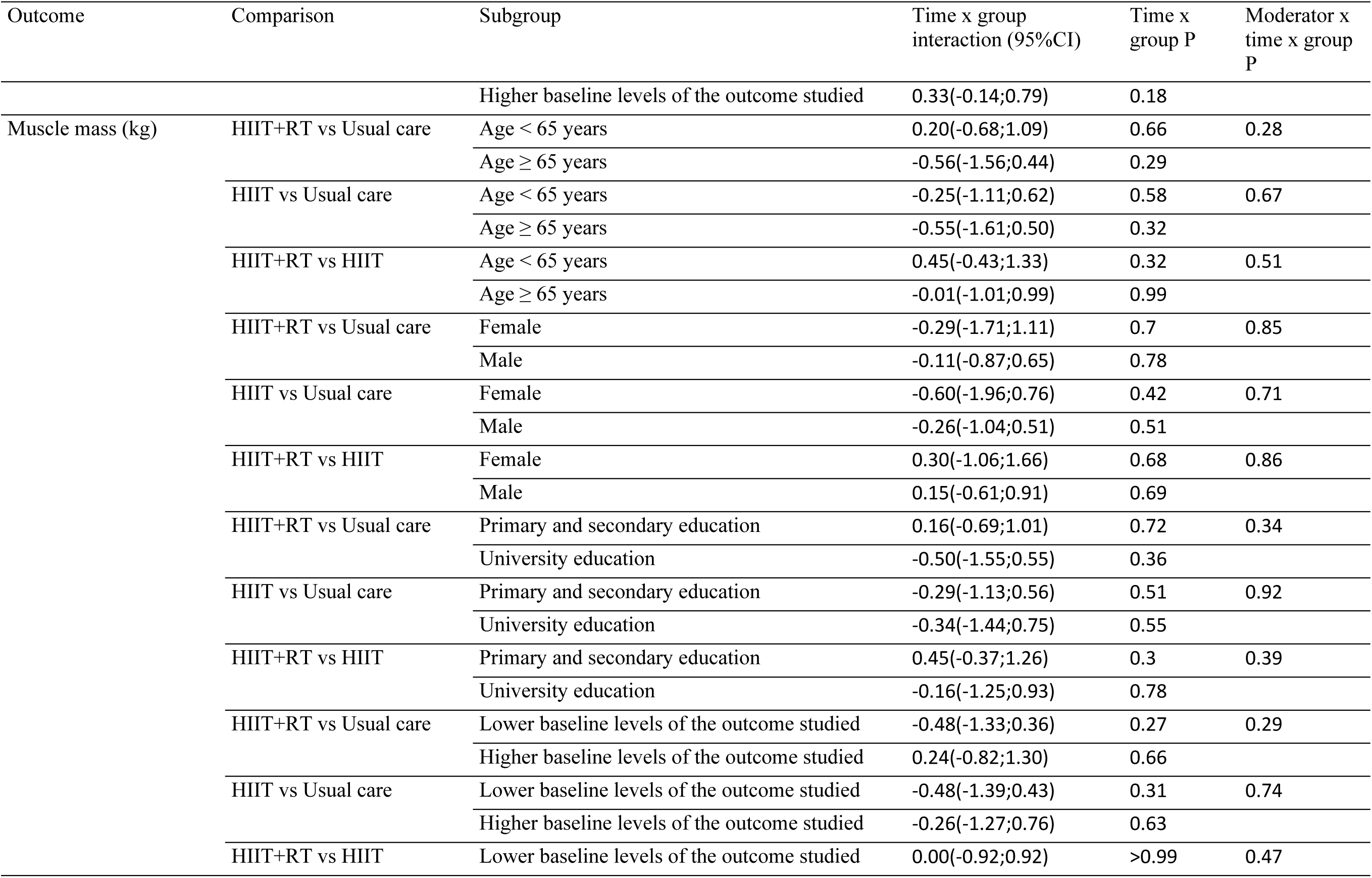
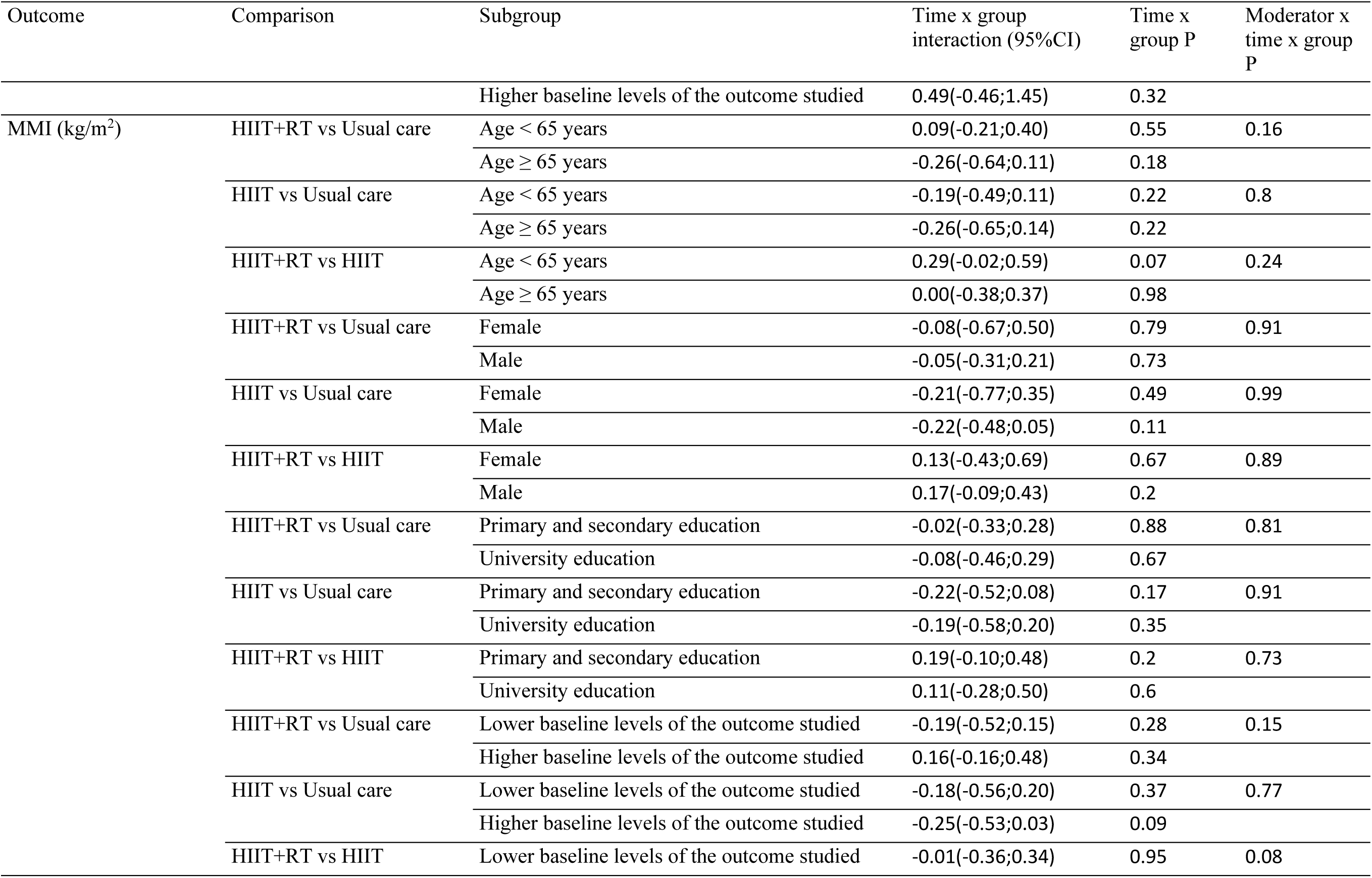
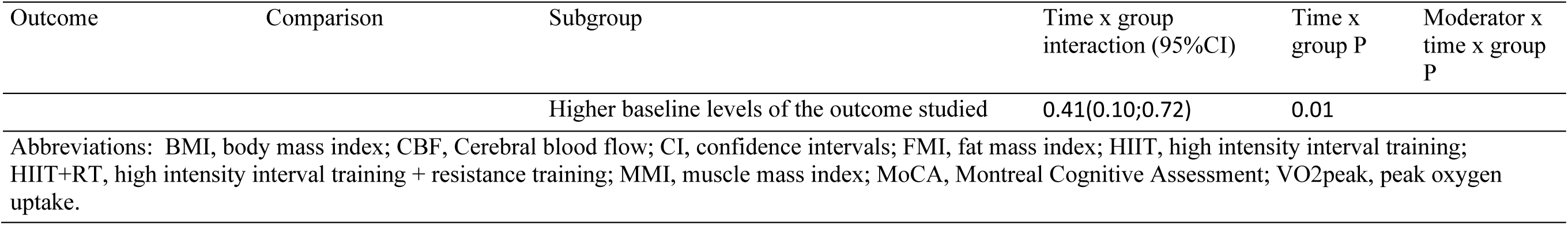
Moderation analyses three-way interaction effect on secondary outcomes.

**Figure S2.**
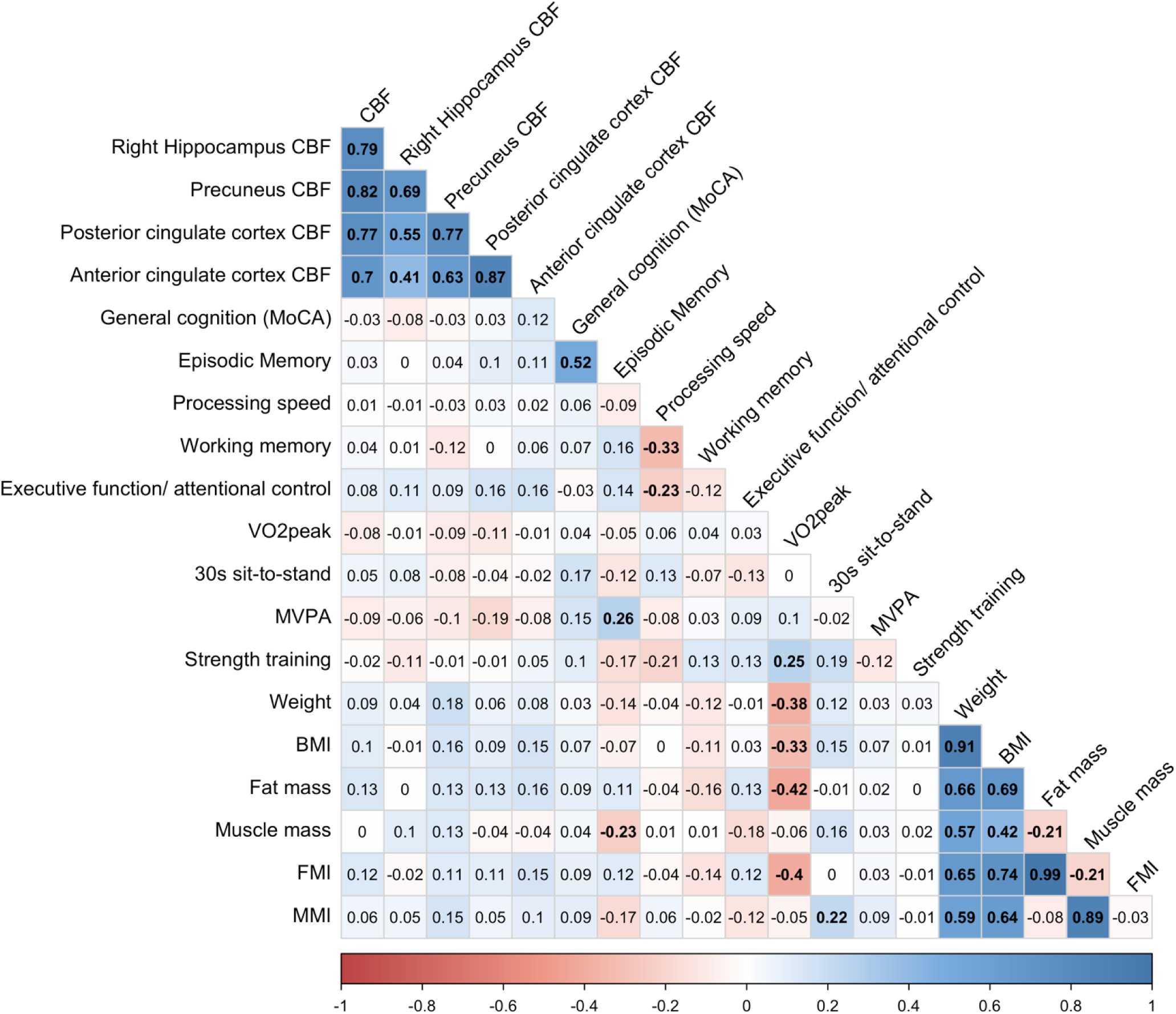
Matrix of Pearson correlation coefficients (r) between changes on primary and secondary outcomes. Numbers in bold indicate statistically significant correlations (p<0.05), the colours indicate the strength of the correlation, as shown in the explanatory scale on the bottom of the figure. BMI, body mass index; CBF, Cerebral blood flow; CI, confidence intervals; FMI, fat mass index; HIIT, high intensity interval training; HIIT+RT, high intensity interval training + resistance training; MMI, muscle mass index; MVPA, moderate-vigorous physical activity; MoCA, Montreal Cognitive Assessment; RT, Resistance training; VO2peak, peak oxygen uptake.

**Table S11.**
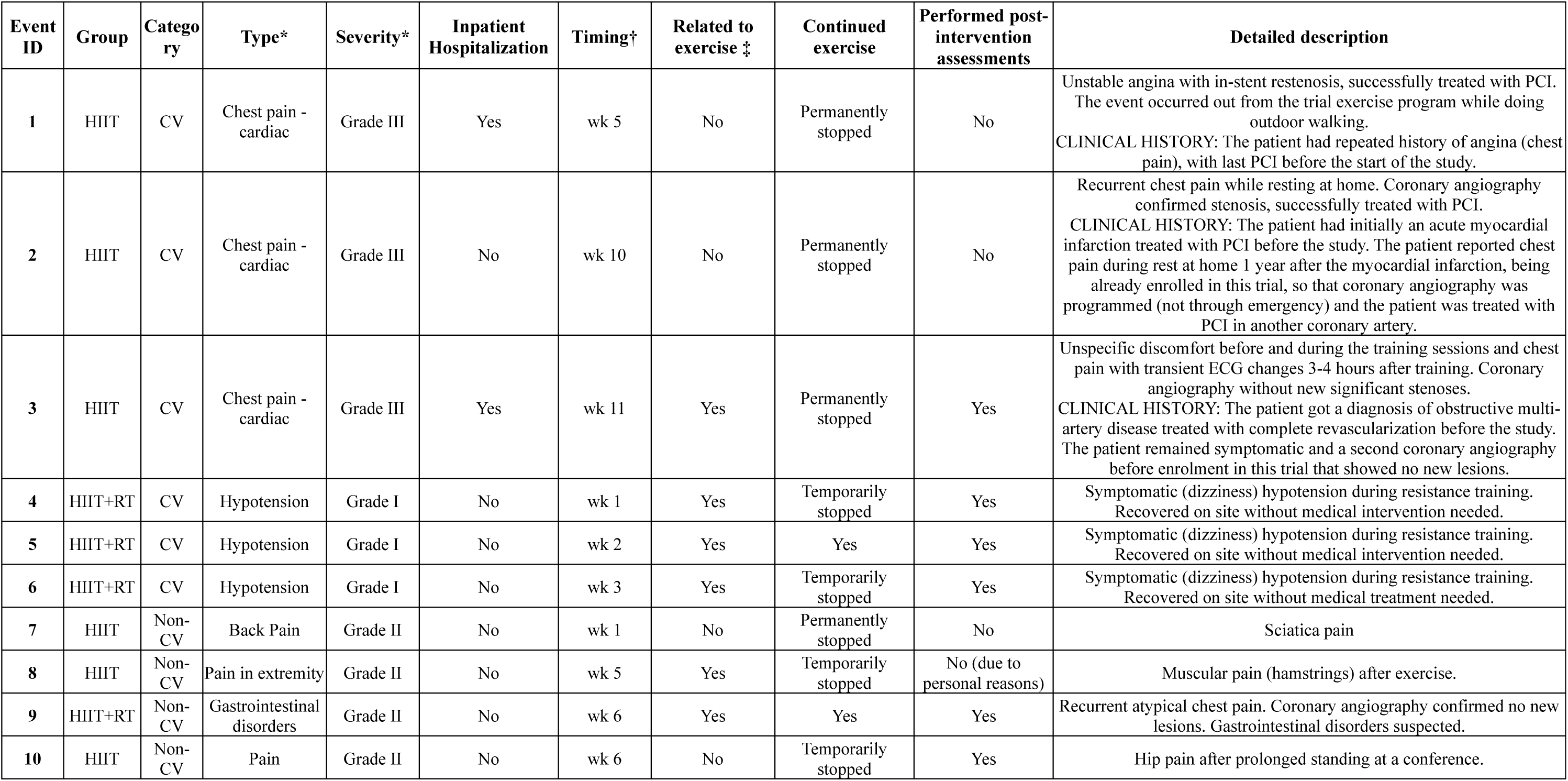
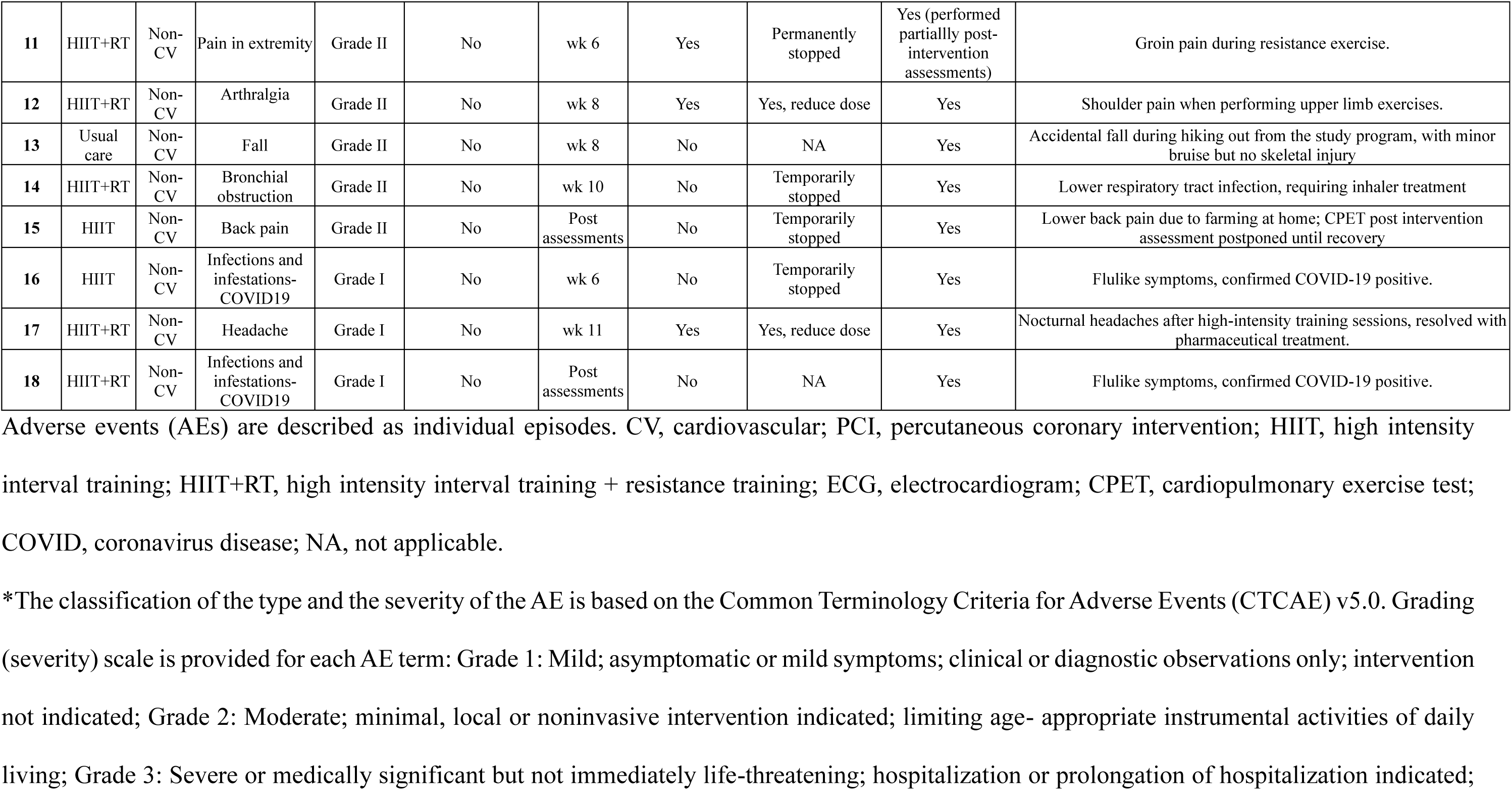
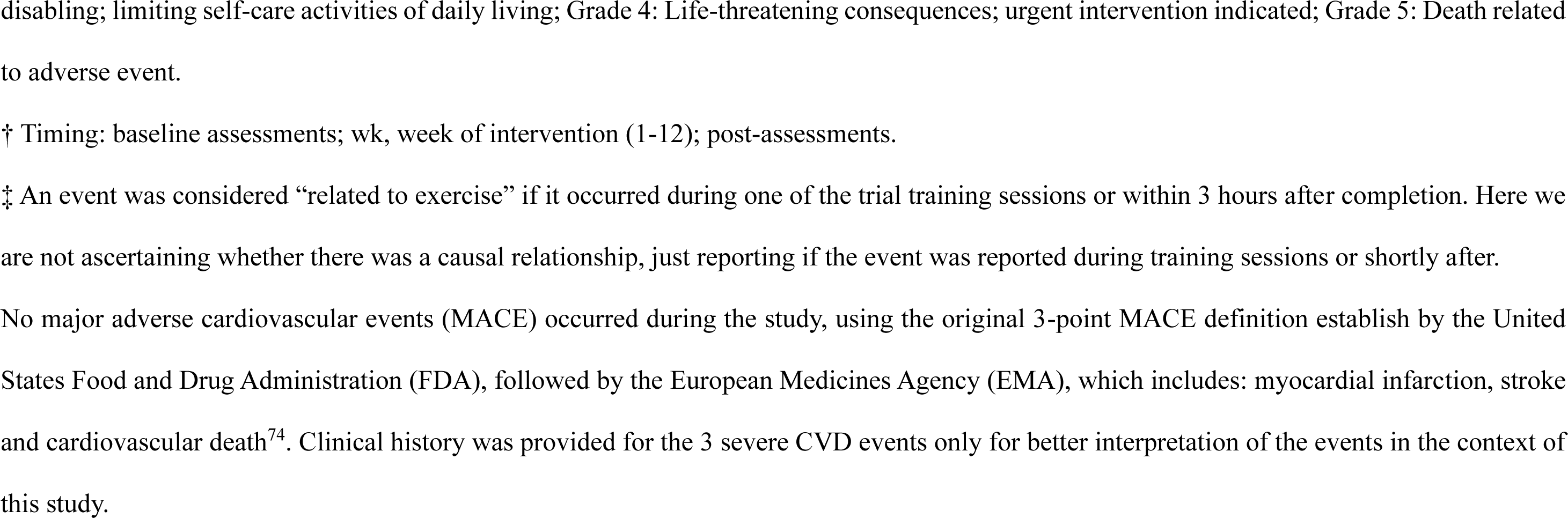
Adverse events registered during the study.

